# Multidomain lifestyle interventions for the risk reduction of cognitive impairment and dementia: A systematic review and meta-analysis

**DOI:** 10.64898/2026.07.12.26357861

**Authors:** Mariagnese Barbera, Ruth Stephen, Esko Levälahti, Jenni Lehtisalo, Anna Rosenberg, Sam Asher, Celeste A De Jager Loots, Eija Kekkonen, Kavita Kohtari, Ana Sabsil Lopez Rocha, Gazi Saadmaan, Natalia Soldevila Domenech, Rafael de la Torre Fornell, Tiia Ngandu, Markku Peltonen, Alina Solomon, Miia Kivipelto, Francesca Mangialasche

## Abstract

**Background:** Multidomain lifestyle interventions targeting multiple risk factors have been proposed to reduce cognitive impairment and dementia risk. However, mixed findings hamper their application. More evidence is needed to optimise approaches in different settings. The increasing number of clinical trials being conducted warrants an up-to-date synthesis.

**Methods:** We conducted a systematic review and meta-analysis of randomised controlled trials (RCTs) testing multidomain interventions (three or more components) on cognition or dementia incidence. Maximum-likelihood random-effect models were applied. Sensitivity analyses were conducted to explore source of heterogeneity. Meta-regression analyses were conducted, including by intervention duration and intensity. Risk of bias on cognition was assessed using the revised Cochrane risk-of-bias tool for RCTs (RoB-2). Heterogeneity was estimated using Chi² test, I² statistics, and 95% prediction intervals. GRADE was used for evidence certainty assessment.

**Results:** After screening 4,759 and full-text reading 128 publications, 43 RCTs were eligible and 41 included in the meta-analysis (N=23,209). Risk of bias was generally “low”, with most concerns in older studies. Small but statistically significant intervention benefits were found for global cognition (composite score of validated neuropsychological tests; SMD=0·28; 95% CI: 0·10 to 0·45), and most of the other cognitive measures. High heterogeneity was observed for global cognition composite scores and could be only partially explained by three smaller RCTs. Intervention effect-size was significantly associated with shorter duration (P-value=0.009) and higher observed intensity (P-value=0.008).

**Interpretation:** Multidomain interventions have small but consistent beneficial effects on cognitive measures, suggesting the potential to reduce cognitive impairment and dementia risk. High heterogeneity across RCTs can hinder data pooling. More evidence on longer-term effect is needed. Future research should prioritise harmonisation of methodologies and reporting, long-term extended follow-up data, clinically relevant dementia-risk surrogate outcomes, and evidence from more diverse cultural, geographical, and socio-economic contexts.

## INTRODUCTION

Risk-reduction and prevention are crucial to address the dementia public health challenge, with an excess of 152 million cases expected globally by 2050.^1^ The Lancet Commission on Dementia identified 14 modifiable risk factors accounting for approximately 45% of all dementia cases, providing opportunity for prevention.^2^ Several of these factors (e.g., physical inactivity, obesity, hypertension, high LDL-cholesterol, diabetes) are associated to lifestyle and cardiometabolic health, suggesting that adoption of healthy lifestyles and optimal management of cardiometabolic disorders could help reduce dementia risk.

Dementia has a multifactorial, heterogeneous etiopathogenesis. Multidomain lifestyle interventions, addressing multiple risk factors and disease mechanisms at the same time through health and behavioral changes, have thus emerged as promising and-cost effective preventive approaches.^3–6^ Consistently, the World Health Organization (WHO) Guidelines for the Risk Reduction of Dementia and Cognitive Decline,^7^ mainly based on intervention studies evidence, highlighted multidomain approaches as a promising strategy.

Multidomain randomized controlled trials (RCTs) usually address modifiable factors such as exercise; diet; cardiometabolic management; cognitive and social stimulation.^8^ The adaptability of multidomain interventions to the individual needs and lifestyles might support uptake and efficacy.^9^ Although their biological underpinnings are not yet fully understood, they might act via multiple mechanisms, like improving cardiometabolic health, enhancing cognitive reserve, and reducing inflammation.^8^ Despite the increasing number of multidomain RCTs conducted in the last 15 years, the evidence of their efficacy on cognitive outcomes is still mixed, with some RCTs showing benefits and others reporting modest or no effects.^10,11^ This could be due to high variability in study design, regarding, e.g., selection of the target population and intervention structure, which can affect adherence levels and overall intervention response. Previous systematic reviews and meta-analyses on the efficacy of multidomain interventions for dementia prevention and risk-reduction indicated small or modest cognitive benefits.^10,11^ As many multidomain RCTs have been initiated and completed since,^12–15^ a review of more recent evidence could provide novel insights on the efficacy of these interventions and the factors that are likely to determine their success. Additionally, evidence is still very limited from low- and middle-income countries (LMICs), as well as lower socio-economic sections of high income countries society, where the highest increase of dementia cases is expected by 2050, and prevention should, thus, be a high priority.^1^

To address these gaps, we conducted a systematic review and meta-analysis of the currently available evidence on the efficacy of multidomain interventions in adults with normal cognition or mild cognitive impairment (MCI), on outcomes related to dementia and cognitive function.

## METHODS

This study was conducted in connection with the update of the WHO Guidelines for the Risk Reduction of Dementia and Cognitive Decline,^7^ following the WHO methodology for evidence synthesis.^16^ The protocol, including the review question, was preregistered (PROSPERO; ID: CRD420251018238). Results are reported according to the Preferred Reporting Items for Systematic Reviews and Meta-analyses (PRISMA).^17^

### Eligibility criteria

Only RCT were included in this review, as the gold standard for intervention studies. Following the PICO principles, the inclusion criteria included (P)Population: Adults with normal cognition or MCI or dementia-free prodromal Alzheimer’s disease (AD); (I)Intervention: Multidomain interventions, addressing three or more risk factors, with at-least a lifestyle component; (C)Comparator: Usual care, any other type of lower-intensity/less-tailored interventions, or general health advice/education; (O)Outcomes: Cognitive function, Incident MCI, or Incident Dementia. Detailed eligibility criteria are reported in Supplementary Methods.

### Literature Search and Study Selection

On the 2^nd^ of May 2025, Medline via OVID; EMBASE via Elsevier; The Cochrane Library via Wiley; ClinicalTrials.gov; and the WHO International Clinical Trials Registry Platform were searched for RCTs or systematic reviews (SRs) of RCTs published from the 1st of January 2010 (based on the time of publication of the first multidomain RCTs in the context of dementia prevention and by adding the previous five years for broader coverage)^9^ until the date of search. Detailed search terms and strategies are presented in Supplementary Methods.

### Pre-defined outcomes and outcome measures

Pre-defined outcomes were cognitive function; incident MCI; incident Dementia. Multiple cognitive function outcome measures were included based on data availability, when reported by at least two RCTs. The main cognitive function measure was global cognition measured as composite score of validated neuropsychological tests. Other cognitive function measures are described in Supplementary Methods.

### Data Extraction and Risk of Bias

Detailed data from the eligible RCTs were independently extracted by at least two researchers (SA, EK, JL, ASRL, AR, GS), and discrepancies resolved through discussion with a third researcher (MB, JL, AR). Extracted data included publication identifiers; RCT design; participants characteristics; intervention; comparator; outcome measures; intervention effect; statistical methods.

The risk of bias was independently assessed by two researchers (MB, RS) using the revised Cochrane risk-of-bias tool for randomized trials (Cochrane RoB-2)^18^ on the cognitive function outcome. Full details are provided in Supplementary Methods. Publication bias was assessed with Egger’s test and visual inspection of funnel plots.

### Data Synthesis

Data analyses were performed using STATA19 (StataCorp., College-Station, TX). Effect-size for continuous outcomes was calculated as standardized mean difference (SMD) between intervention and control groups mean change from baseline to the end of the intervention period, with standard error of mean difference. A random-effect meta-analysis was fitted using maximum likelihood estimation of between-study variance (τ²) to calculate weighted effect-sizes and confidence intervals (CIs). Threshold significance was set, for all analyses, at P-value <0.05. Heterogeneity was estimated using Cochrane Chi² test (with a Chi² P = 0.1 indicating significant heterogeneity); I² statistics; and 95% prediction intervals (PIs).

To investigate potential causes of heterogeneity on the main outcome measure for cognitive function (i.e., global cognition measured as composite score of validated neuropsychological tests), two sensitivity analyses were conducted, first excluding each RCT in a stepwise manner, one at the time; and excluding smaller-size studies, defined as N<100. To investigate factors affecting intervention efficacy, univariate meta-regression analyses were also conducted on global cognition (composite score) by intervention duration (months); per-protocol intervention intensity (ratio between total intervention sessions planned and intervention duration^19^, Supplementary Table 1); observed intervention intensity (ratio between adherence-adjusted attended intervention sessions and intervention duration, Supplementary Methods and Supplementary Table 2); selection of target populations (number of modifiable risk factors used both as intervention targets and eligibility criteria); or target population cognitive status (healthy at-risk vs MCI/Prodromal AD). Corresponding subgroup analyses were conducted with pre-defined cut-offs for interventions lasting <12 and ≥12 months; per-protocol intensity of <nine and ≥nine (based on previous systematic review^19^); RCTs with <three or ≥ three risk factors used both as intervention targets and eligibility criteria (Supplementary Table 3); or target population healthy at-risk vs MCI/Prodromal AD. To investigate the impact of risk of bias, the meta-analysis on the main cognitive function outcome measure, meta-regressions, and subgroup analyses were conducted including only studies with low risk of bias or minimal concern (i.e., some concern in only one domain).

Evidence certainty was assessed for cognitive function outcome measures based on the Grading of Recommendations Assessment, Development and Evaluation (GRADE; http://gradepro.org) by two researchers (MB, RS; Supplementary Methods).

This systematic review and meta-analysis was completed in April 2026. 9; Additional methodological aspects not included in the protocol registration are provided in Supplementary Methods.

## RESULTS

### Study selection and risk of bias

After duplicate removal, 4,759 articles were screened and 128 assessed at full text level (Figure 1). Of these, 43 were found eligible (Figure 1, Supplementary Table4) and 85 excluded (Supplementary Table 5). Detailed description of eligible RCTs is provided in Table 1, Supplementary Table 6, and Online Material. Risk of bias was generally “low”, with most concerns in older studies (detailed results in Supplementary Results and Supplementary Figure1).

**Figure 1.**
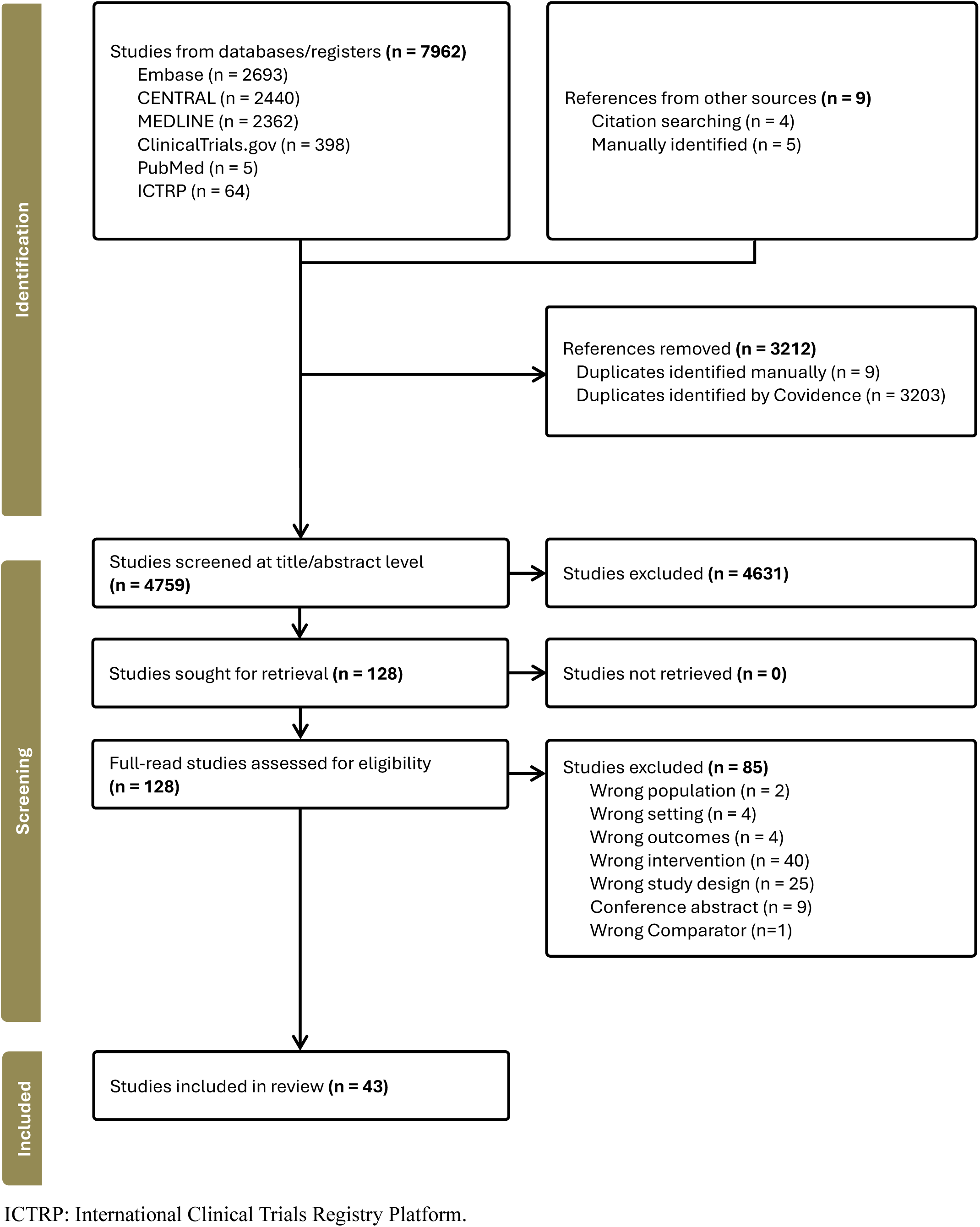
PRISMA Flowchart

**Table 1.**
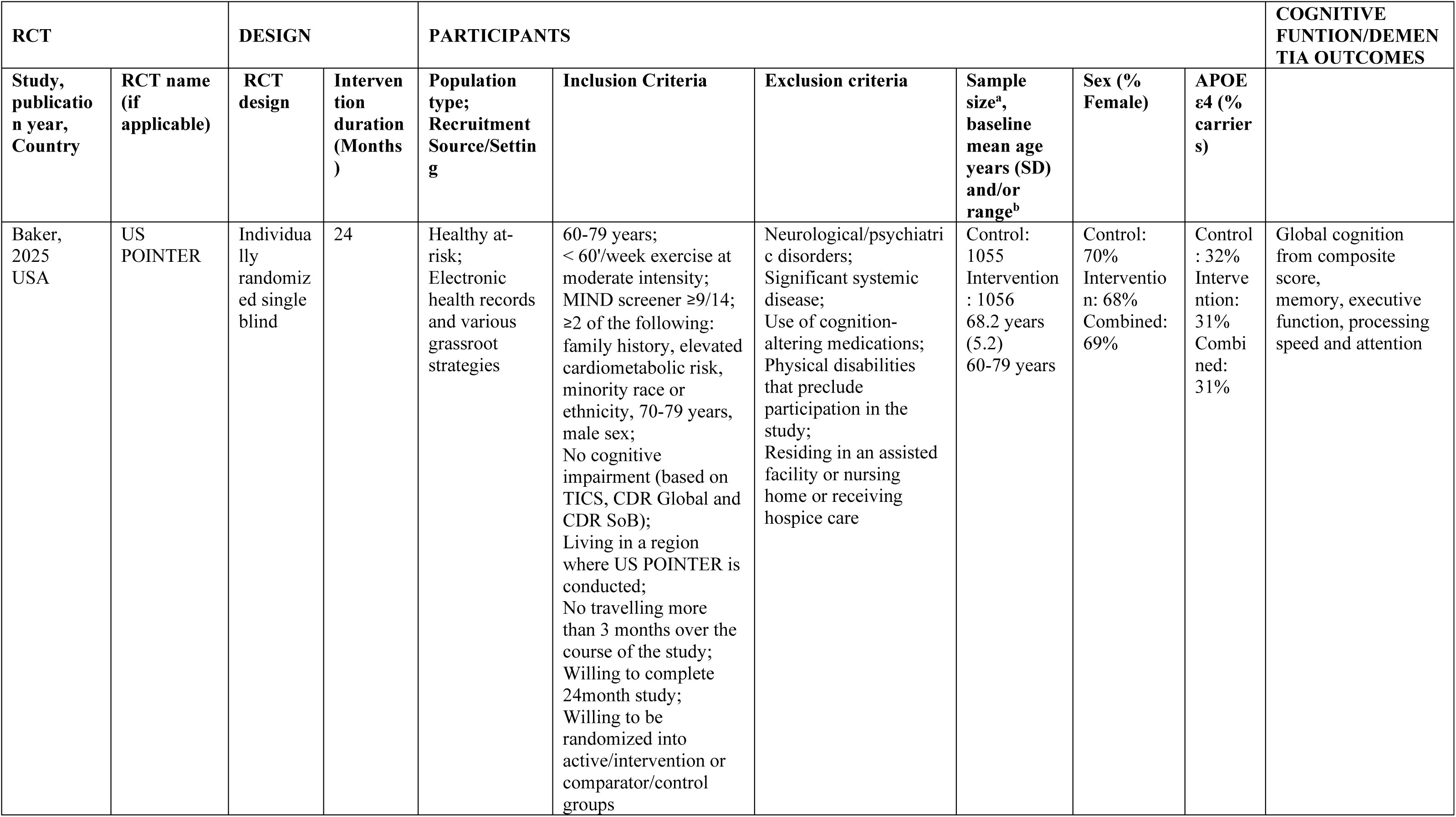
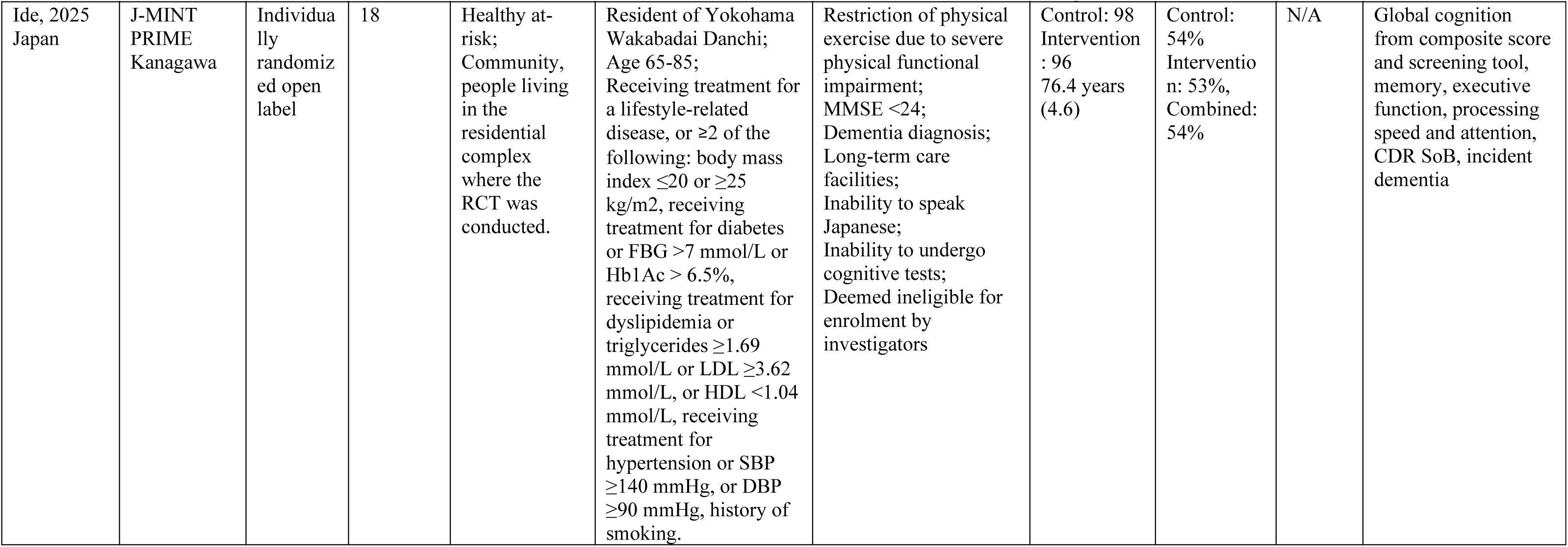
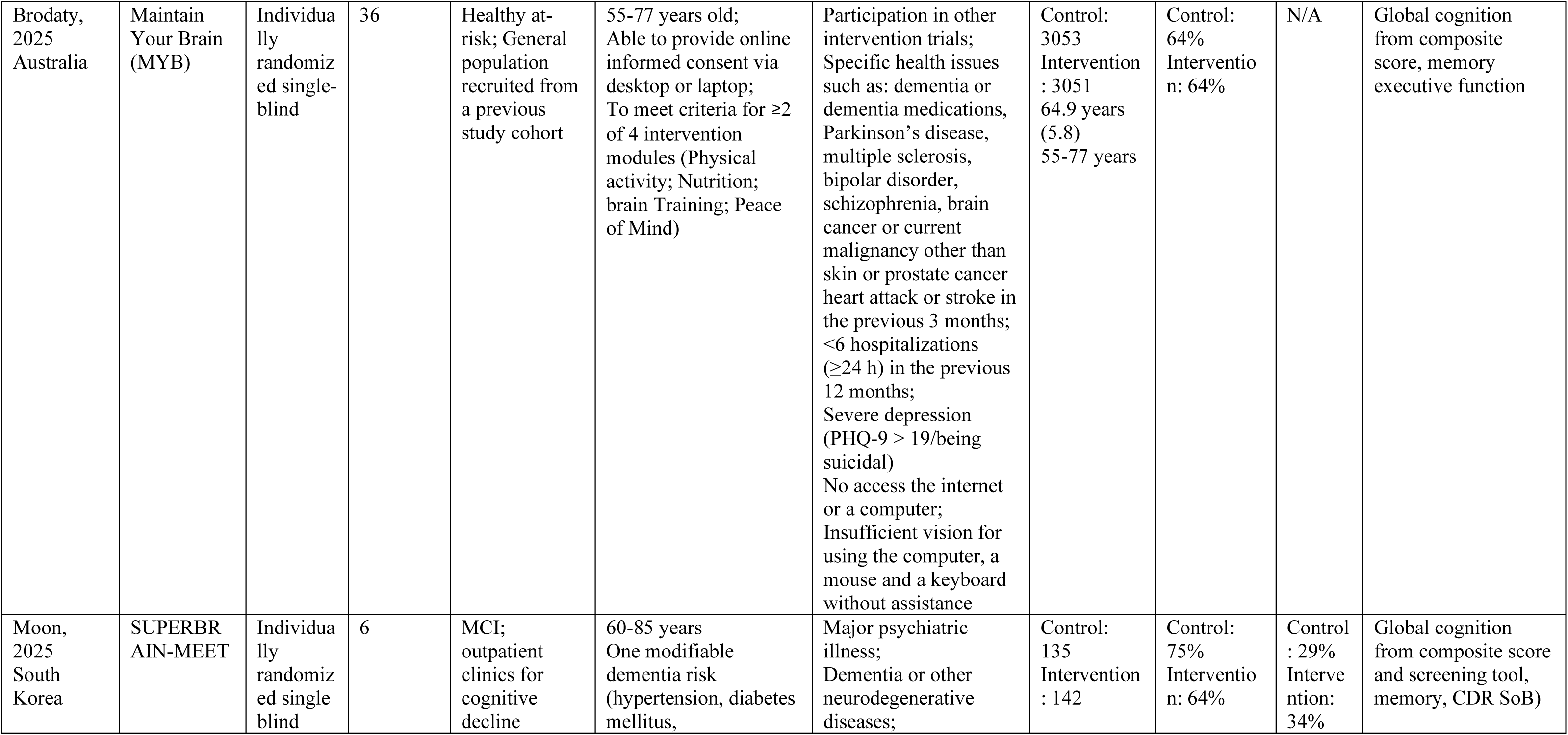
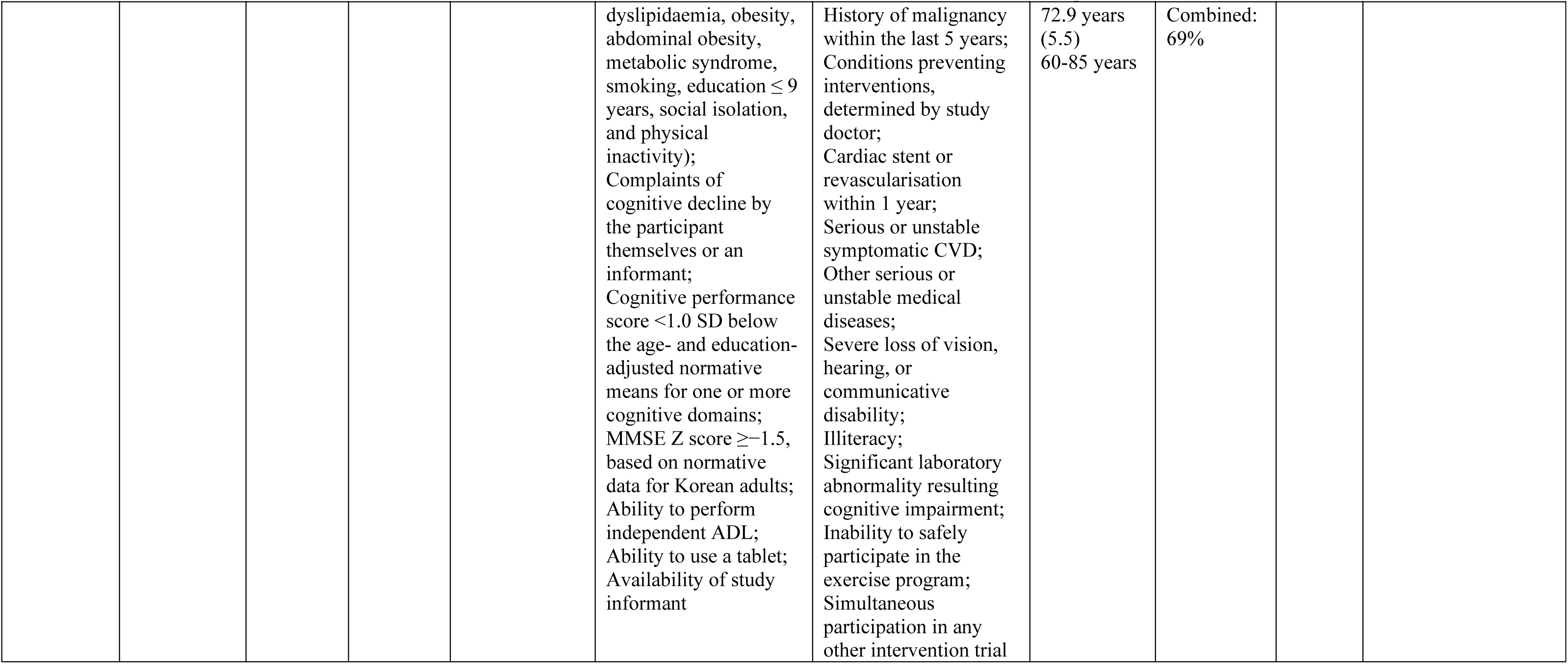
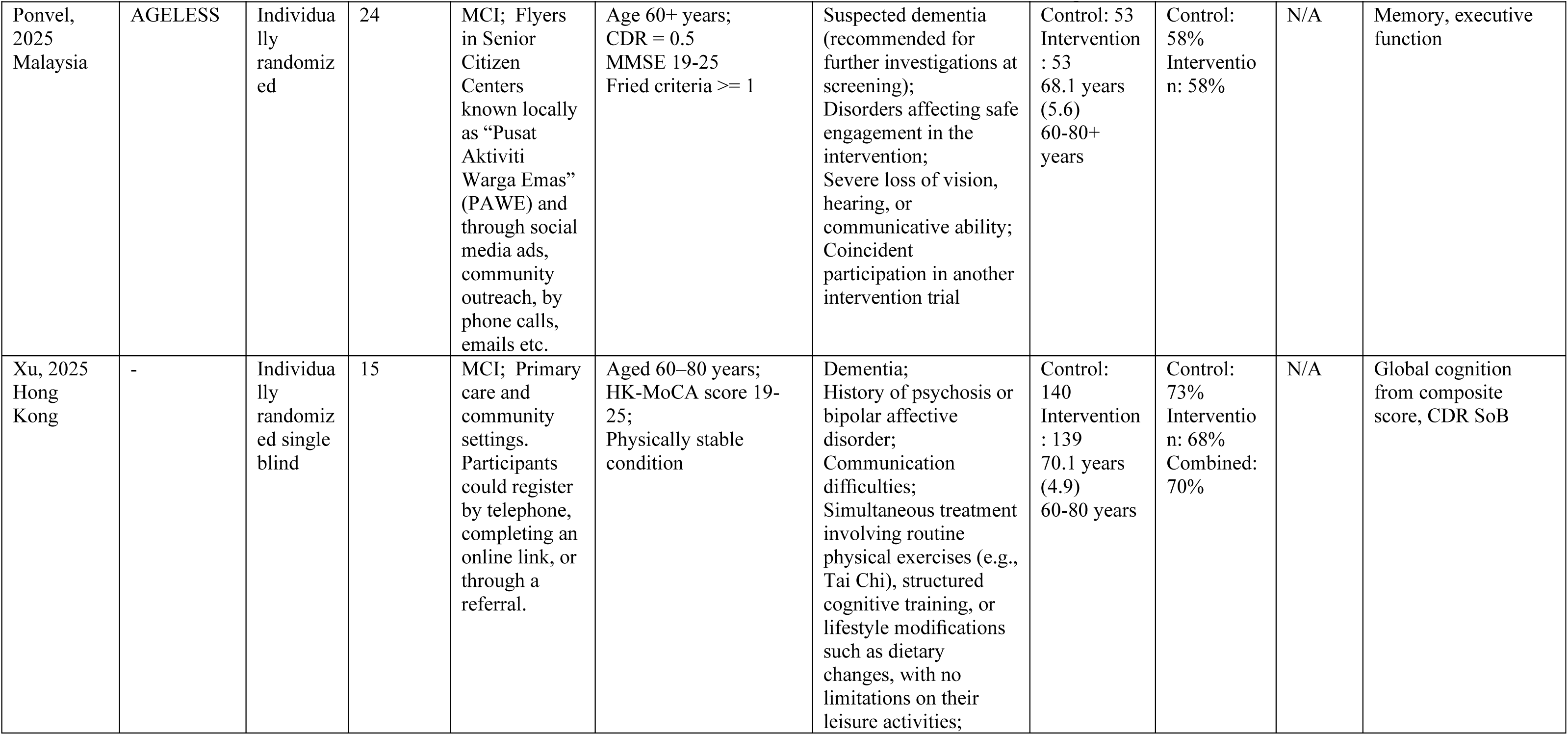
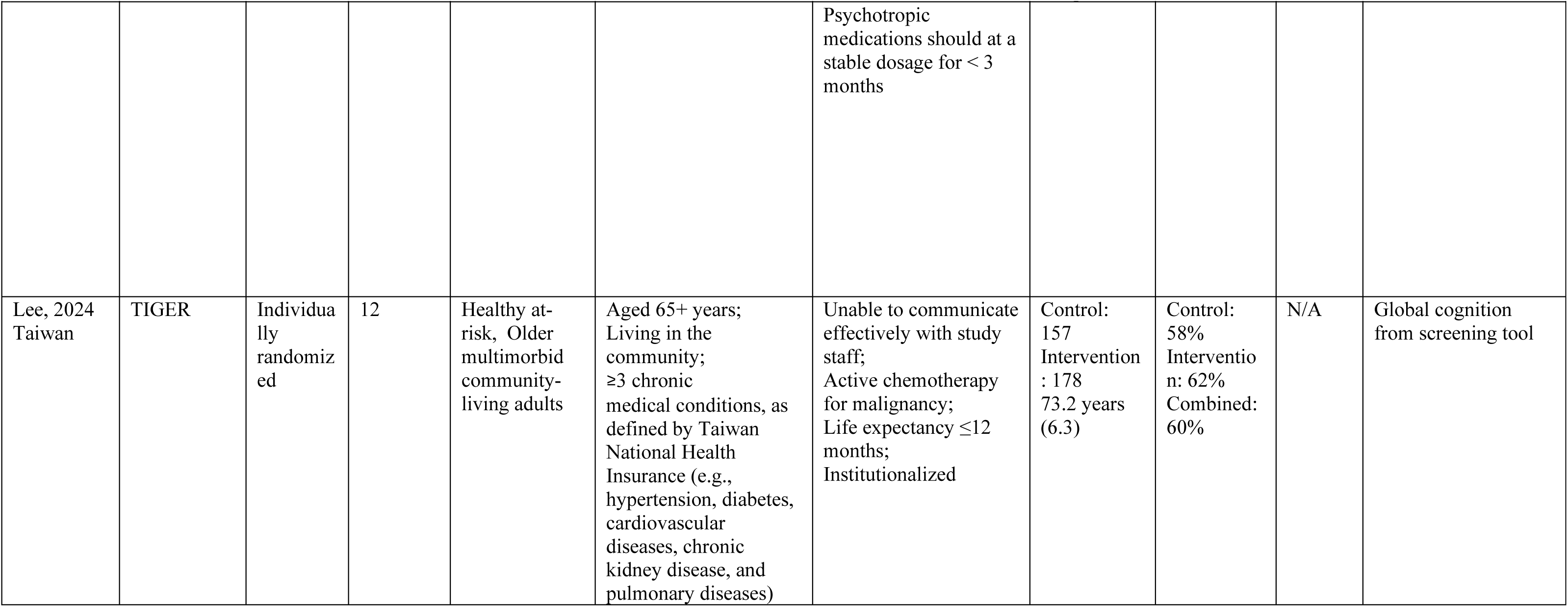
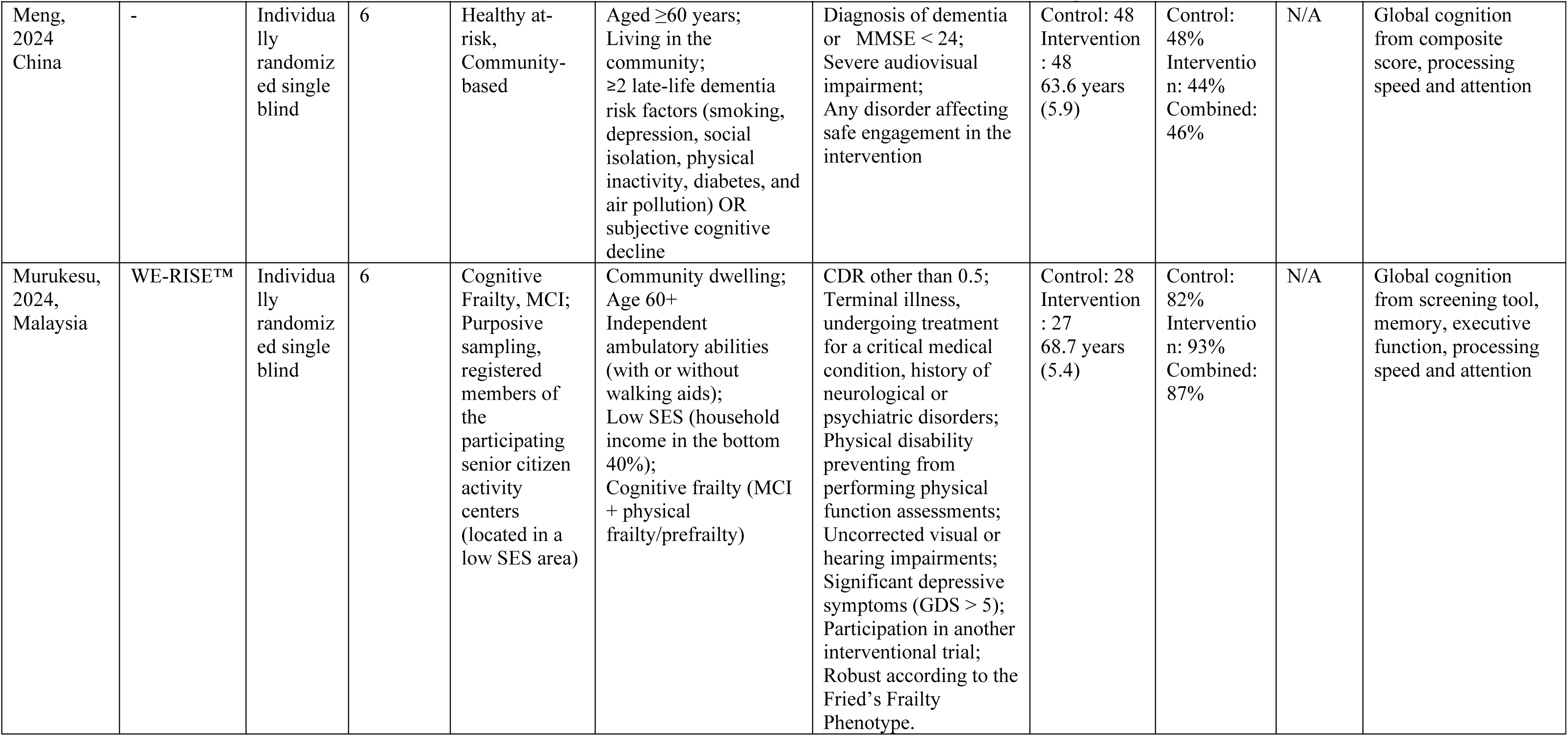
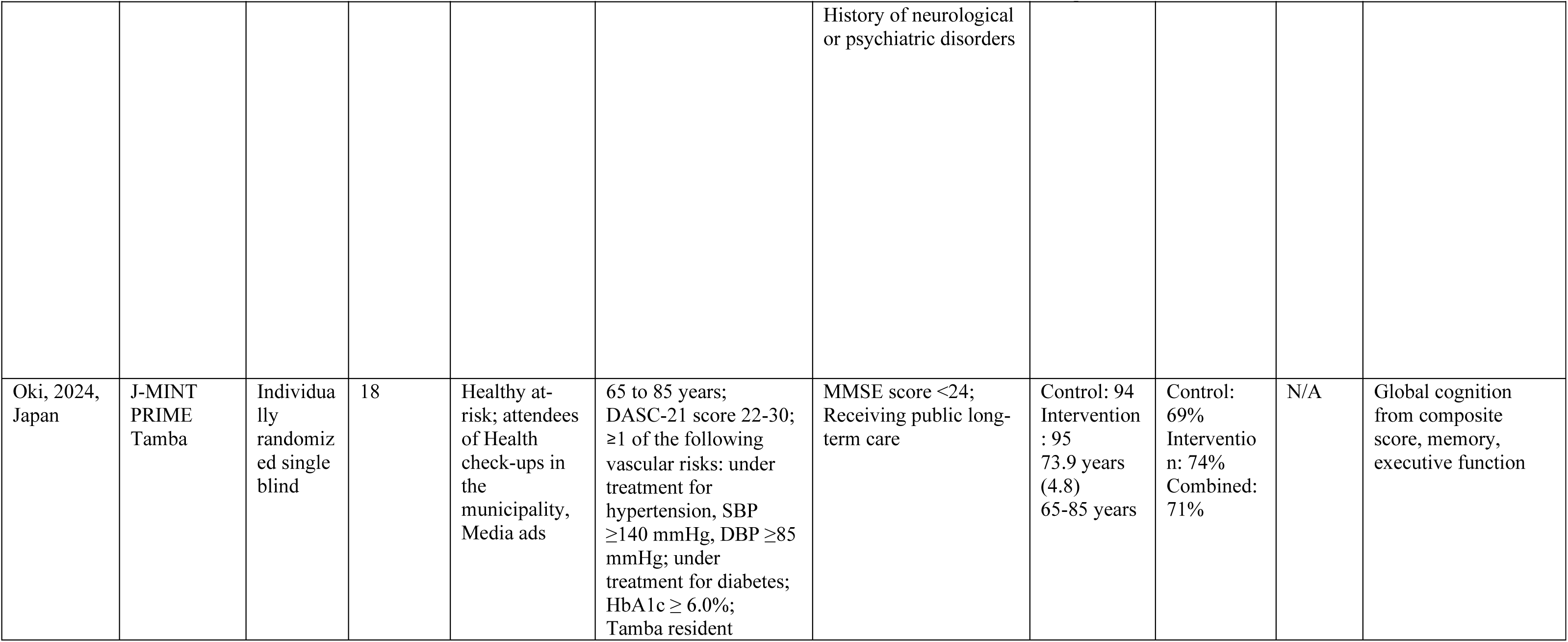
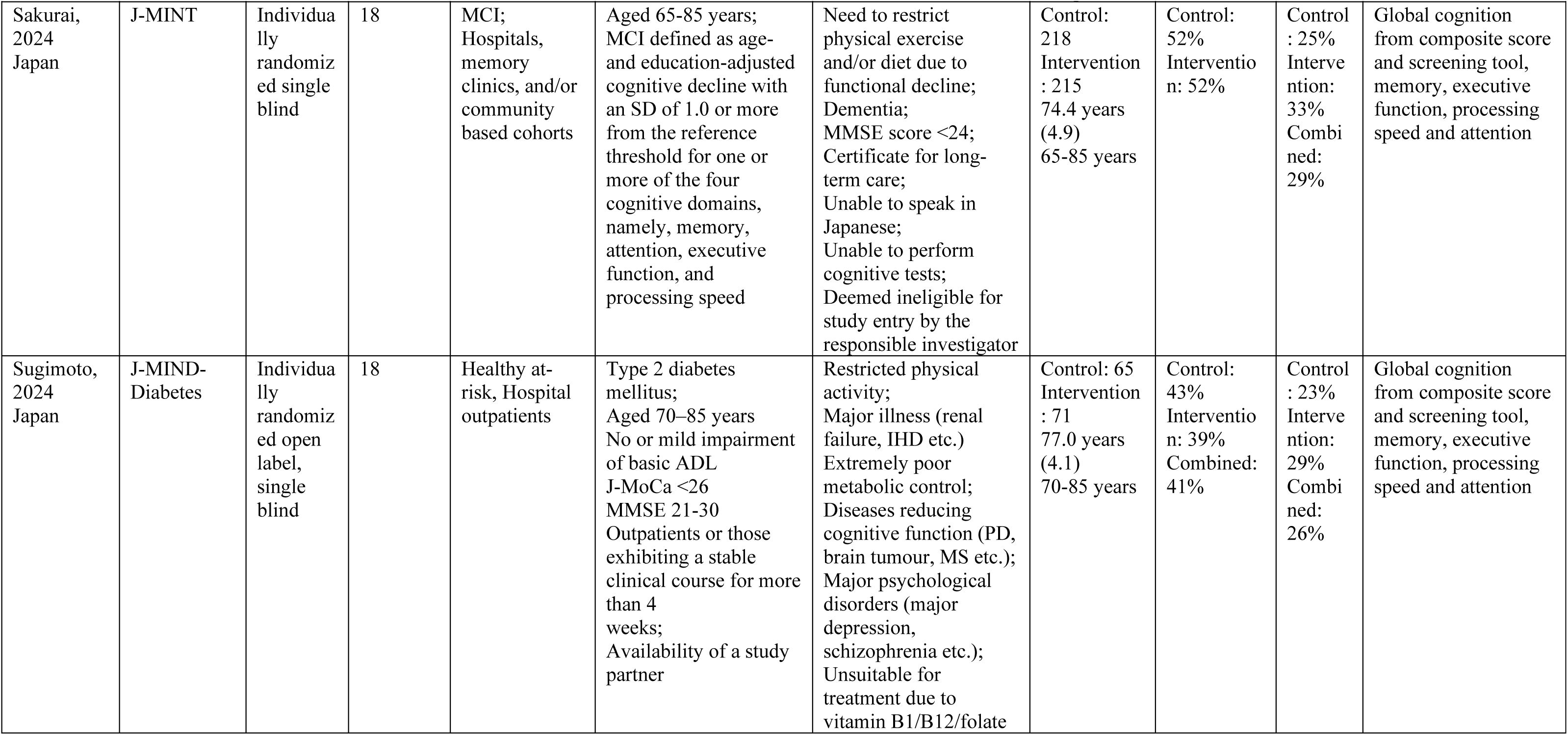
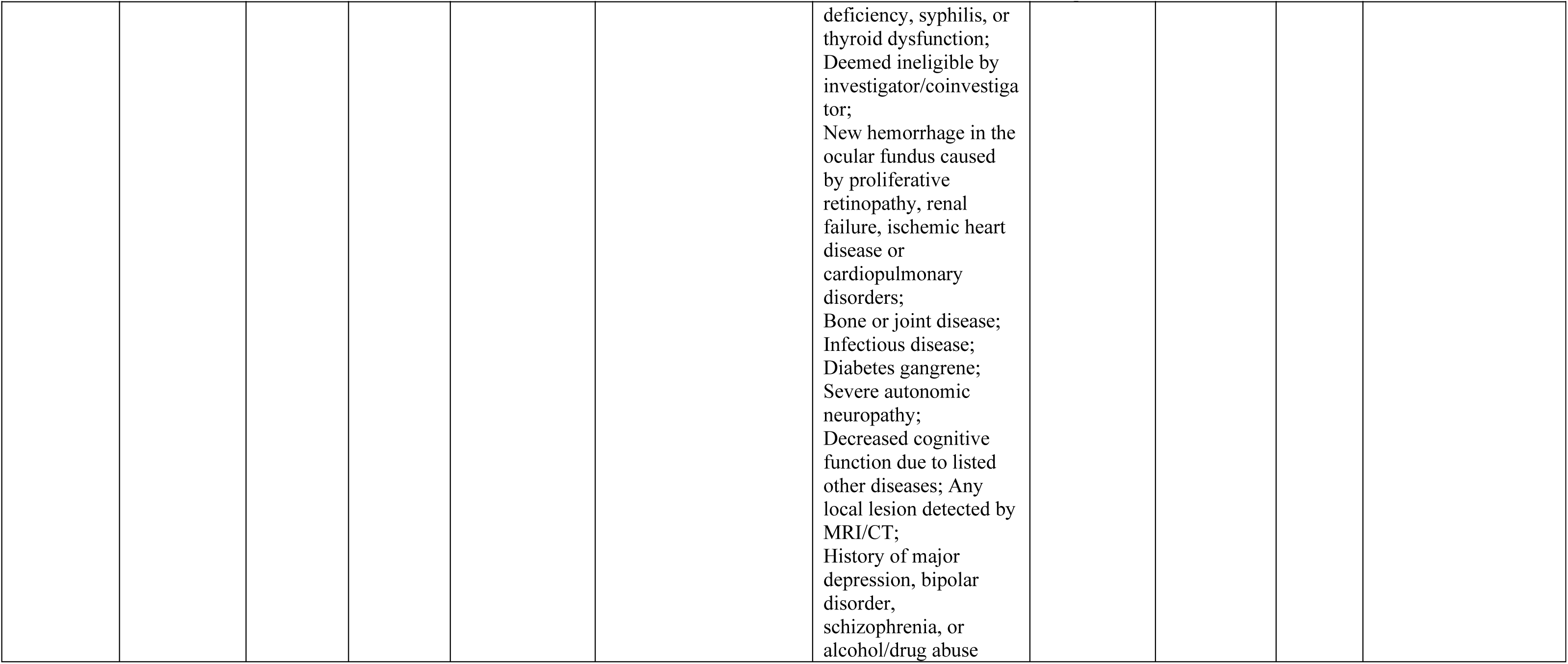
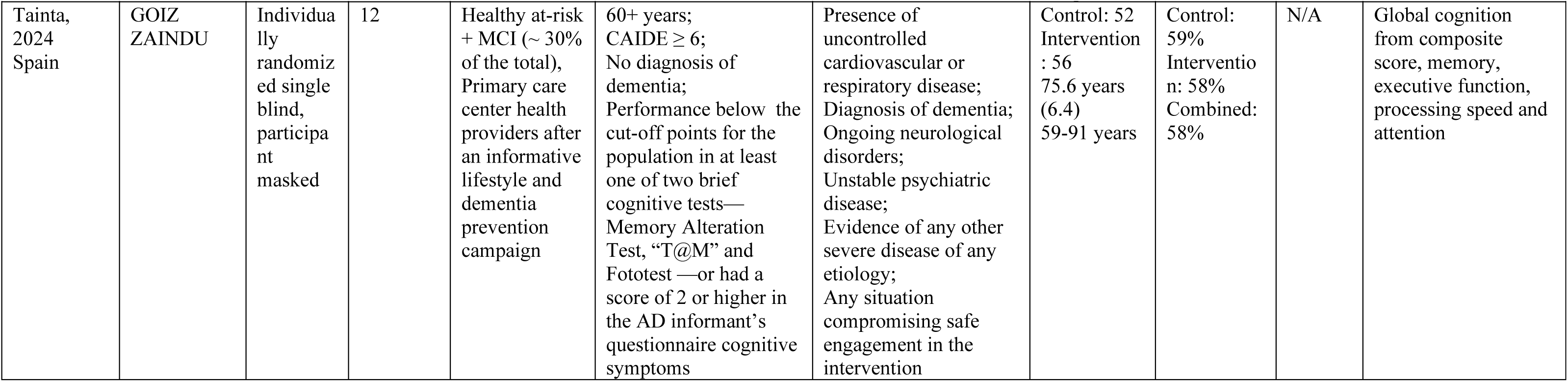
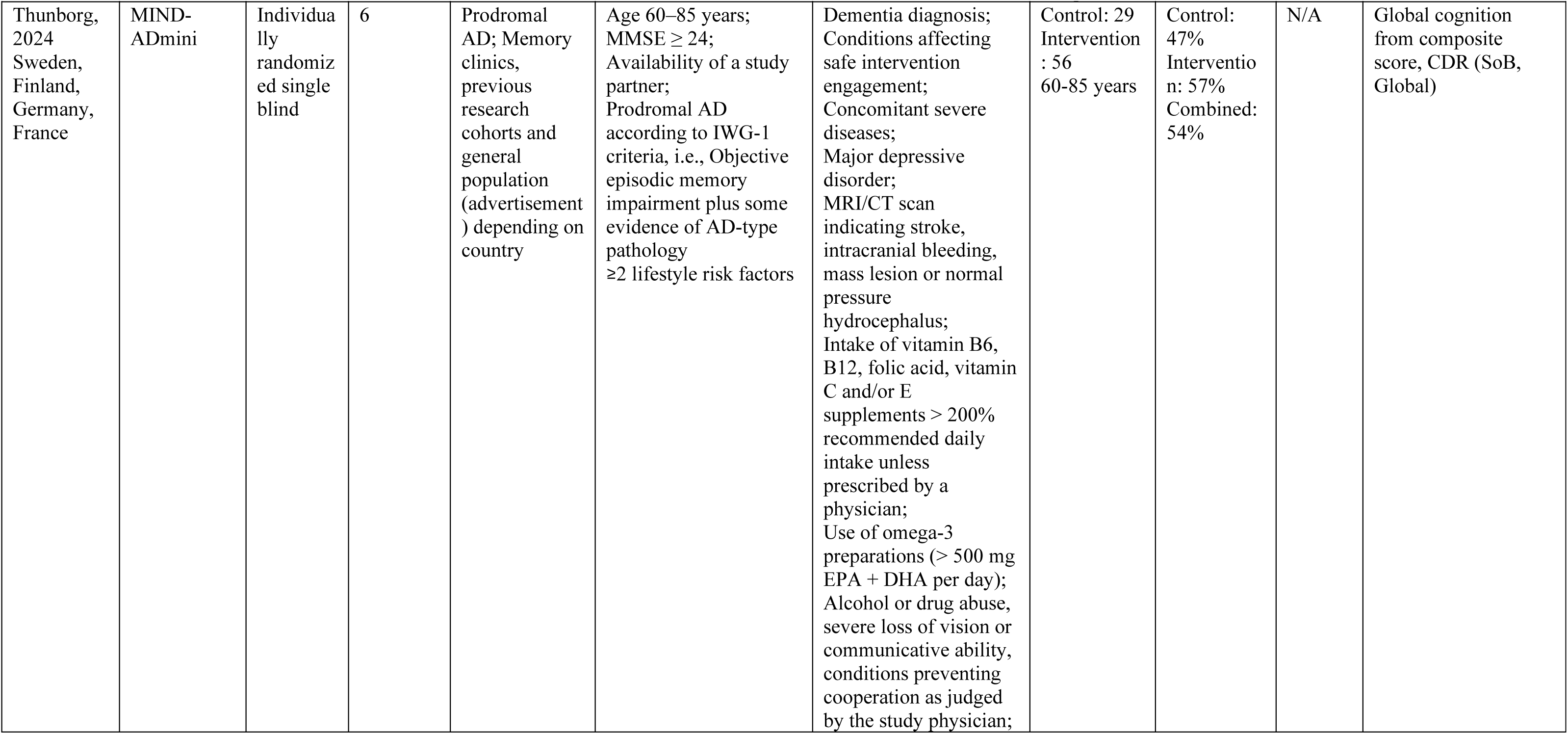
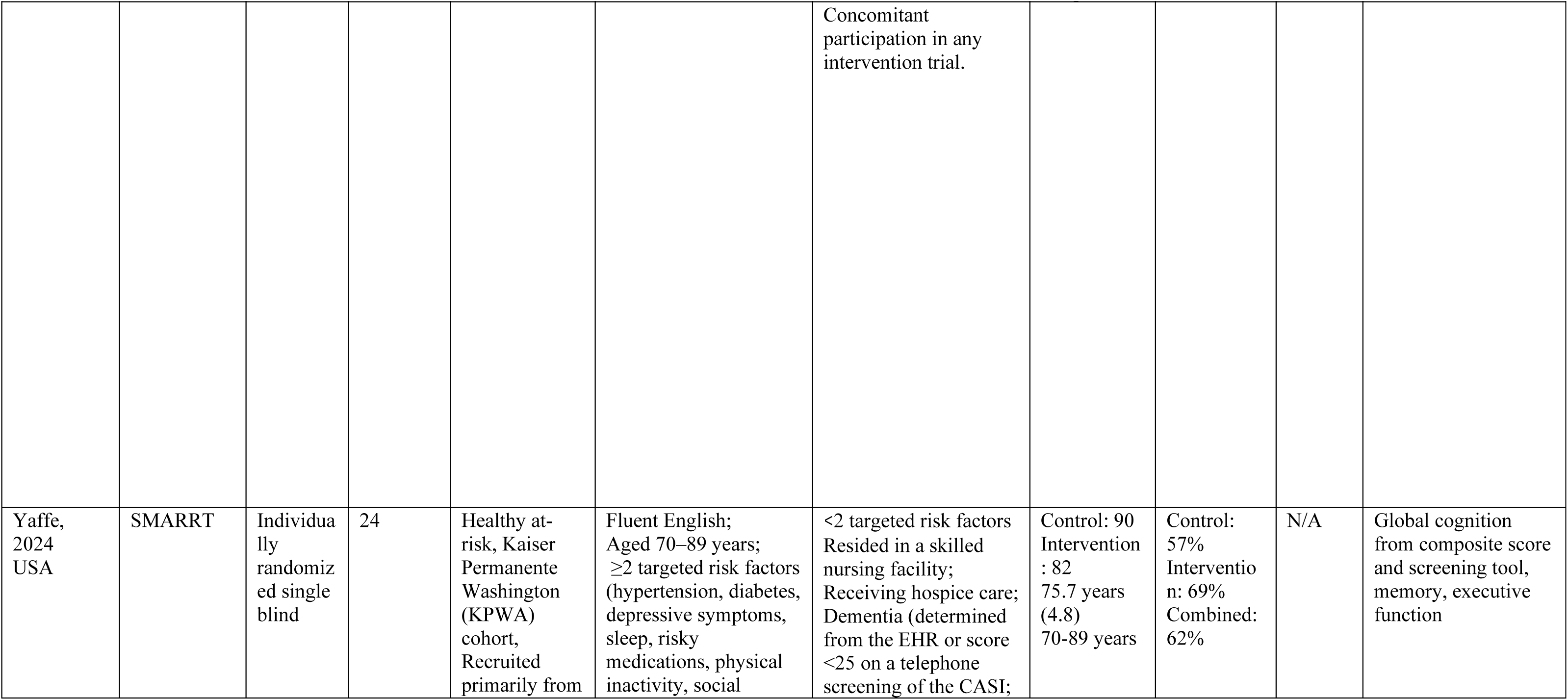
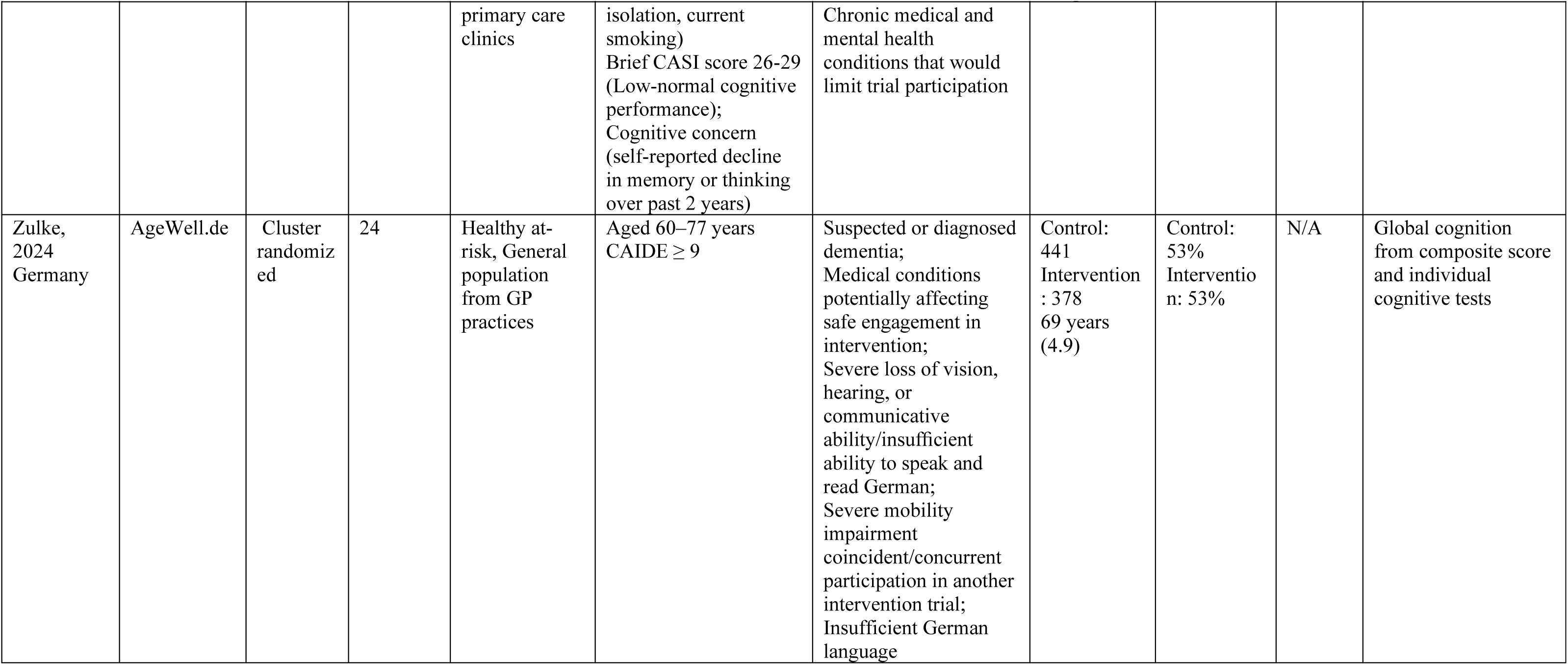
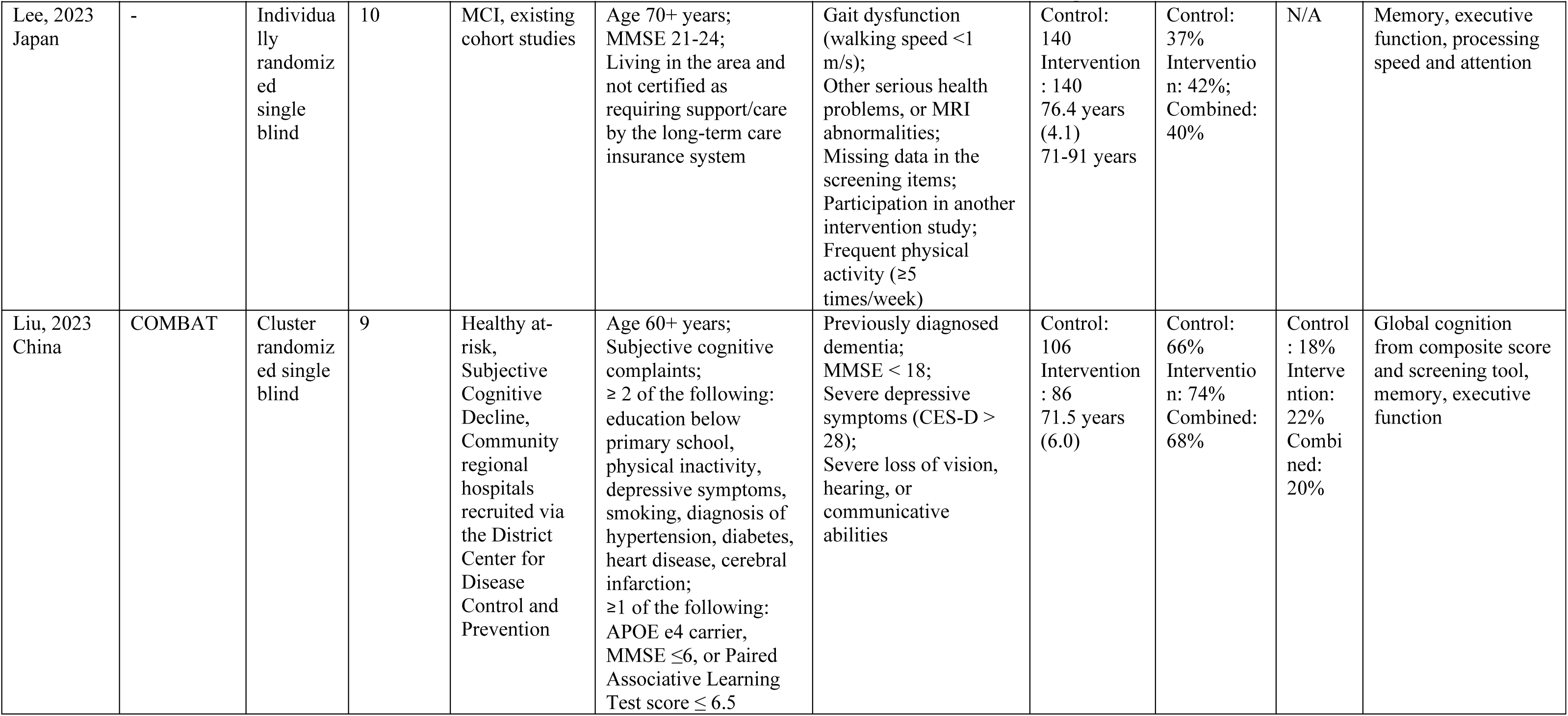
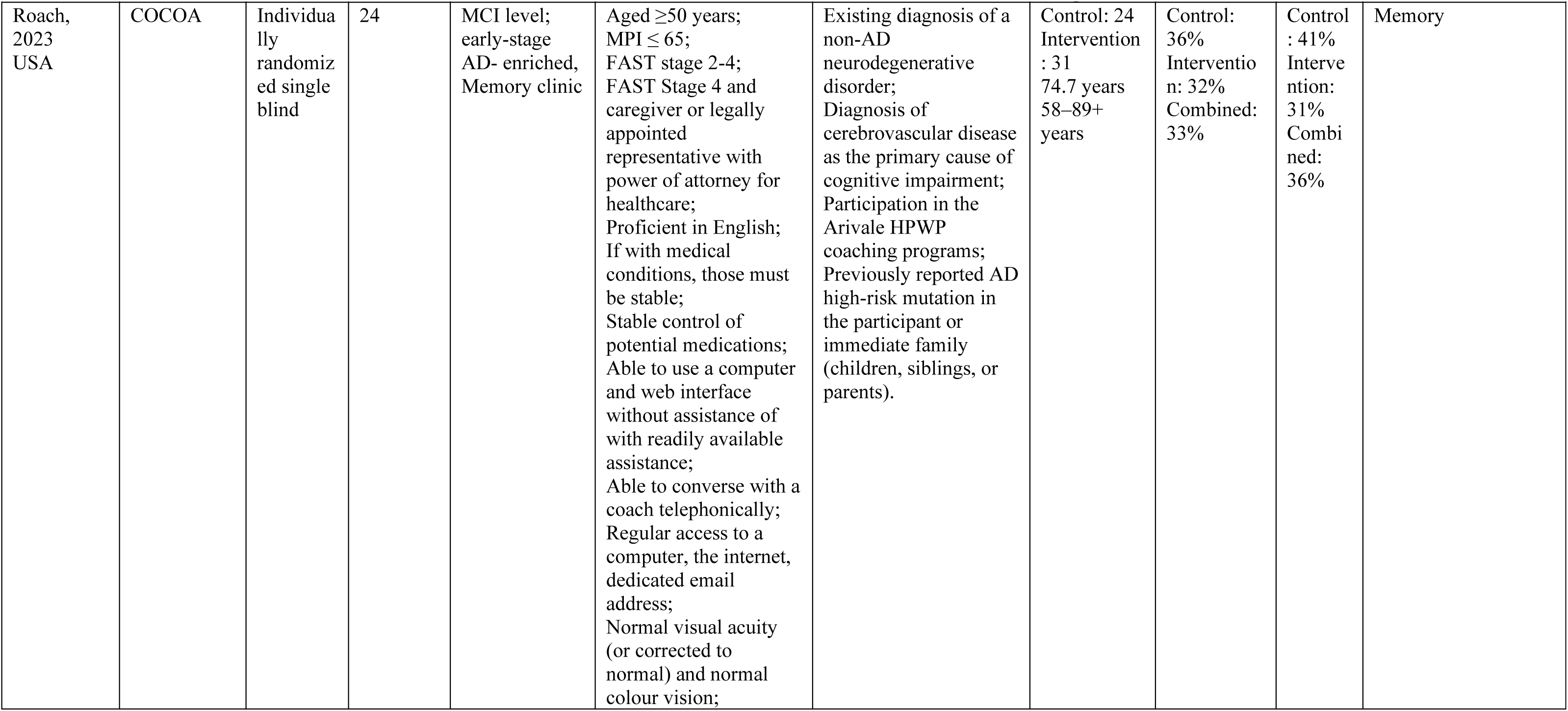
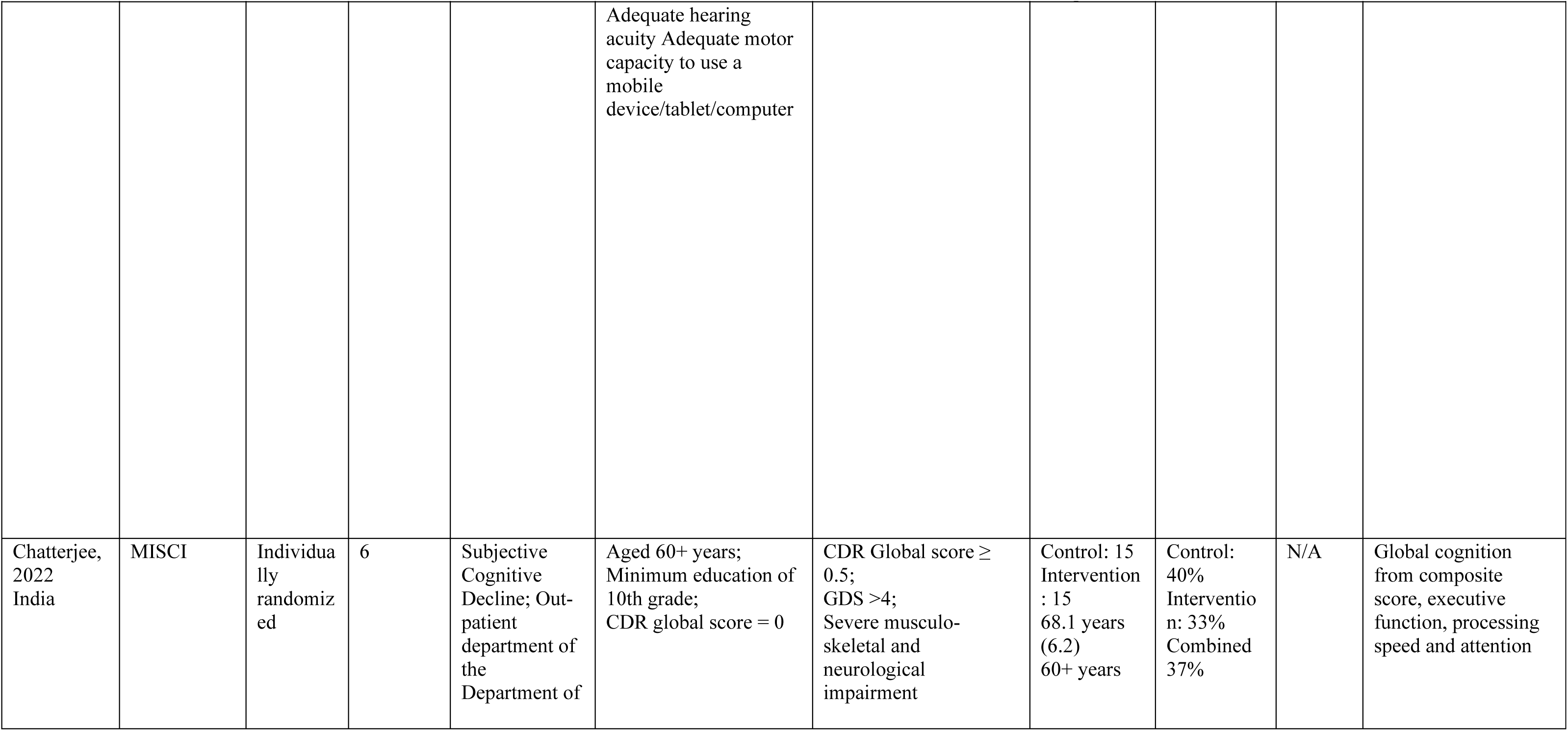
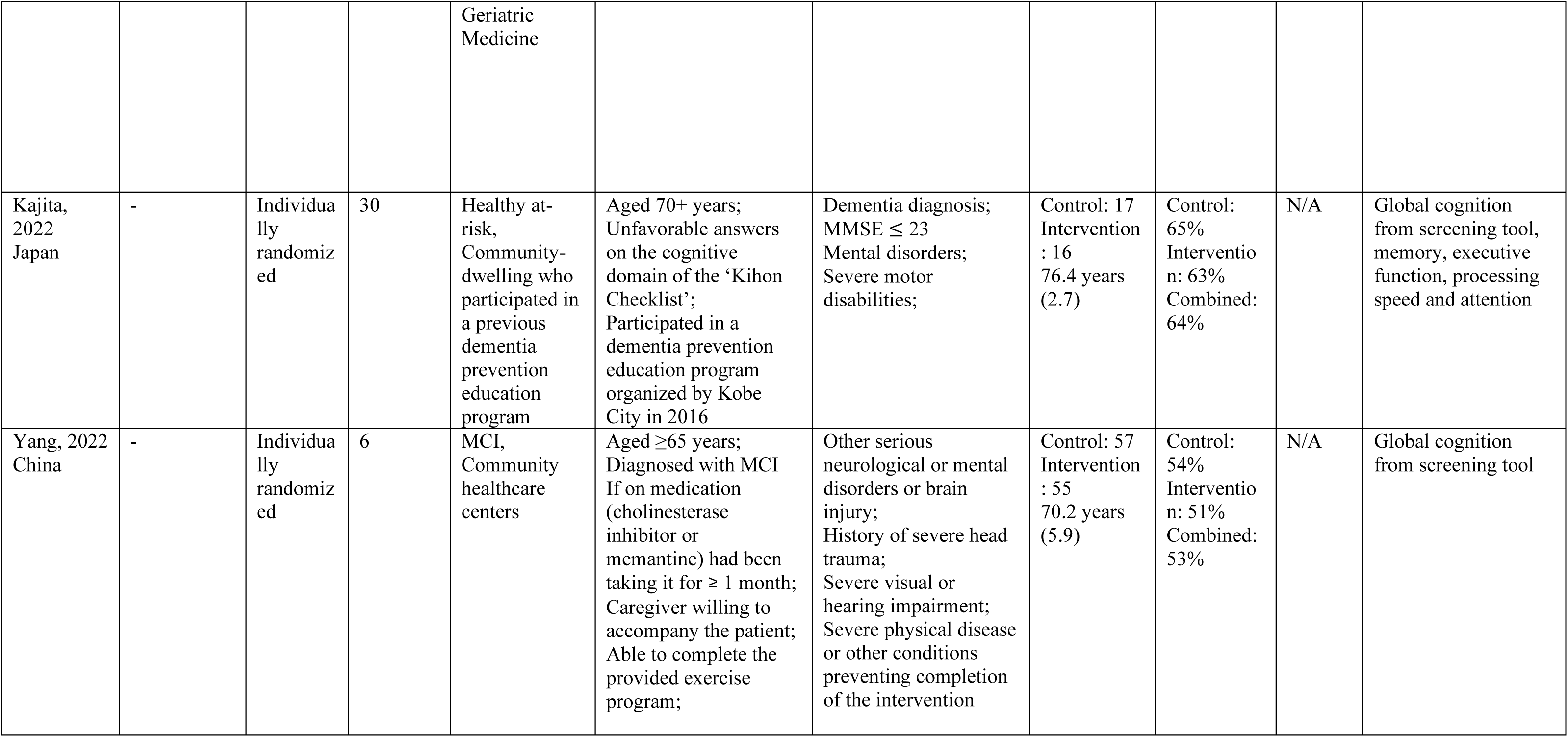
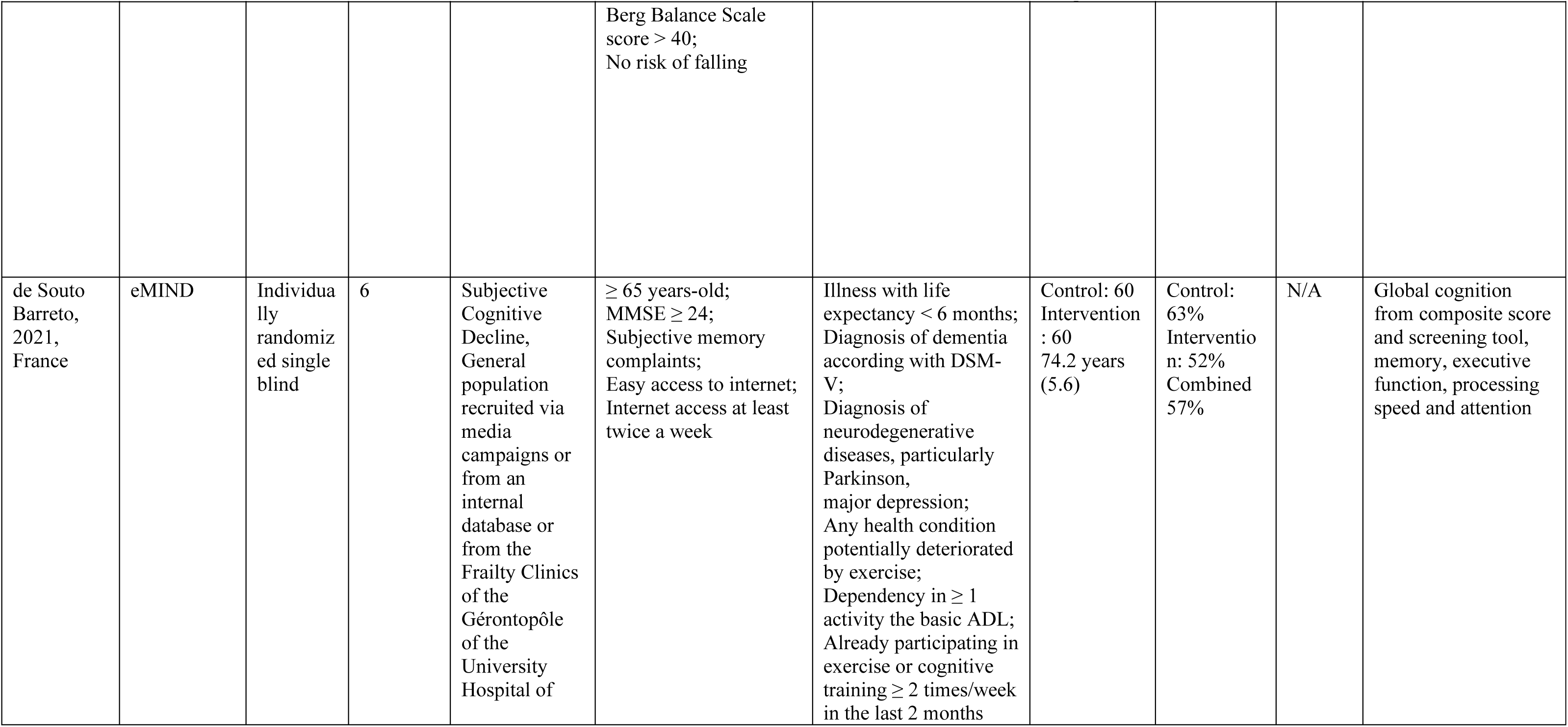
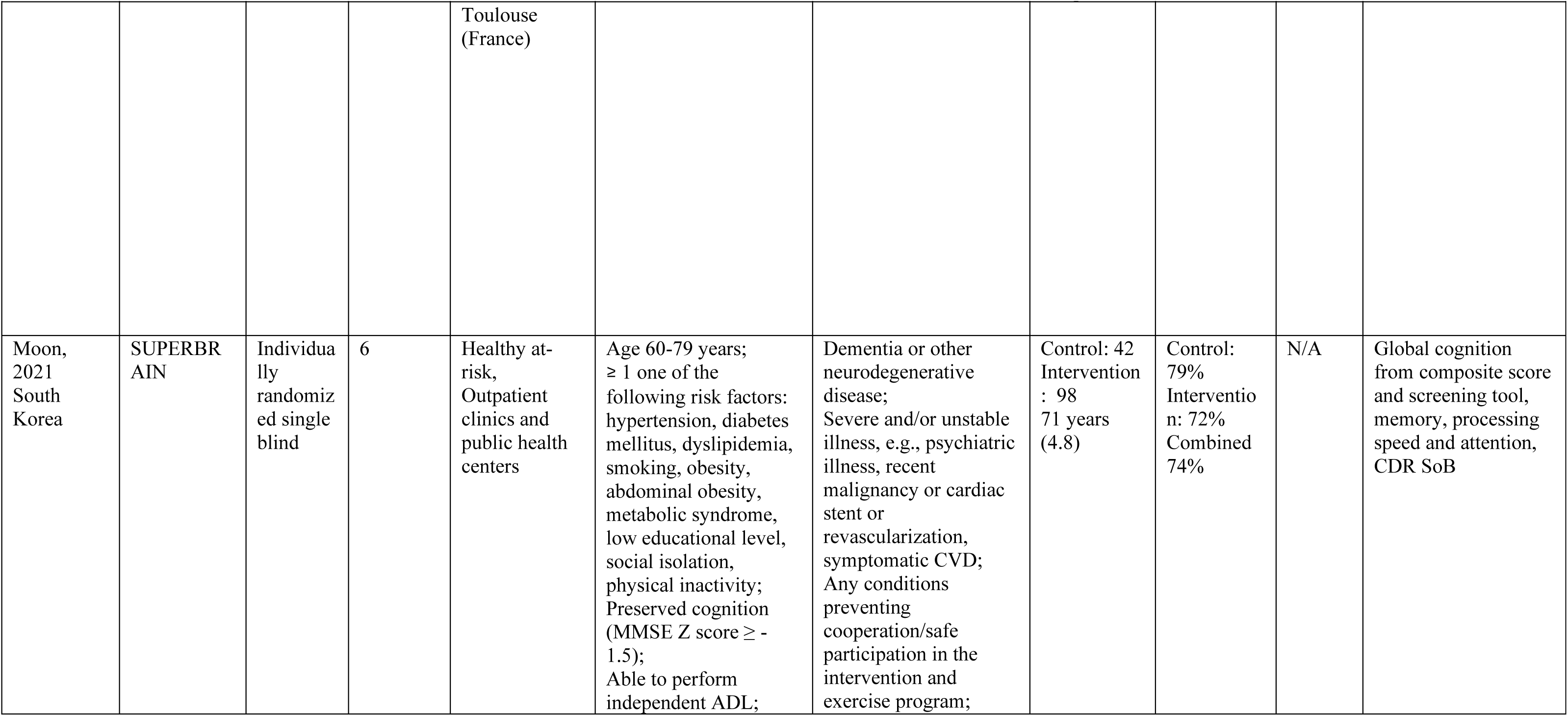
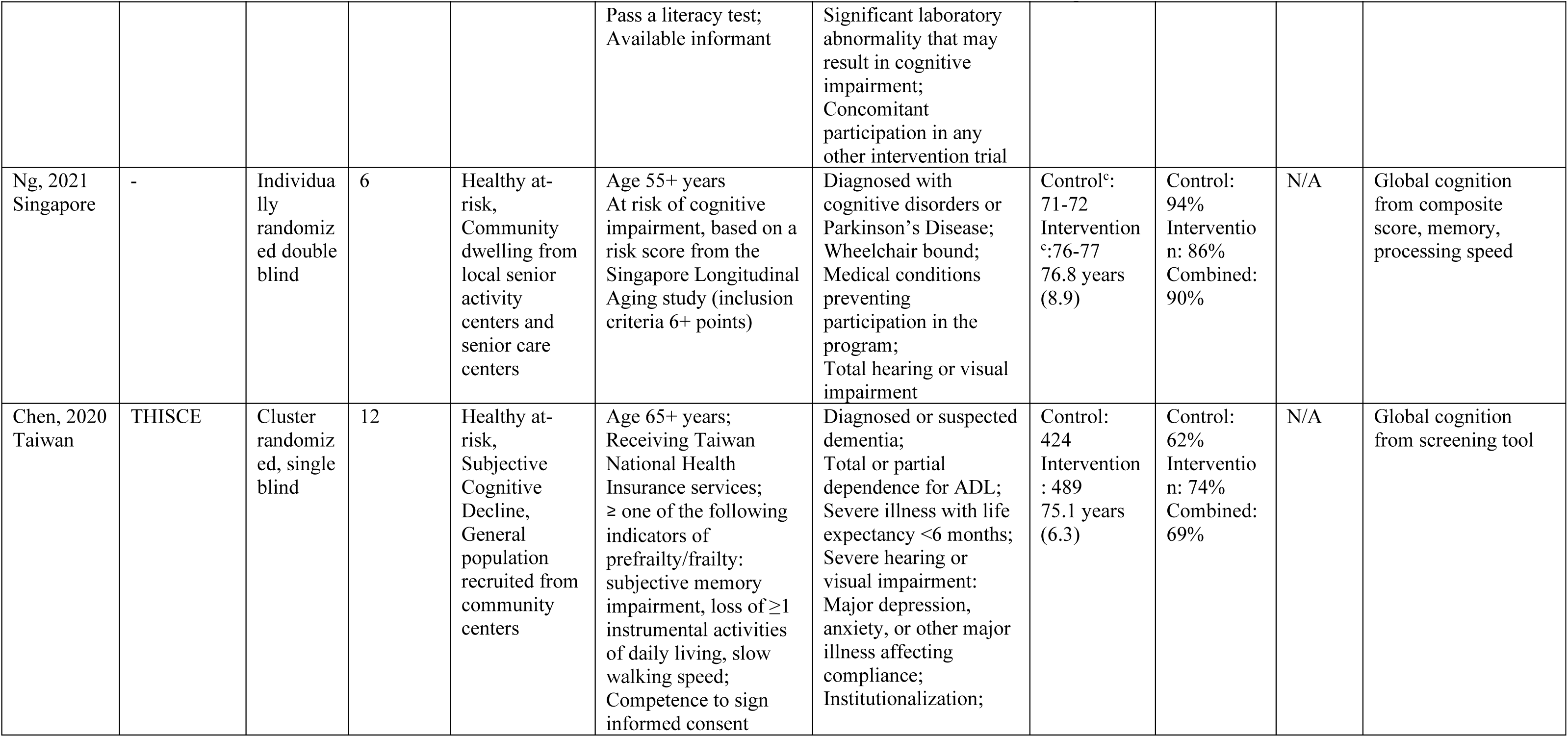
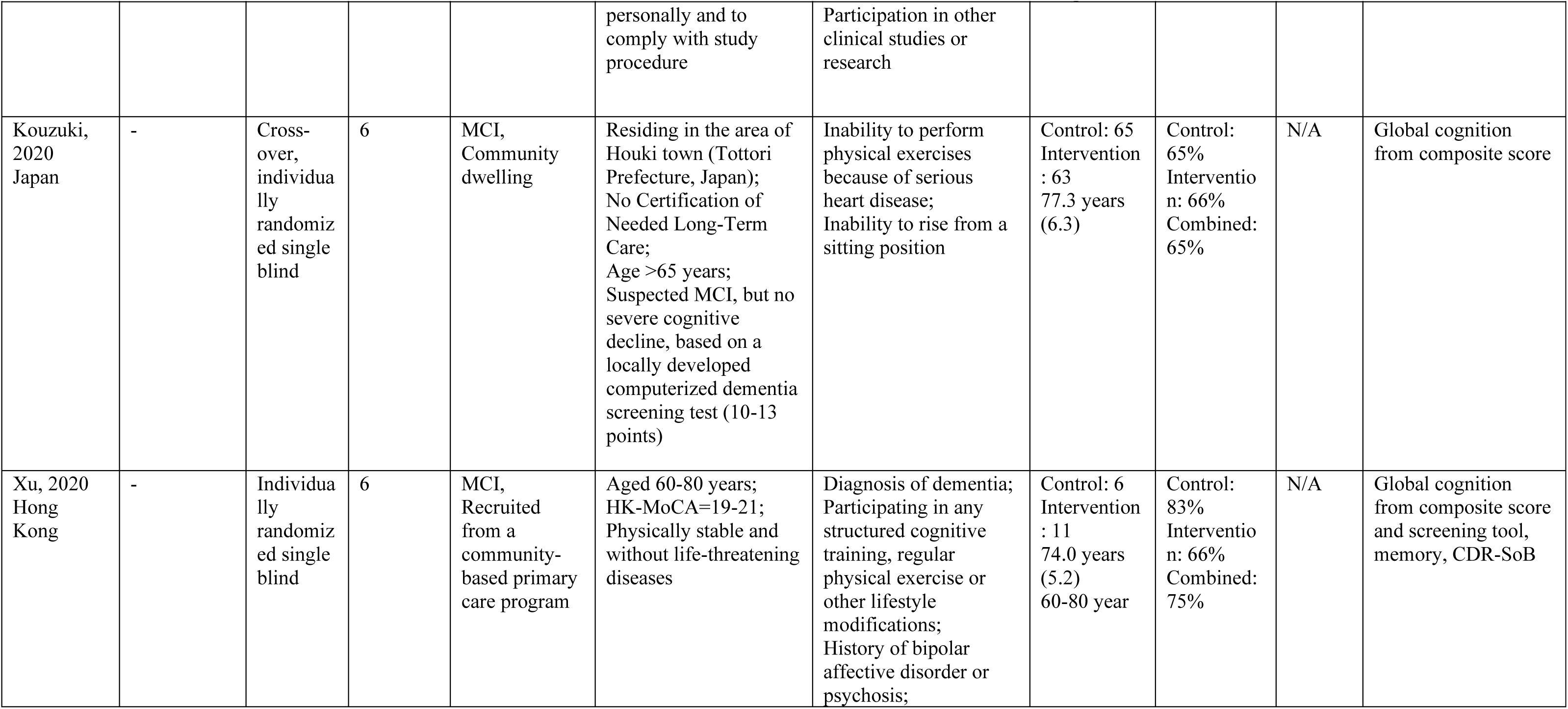
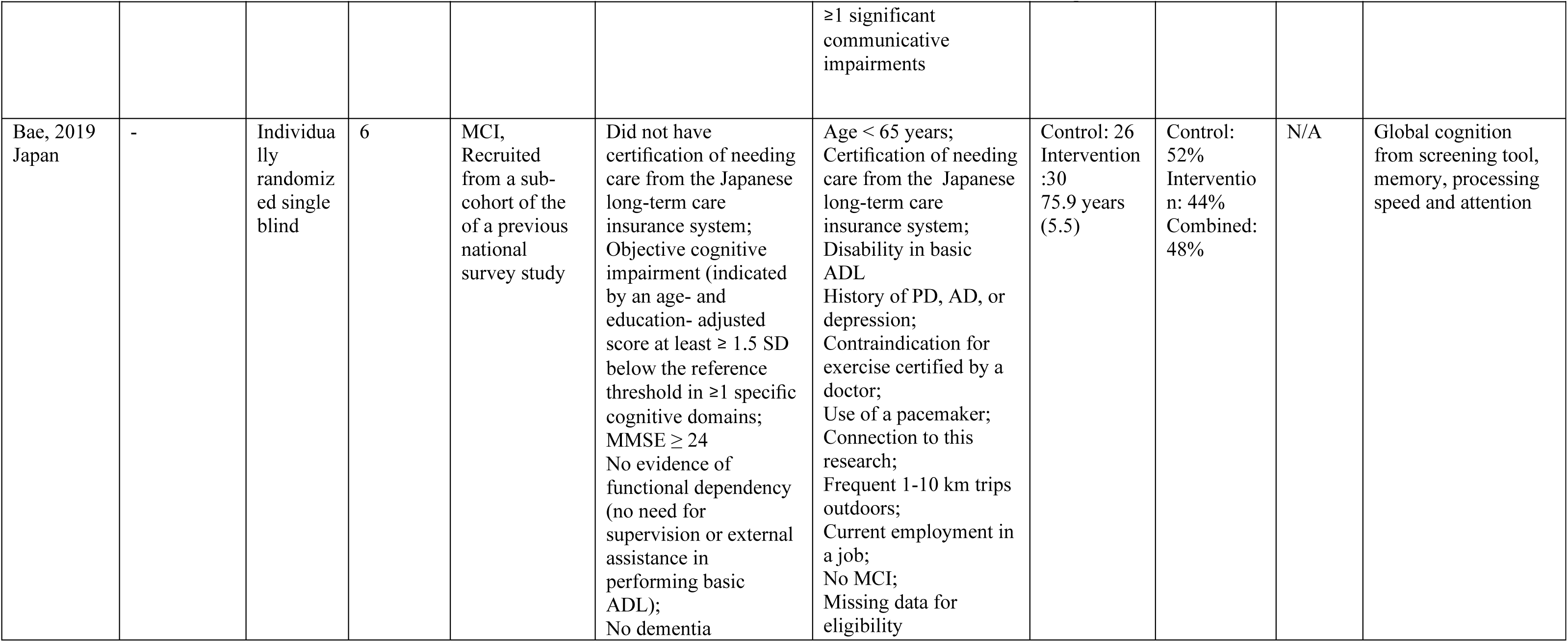
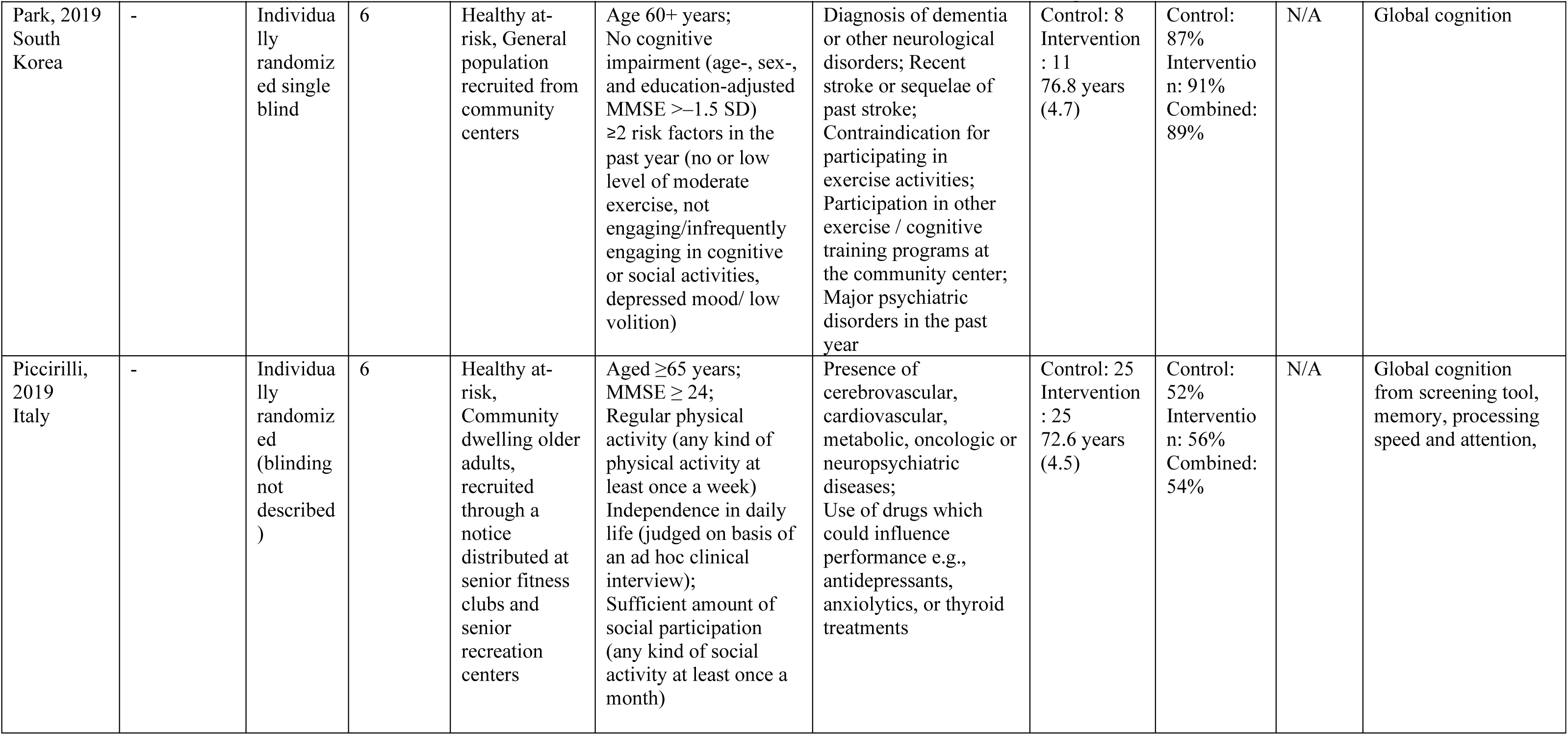
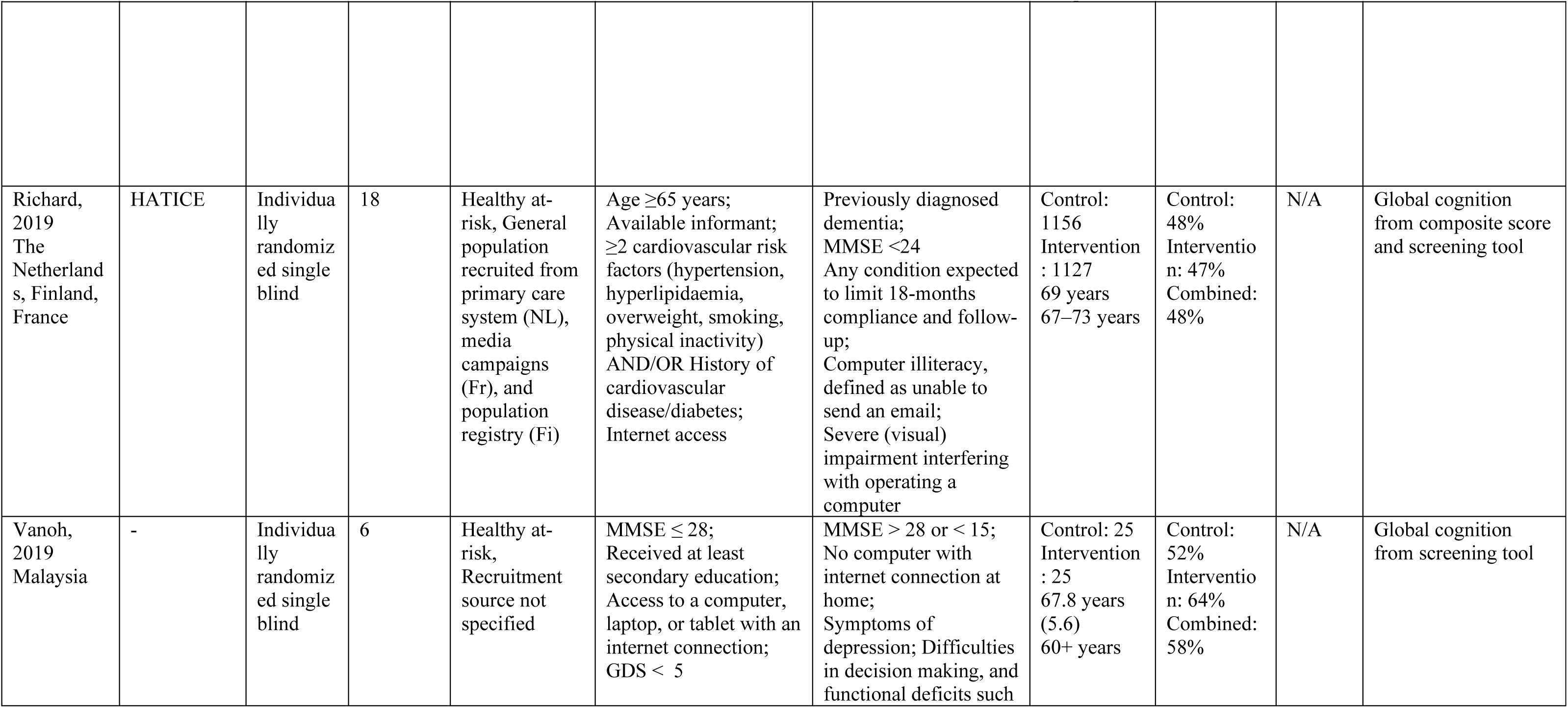
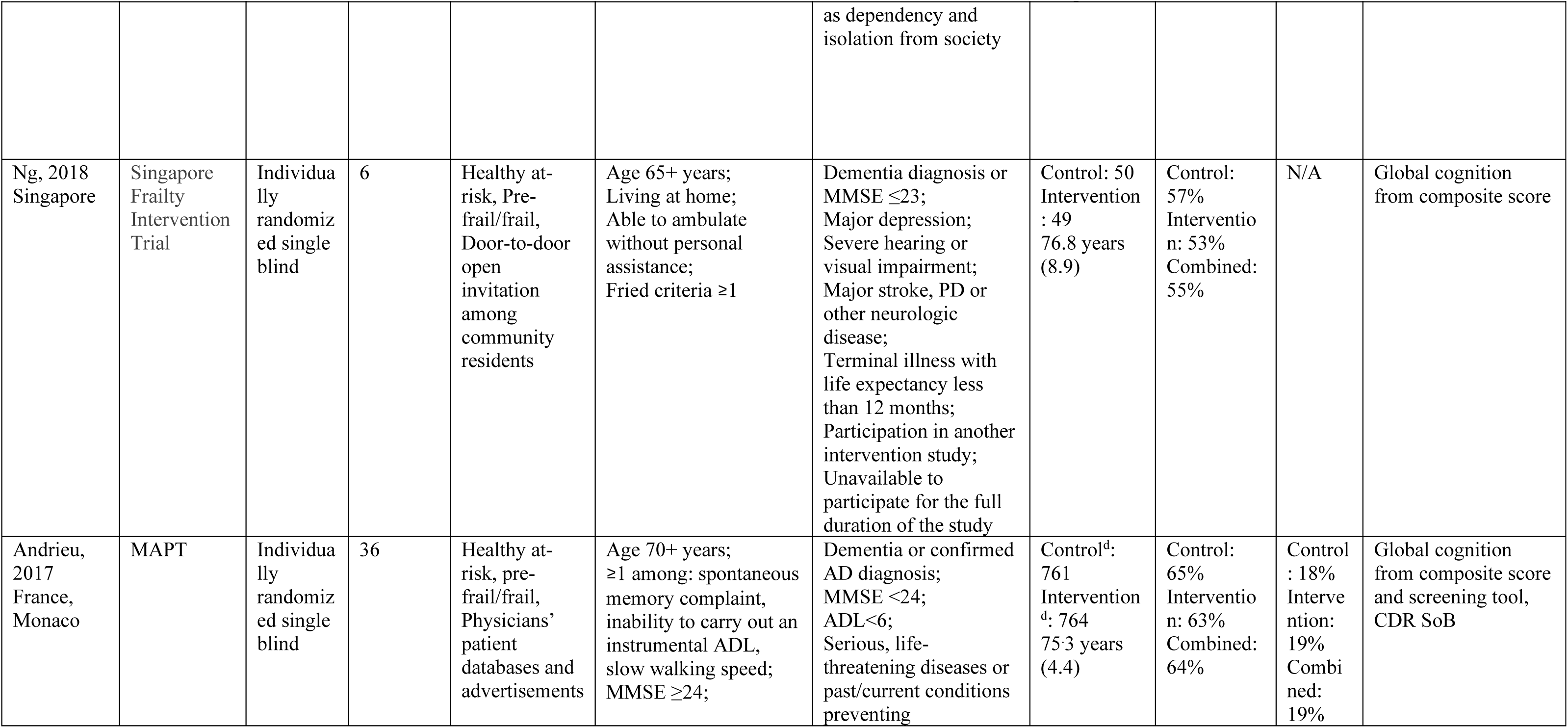
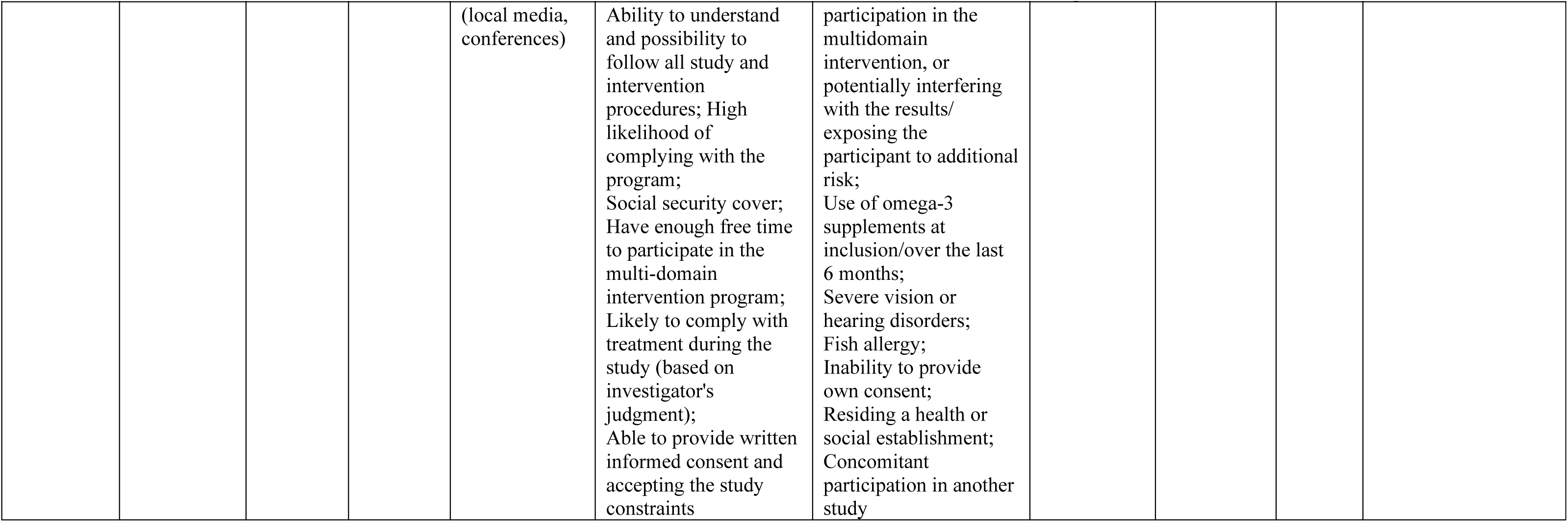
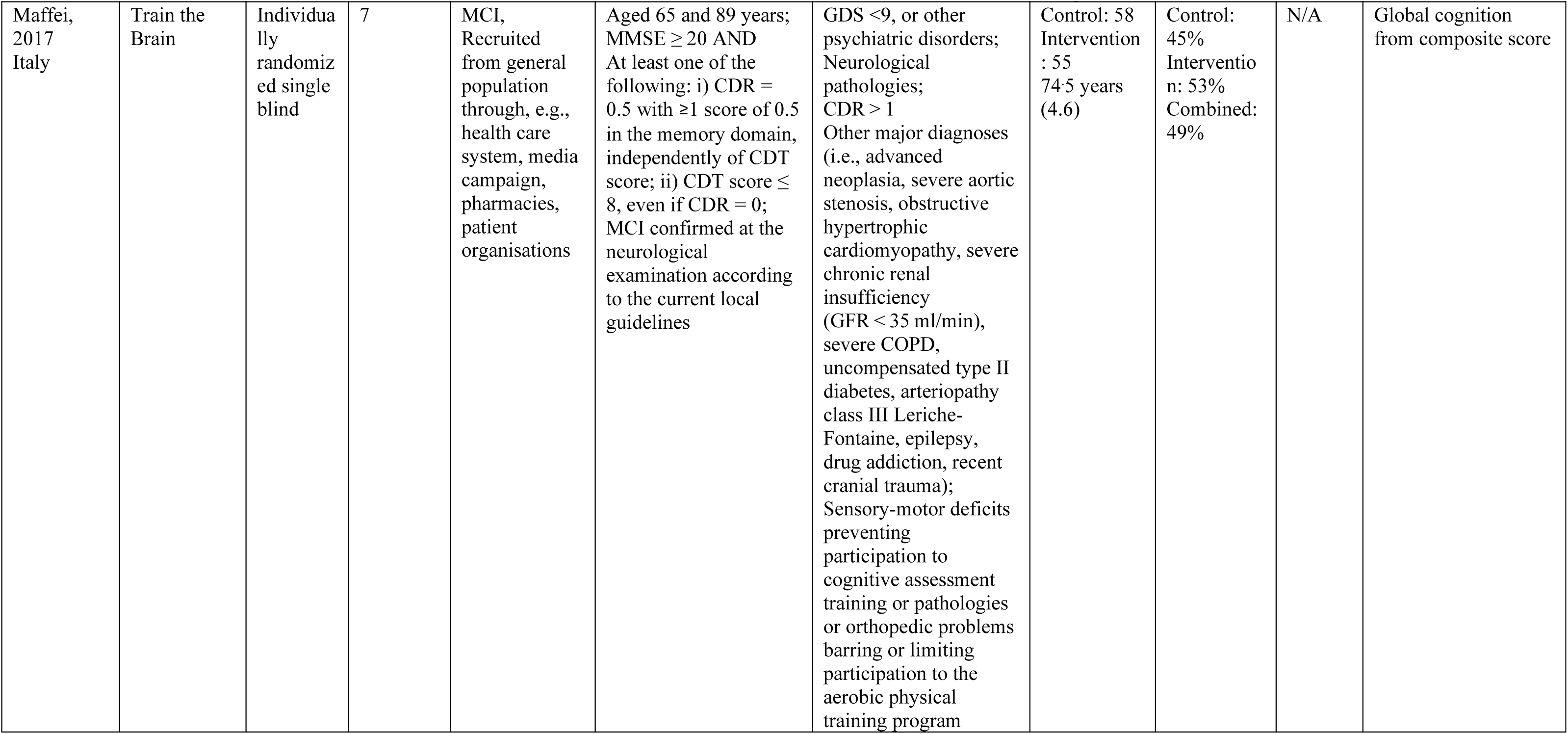
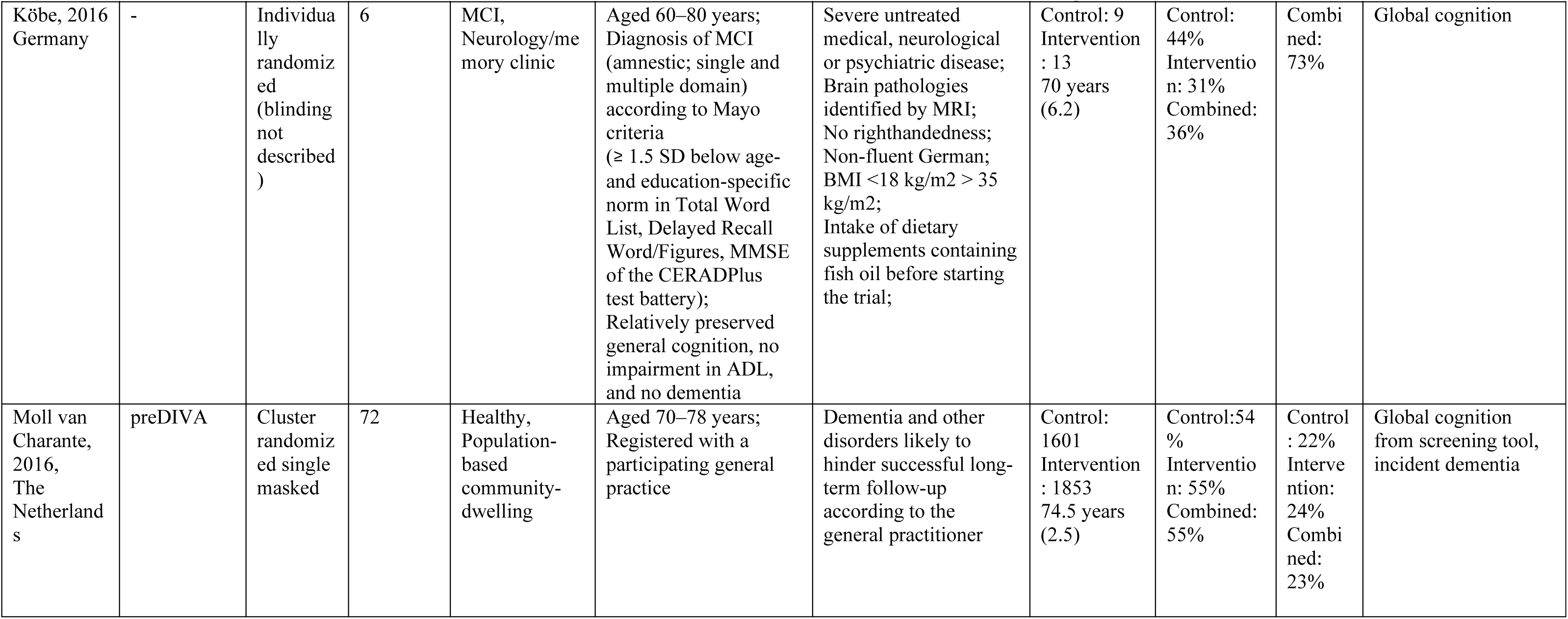
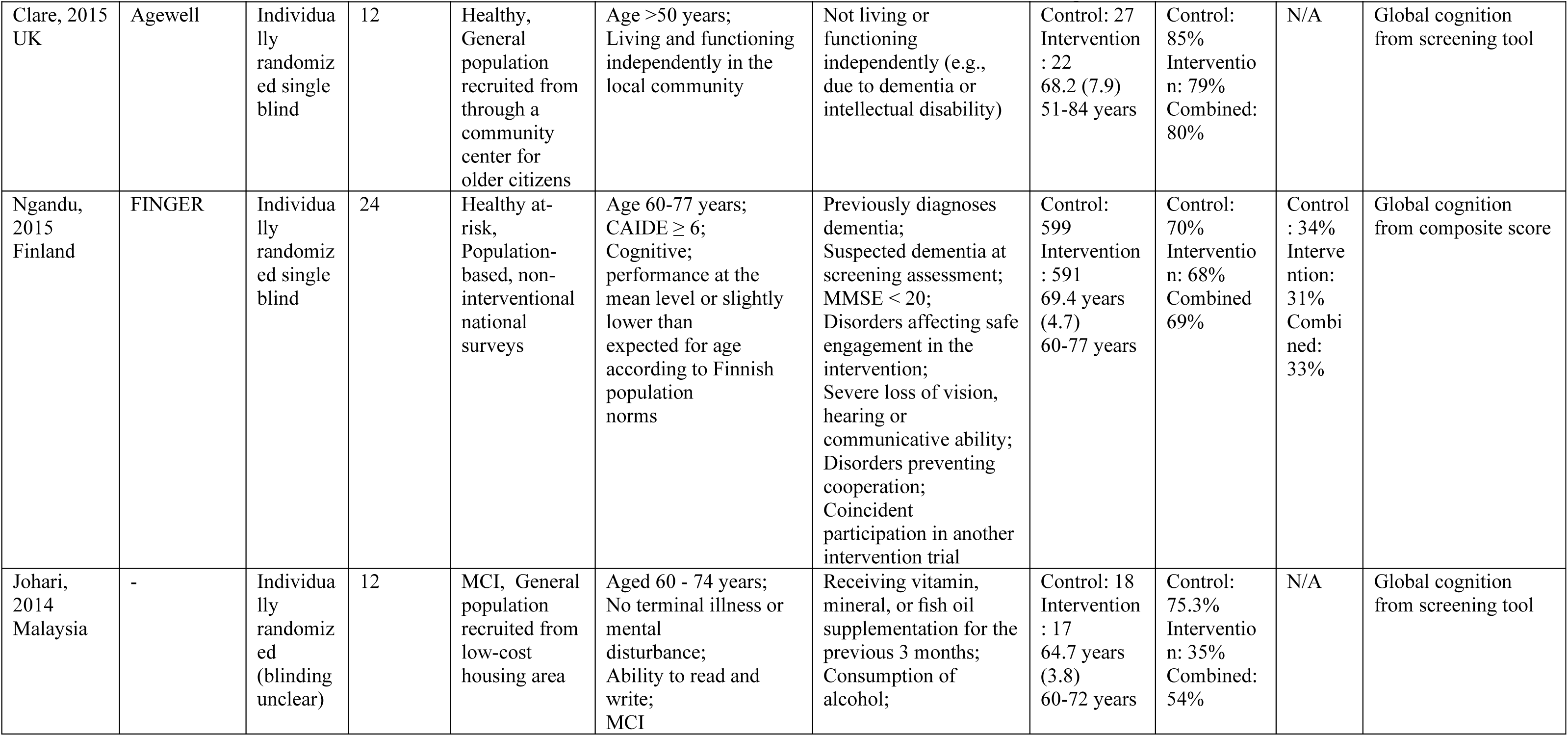
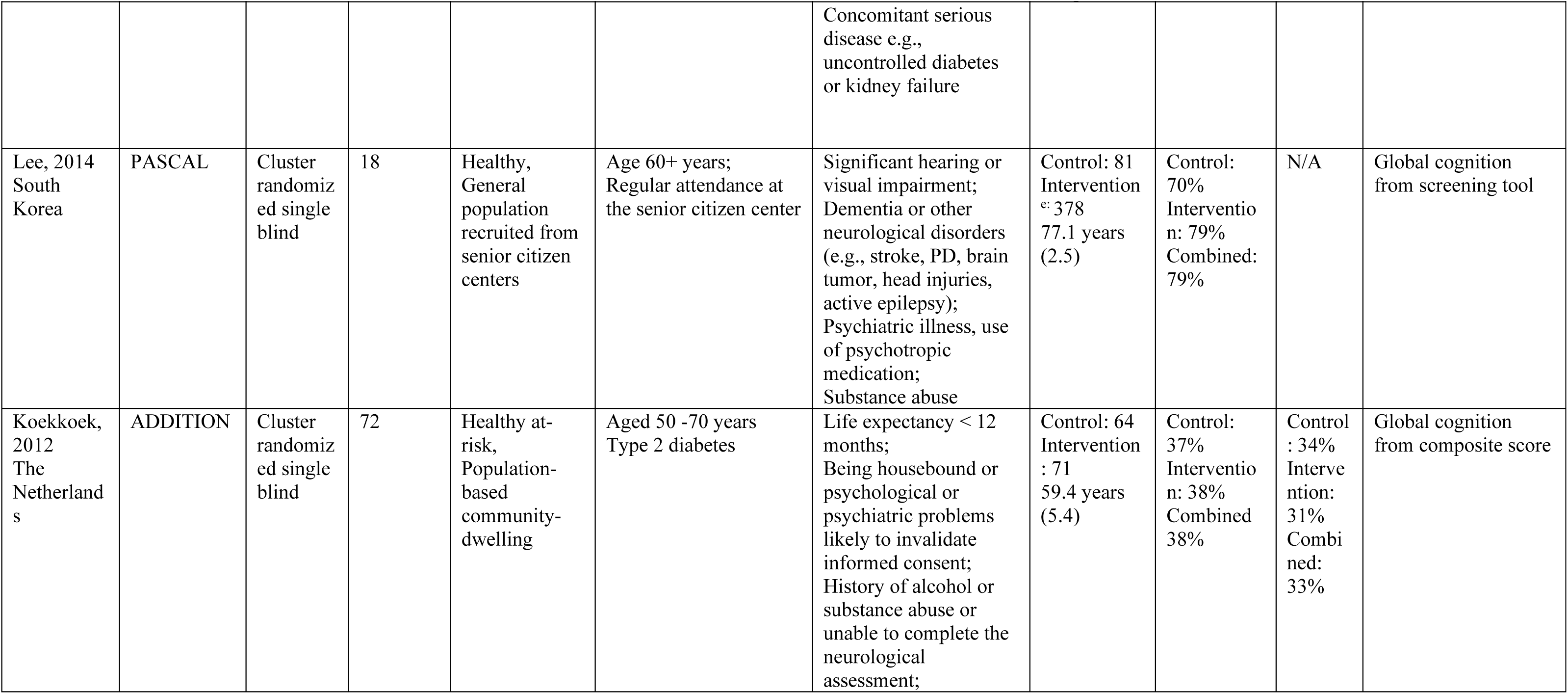
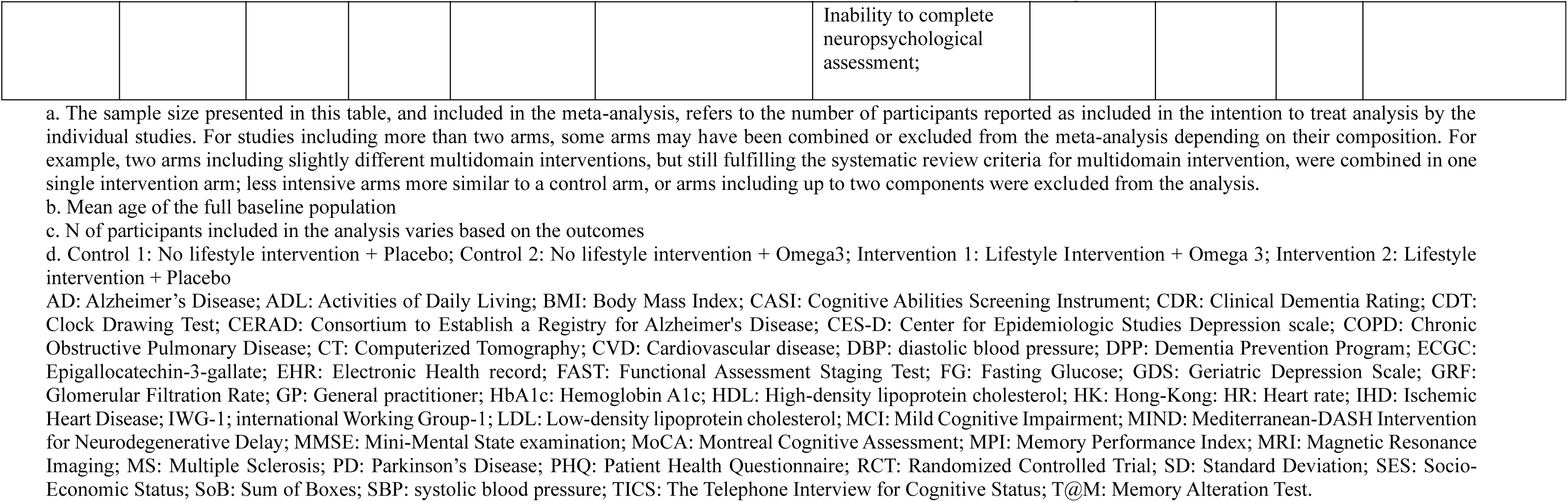
Details of the included RCTs’ study design (full study references provided in Supplementary. Table 4)

### Global cognition from composite score of validated neuropsychological tests

The pooled analysis from the 25 RCTs that measured global cognition using a composite score of validated neuropsychological tests showed significant beneficial intervention effects (RCTs=25; N=17,082; SMD=0.28; 95% CI: 0.10 to 0.45; Figure 2). Significant and high heterogeneity was estimated (Q<0.01; I^2^=96.17%; 95% PI=-0.602 to 1.154; Figure 2) and indicators of potential publication bias (Supplementary Figure 2; Supplementary Table 7) were identified, in connection with high heterogeneity. When the analysis was repeated including only studies with low or minimal risk of bias (RCTs=22; N=16,656) similar results (SMD=0.30; 95% CI: 0.10 to 0.49; Supplementary Figure 3, overall results for all panels) was observed, with similar indicators of heterogeneity (Q=0.05; I^2^=98.83%, 95% PI=-0.635, 1.232). From the sensitivity analysis excluding studies one at the time, in a stepwise manner, three^20–22^ were identified as important source of heterogeneity. When these were removed (Table 2; Supplementary Figure 4a; 22 RCTs; N=16,639) a smaller but still significant intervention effect was observed (SMD=0.13; 95% CI: 0.07 to 0.20), with significant, but substantially reduced, heterogeneity (Q<0.01, I2=59.42%; 95% PI=-0.077 to 0.344). Excluding smaller (N<100) RCTs (Table 2; Supplementary Figure 4b; 21 RCTs; N=16,846) also resulted in a smaller but still significant intervention effect (SMD=0.18; 95% CI: 0.06 to 0.30), but with a lesser impact on heterogeneity reduction (Q<0.01; I^2^=91.21; 95% PI=-0.351 to 0.710).

**Figure 2.**
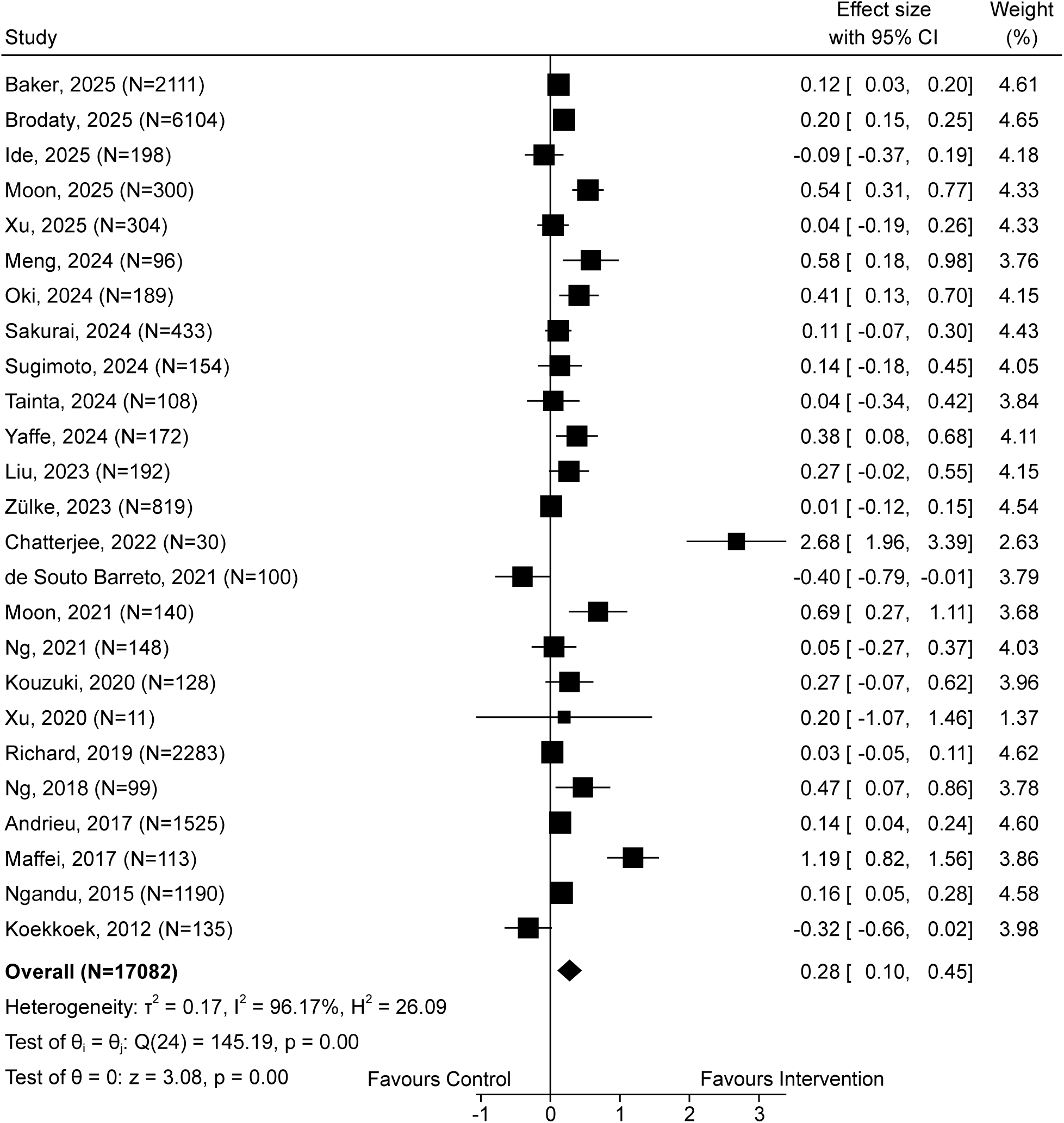
Meta-analysis of intervention effect on global cognition (composite score of validated neuropsychological tests)

**Table 2.**
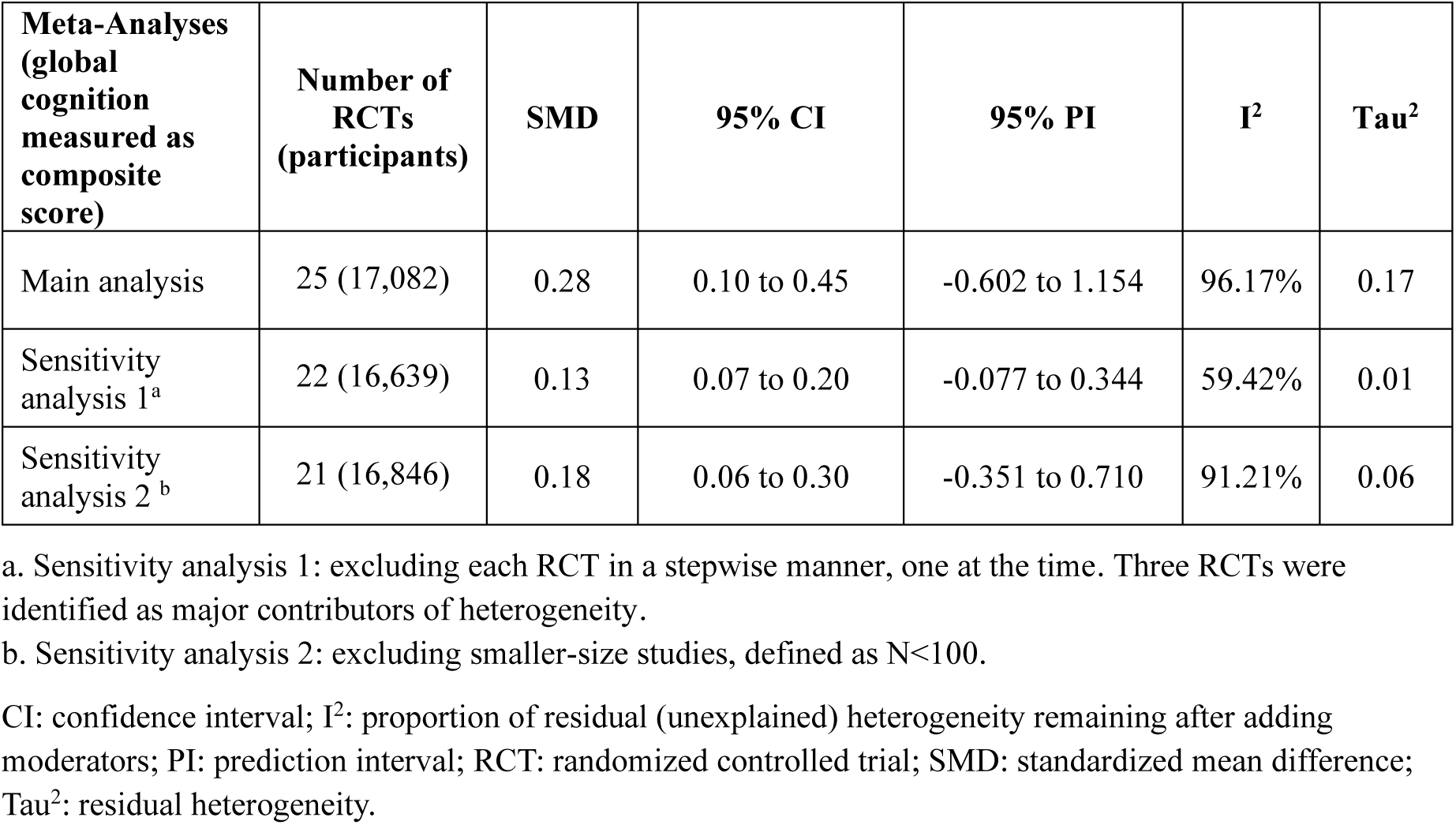
Meta-analysis of multidomain interventions effect on global cognition measured as a composite score of validated tests and corresponding sensitivity analyses.

| **Meta-Analyses**<br>**(global**<br>**cognition**<br>**measured as**<br>**composite**<br>**score)** | **Number of**<br>**RCTs**<br>**(participants)** | **SMD** | **95% CI** | **95% PI** | **I<sup>2</sup>** | **Tau<sup>2</sup>** |
| --- | --- | --- | --- | --- | --- | --- |
| Main analysis | 25 (17,082) | 0.28 | 0.10 to 0.45 | -0.602 to 1.154 | 96.17% | 0.17 |
| Sensitivity<br>analysis 1<sup>a</sup> | 22 (16,639) | 0.13 | 0.07 to 0.20 | -0.077 to 0.344 | 59.42% | 0.01 |
| Sensitivity<br>analysis 2<sup>b</sup> | 21 (16,846) | 0.18 | 0.06 to 0.30 | -0.351 to 0.710 | 91.21% | 0.06 |

a. Sensitivity analysis 1: excluding each RCT in a stepwise manner, one at the time. Three RCTs were identified as major contributors of heterogeneity.
b. Sensitivity analysis 2: excluding smaller-size studies, defined as N<100.
CI: confidence interval; I<sup>2</sup>: proportion of residual (unexplained) heterogeneity remaining after adding moderators; PI: prediction interval; RCT: randomized controlled trial; SMD: standardized mean difference; Tau<sup>2</sup>: residual heterogeneity.

Univariate meta-regression analyses (Table 3) showed a significant association between intervention effect-size and duration (P-value=0.009), but not with per-protocol intervention intensity (full set or sub-set of RCTs) target population selection (i.e., number of modifiable risk factors used both as intervention targets and eligibility criteria) or target population cognitive status (healthy at-risk vs MCI). Accordingly, in subgroup analysis significant difference was found only between RCTs with intervention <12 months and ≥12 months (Supplementary Figure 3). In the meta-analysis of RCTs with longer intervention (≥12 months), reduced heterogeneity was observed (Q=0.01; I2=41.07; 95% PI= −0.031;0.255; Supplementary Figure 3). Significant association was also found between intervention effect-size and intervention observed intensity, with larger effect being associated with higher observed intensity (P-value=0.008; Table 3).

**Table 3.**
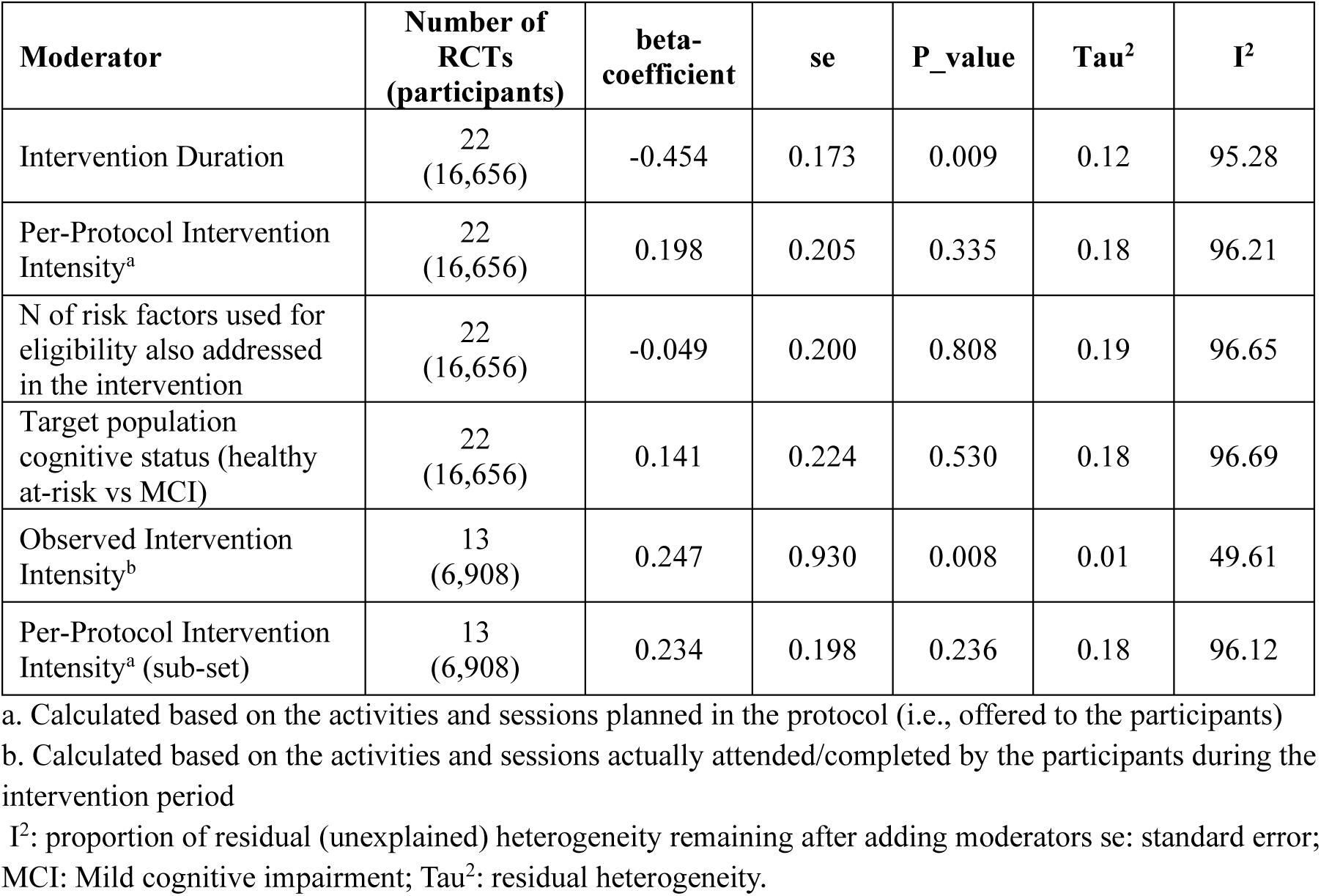
Meta regression analysis on the main cognitive outcome measure (global cognition measured as a composite multiple validated tests).

Results related to meta-analysis on other cognitive function measures (Supplementary Figures 4 and 5), GRADE assessment (Supplementary Table8), and incident MCI and dementia (Supplementary Figure 7) are presented in Supplementary Results.

## DISCUSSION

This up-to-date systematic review and meta-analysis included 41 RCTs, many of which had robust cognitive primary outcome measures. Pooled analysis (N=23,209) showed a significant, albeit small, intervention benefit on global cognition, memory, executive function, and processing speed. With only a minimal proportion at high-risk of bias, the RCTs’ quality was generally high and increased over time, reflecting the field’s rapid evolution. Although longer-term effects are less clear, this corroborates previous indications that multidomain interventions might reduce the risk of cognitive decline in older adults at-risk for dementia.^6^ A diverse range of populations and countries was represented, including some LMICs in Asia.^3,20^

The evidence certainty was graded “low” for global cognition measured as a composite score, due to high heterogeneity, and “high” for memory, executive function, and processing speed. Sensitivity analyses suggest that heterogeneity for global cognition could be partially explained by three smaller RCTs.^20–22^ However, when small RCTs (N<100) were excluded, the reduction in heterogeneity was limited, suggesting a more complex interplay of multiple factors. Neuropsychological test batteries measuring global cognition as a composite score are the combination of many different components, while domain-specific cognitive tests, even when reported as composite scores, include smaller subsets of tests across different RCTs, reducing variability. This could have played a role in our heterogeneity findings. Variability in intervention design, such as duration; intensity; and selection of target population, could also underlie the high heterogeneity and contribute to the observed small effect-size. For example, insufficiently intensive interventions may lead to not significant or smaller effects, irrespective of adherence; insufficient alignment of population selection with intervention targets may result in limited room for improvement; and duration may also affect adherence (e.g., reduced engagement over time for longer interventions), thus impacting effect-size.^6^

Longer intervention duration was associated with smaller but still significant effect-size, indicating the potential to provide long-term cumulative benefits. Heterogeneity was also substantially lower when longer intervention RCTs were pooled, suggesting a more reliable effect-size estimate. However, the sub-group analysis finding should be interpreted with caution, as engagement in lifestyle interventions often decreases over time^19,23^ which could explain the smaller effect-size in longer interventions. Higher adherence-adjusted observed intensity, reflecting actual intervention engagement, was also significantly associated with larger intervention benefits, confirming the crucial role of adherence. Preliminary evidence indicates that intervention related healthy lifestyle changes may be sustainable in the longer term^24^, supporting potential long-term effects on dementia risk-reduction. Per-protocol intensity and selection of the target population were not associated with intervention effect-size. Although the subgroup analysis by target population selection was not statistically significant, pooled analysis of RCTs targeting less than 3 modifiable risk factors also included as eligibility criteria did not show significant intervention effect. This suggests the importance of targeting the appropriate at-risk populations with sufficient potential for improvement. However, interventions with more components can also have more complex design, structure, and format variations, impacting adherence and efficacy. Therefore, a larger number of intervention components may not automatically translate into greater cognitive benefits, as indicated by a recent systematic review and network meta-analysis on single and multidomain interventions.^25^ No difference was observed in the meta-regression and sub-group analysis by target population cognitive status. As RCTs focusing on cognitively healthy populations do not systematically exclude MCI participants based on a full clinical workup, the presence of a certain proportion of participants with MCI at baseline in these RCTs cannot be completely ruled out, which would help explain this result. Results for global cognition measured with clinical screening tools were in line with those of cognitive domains, with small, statistically significant, beneficial intervention effects, and no heterogeneity. The lower heterogeneity likely reflects the use of the same two tools (Mini-Mental State Examination and Montreal Cognitive Assessment) in most RCTs. However, screening tools are less sensitive and more prone to a ceiling effect in cognitively unimpaired people.^26^ Given their limitations in general populations, they could be more useful for the selection of trial participants without dementia, than as outcome measures.

No significant intervention effect was found for clinical dementia rating sum-of-boxes (CDR-SoB;N=2280) and incident dementia (N=3,652), in a small number of RCTs. As diseases leading to dementia can take up to decades to develop, dichotomous outcomes such as incidence of dementia are unlikely to be feasible in early risk-reduction trials. CDR-SoB is now widely used as outcome in preclinical and prodromal AD drug trials but it is unclear whether it can capture the subtle cognitive-functional changes occurring in unimpaired at-risk general populations.^26^

Several limitations were identified. High heterogeneity in global cognition as composite score can hinder the pooling of RCTs and it could be only partially explained by our sensitivity analyses. While other factors such as adherence could play a role, adherence-adjusted observed intensity could be calculated only for 13 RCTs (N=6,908), limiting analytical power. Thus, interpretation of cognitive effect-sizes is currently unclear. Clinical meaningfulness of outcome measures is still being debated for both lifestyle and AD drug trials,^26^ and definition of meaningful benefits varies across risk and disease-stages as well as stakeholders. Few trials have, so far, reported intervention effects on surrogate outcomes such as markers of neurodegeneration or validated dementia risk-prediction tools.^27,28^ Only two RCTs reporting intervention effect on incident dementia were identified. Extended RCTs follow-up data from large multidomain RCTs and evidence on sustained, post-intervention, effect on dementia risk are currently lacking. With significant variability in protocols and limited harmonization across the 41 eligible RCTs, individual participants meta-analysis for dose-response assessment and intervention modifiers could provide additional insights but were not feasible. Factors such as sex, gender, and genetics might be particularly important modifiers of interventions response. However, an in-depth analysis would require individual participant data, or consistent sub-group measures reporting from individual RCTs. For example, a meta-analysis of three RCTs included in this review reported that *APOEε4* carriers might benefit more from multidomain lifestyle interventions.^29^ However, necessary data from sub-group analyses in the individual RCTs was also scarce and inconsistent. Finally, unlike single interventions targeting one risk factor at the time, multidomain interventions aim to target heterogeneous risk groups with different combinations of often interdependent risk factors. Their implementation, including tailoring to individual participants needs and habits, and stepwise introduction of activities, leads also to varying intensity per participant and across the intervention period. Therefore, it is not possible to accurately determine the most effective components for a multidomain intervention or directly compare effect sizes between single and multidomain interventions. Although beyond the scope of this study, meta-analyses of intervention effect on targeted risk factors could provide supporting evidence on this regard, but a more consistent reporting approach is needed for these outcomes.

Nonetheless, our findings suggest consistent, albeit small, beneficial effects of multidomain intervention on cognitive function, with intervention duration and adherence associated to intervention effects. By targeting risk factors shared with other late-life chronic non-communicable disorders (e.g., cardiovascular disease, diabetes), whilst focusing on populations and outcome measures relevant for dementia risk-reduction, multidomain interventions can have a wider impact on multiple health outcomes.^30^ More and better-quality evidence (e.g., lower heterogeneity) is needed from RCTs to capture the role of health and behavioral changes in dementia risk-reduction. Even if the effects on cognition observed at individual levels are modest, they might translate to substantial preventive effects at population level.^31^ Two of the included RCTs^32,33^ are indeed implementation studies embedded in the community and, importantly, some European countries have started embedding multidomain strategies in their national healthcare programs for the prevention of neurodegenerative diseases and dementia.^34,35^ These models can inform the definition of standards for multidomain intervention implementation and scalability in the general population.

In conclusion, future RCTs should prioritize harmonization of methodologies and consistent reporting (outcome measures and adherence, in particular), support long-term extended follow-up data, clinically relevant dementia-risk surrogate outcomes, and evidence from more diverse cultural, geographical, and socio-economic contexts, including cost-effectiveness and potential impact on health equity and sustainability. As 26 ongoing RCTs were identified during the systematic review, more evidence will be available in the coming years. Ongoing trials including also LMICs (e.g., Africa, Latin America), and joint international initiatives (i.e., World-Wide FINGERS^15^, International Research Network on Dementia Prevention,^36^ and European Platform for Neurodegenerative Disorders) will provide crucial additional evidence for the most understudied populations.

## Supporting information

PRISMA table

## Authors contributions

MB; FM; AS; RS; JL: study design. KK and RS: literature search. MB; FM; RS: screening, study selection, risk of bias assessment, and manuscript drafting. SA; MB; EK; JL; ASLR; AR; GS: data extraction. MB, EL, JL, FM, NSD, RTF, RS, TN, MP: data synthesis. MK, FM, AS, TN: securing funding. All authors revised and approved the final version of the manuscript.

## Data Availability

material from the meta-analysis we are open to share includes template data collection forms; data extracted from included studies; data used for all analyses; analytic code; any other materials used in the review, may be provided upon reasonable request to the corresponding author and upon approval of all authors.

## Acknowledgements

The authors would like to acknowledge the WHO team members, Tarun Dua, Katrin Seeher, Primrose Nyamayaro, and Ani Movsisyan for their leading role in the update of the 2019 WHO Guidelines for risk reduction of cognitive decline and dementia, which provided the base for this work.

## Material sharing

Template data collection forms; data extracted from included studies; data used for all analyses; analytic code; any other materials used in the review, may be provided upon reasonable request to the corresponding author and upon approval of all authors.

