## Supplemental Table 1-8 and figure 1-7 for "Multidomain lifestyle interventions for the risk reduction of cognitive impairment and dementia: A systematic review and meta-analysis"

#### SUPPLEMENTARY MATERIAL

Supplementary Figure 3. 53
Supplementary Figure 4. 54
Supplementary Figure 5. 55
Supplementary Figure 6. 56
Supplementary Figure 7. 57
5. Supplementary References 58

### 1. Supplementary Methods

#### 1.1. Eligibility criteria

Multidomain interventions were defined having minimum three components. This cut-off was deemed to better reflect the multifactorial etiopathogenesis of cognitive impairment and include interventions adaptable to individuals complex risk profiles. Other inclusion criteria were a randomized controlled design; and minimum intervention duration of six months, which was deemed as the minimum study duration that would reflect full engagement in a multidomain intervention, sustainable improvements, and meaningful cognitive changes. RCTs were excluded if included participants with suspected or known dementia diagnosis, other major neurological or psychiatric disorders; focused on rehabilitation programs for recent medical events and other conditions; included children, adolescents, or mostly young adults (age <40 years).

#### 1.2. Literature search and study selection

The search strategy was developed together with a health information specialist experienced in evidence retrieval and automation (KK) and cross-checked against a list of seed studies (for detailed search terms for each database see section 1.3. Full search terms for databases).
Randomized controlled trials (RCTs) included in eligible systematic reviews (SRs) were individually screened for eligibility. Potentially eligible studies published after the search date were manually identified until the 31st of August 2025 and included in the search results. Additionally, the status of RCTs that were identified as ongoing in the original screening (N=26) was re-assessed in April 2026 and none of those completed since (N=4) had yet been published. The reference lists of the included studies were also screened. No language restrictions were applied to the search. Authors were contacted if required data were not publicly available.
The records were imported and independently assessed in the Covidence systematic review software (https://app.covidence.org/reviews/active) by two researchers (MB, RS). Eligible articles after title/abstract screening were assessed at full text level. Disagreements were resolved with a third researcher (FM).

#### 1.3. Full search terms for databases

**EMBASE**
Database: Embase (Elsevier)
URL: https://www.embase.com/#advancedSearch/

| # | Searches |
|---|---|
| 1 | ((lower* OR reduc* OR loss OR losses OR slower OR better OR improv* OR effect OR Effects OR efficac* OR affect* OR impact* OR persist* OR further OR detect* OR change OR vary OR changing OR risk OR ratio* OR Odds OR impact OR detriment* OR influenc* OR maintenance OR protect* OR enhanc* OR prevent* OR preserv* OR greater OR restor* OR progress* OR adverse OR beneficial OR performance* OR score* OR compensat*) NEAR/8 (cognitive-measurement* OR cognitive-enhanc* OR Cognitive-improv* OR cognitive-outcome* OR cognitive-domain* OR cognitive-skill* OR cognitive-change* OR cognitive-performanc* OR neurocogniti* OR cognition* OR cognitive-function* OR cognitive-decline* OR cognitive-dysfunction* OR cognitive-impairment* OR cognitive-deficit* OR cognitive-behaviour* OR dementia* OR cognitive-abilit* OR alzheimer* OR cognitive-benefit* OR neurodegeneration OR neuro-degenerat* OR neuroprotective* OR MCI OR neurophysiologic* OR Neuropsycholog* OR psychosocial-function* OR psychosocial-outcome*)):ti,ab |
| 2 | ((multi* ) NEAR/2 (domain OR modal OR factor* OR component* OR dimension* OR facet*) NEAR/5 (intervention* OR program* OR Therap* OR treatment OR treated OR training OR activit*)):ti,ab OR ((multidomain OR multimodal OR multifactor* OR multicomponent* OR multidimension* Or multifacet*) NEAR/5 (intervention* OR program* OR Therap* OR treatment OR treated OR training OR activit*)):ti,ab OR ((combin*) NEAR/2 (intervention* OR program* OR training OR activit*)):ti,ab,kw OR world-wide-finger*:ti,ab OR ((lifestyle NEAR/8 diet NEAR/2 exercis*) OR (diet NEAR/2 exercis* NEAR/2 (pharma* OR medication* OR smok* OR drink*))):ti,ab |
| 3 | #1 AND #2 |
| 4 | [animals]/lim NOT ([animals]/lim AND [humans]/lim) |
| 5 | [letter]/lim OR [conference abstract]/lim OR [conference paper]/lim OR [conference review]/lim OR [editorial]/lim OR [note]/lim OR OR 'case report'/exp OR 'preprint'/exp |
| 6 | cancer:ti OR metastasis:ti OR tumour*:ti OR tumor*:ti OR neoplasm*:ti |
| 7 | #3 NOT (#4 OR #5 OR #6) |

**Medline**

Database: Medline (OVID)

| # | Searches |
|---|---|
| 1 | ((lower* OR reduc* OR loss OR losses OR slower OR better OR improv* OR effect OR Effects OR efficac* OR affect* OR impact* OR persist* OR further OR detect* OR change OR vary OR changing OR risk OR ratio* OR Odds OR impact OR detriment* OR influenc* OR maintenance OR protect* OR enhanc* OR prevent* OR preserv* OR greater OR restor* OR progress* OR adverse OR beneficial OR performance* OR score* OR compensat*) ADJ9 (cognitive-measurement* OR cognitive-enhanc* OR Cognitive-improv* OR cognitive-outcome* OR cognitive-domain* OR cognitive-skill* OR cognitive-change* OR cognitive-performanc* OR neurocogniti* OR cognition* OR cognitive-function* OR cognitive-decline* OR cognitive-dysfunction* OR cognitive-impairment* OR cognitive-deficit* OR cognitive-behaviour* OR dementia* OR cognitive-abilit* OR alzheimer* OR cognitive-benefit* OR neurodegeneration OR neuro-degenerat* OR neuroprotective* OR MCI OR neurophysiologic* OR Neuropsycholog* OR psychosocial-function* OR psychosocial-outcome* )).ti,ab |
| 2 | ((multi* ) ADJ2 (domain OR modal OR factor* OR component* OR dimension* OR facet*) ADJ5 (intervention* OR program* OR Therap* OR treatment OR treated OR training OR activit*)).ti,ab OR ((multidomain OR multimodal OR multifactor* OR multicomponent* OR multidimension* Or multifacet*) ADJ5 (intervention* OR program* OR Therap* OR treatment OR treated OR training OR activit*)).ti,ab OR ((combin*) ADJ2 (intervention* OR program* OR training OR activit*)).ti,ab,kw OR world-wide-finger*.ti,ab OR ((lifestyle ADJ8 diet ADJ2 exercis*) OR (diet ADJ2 exercis* ADJ2 (pharma* OR medication* OR smok* OR drink*))).ti,ab |
| 3 | 1 AND 2 |
| 4 | animals/ not (animals/ and humans/) |
| 5 | (letter or historical article or comment or editorial or news or case reports).pt. |
| 6 | 3 NOT (4 OR 5) |

**Cochrane**
Database: Cochrane Library
URL: https://www.cochranelibrary.com/advanced-search - Go to Search Manager tab

| # | Searches |
|---|---|
| 1 | ((lower* OR reduc* OR loss OR losses OR slower OR better OR improv* OR effect OR Effects OR efficac* OR affect* OR impact* OR persist* OR further OR detect* OR change OR vary OR changing OR risk OR ratio* OR Odds OR impact OR detriment* OR influenc* OR maintenance OR protect* OR enhanc* OR prevent* OR preserv* OR greater OR restor* OR progress* OR adverse OR beneficial OR performance* OR score* OR compensat*) NEAR/8 (cognitive-measurement* OR cognitive-enhanc* OR Cognitive-improv* OR cognitive-outcome* OR cognitive-domain* OR cognitive-skill* OR cognitive-change* OR cognitive-performanc* OR neurocogniti* OR cognition* OR cognitive-function* OR cognitive-decline* OR cognitive-dysfunction* OR cognitive-impairment* OR cognitive-deficit* OR cognitive-behaviour* OR dementia* OR cognitive-abilit* OR alzheimer* OR cognitive-benefit* OR neurodegeneration OR neuro-degenerat* OR neuroprotective* OR MCI OR neurophysiologic* OR Neuropsycholog* OR psychosocial-function* OR psychosocial-outcome*)):ti,ab |
| 2 | ((multi* ) NEAR/2 (domain OR modal OR factor* OR component* OR dimension* OR facet*) NEAR/5 (intervention* OR program* OR Therap* OR treatment OR treated OR training OR activit*)):ti,ab OR ((multidomain OR multimodal OR multifactor* OR multicomponent* OR multidimension* Or multifacet*) NEAR/5 (intervention* OR program* OR Therap* OR treatment OR treated OR training OR activit*)):ti,ab OR ((combin*) NEAR/2 (intervention* OR program* OR training OR activit*)):ti,ab,kw OR world-wide-finger*:ti,ab OR ((lifestyle NEAR/8 diet NEAR/2 exercis*) OR (diet NEAR/2 exercis* NEAR/2 (pharma* OR medication* OR smok* OR drink*))):ti,ab |
| 3 | #1 AND #2 |

**WHO ICTRP**
Database: WHO International Clinical Trials Registry Platform
URL: https://trialsearch.who.int/Default.aspx

| # | Search | Results |
|---|---|---|
| 1 | cognition OR cognitive OR dementia OR MCI OR "Neuro Degenerative" OR "Neuro degeneration" OR neuroprotective OR "neuro protective" | Title |
| 2 | "multi domain" OR "multi modal" OR "multi factorial" OR "multi factor" OR "multi component" OR "multi dimension" OR "multi facet" OR multidomain OR multicomponent OR multidimension OR multifacet OR multifactor OR multifactorial OR lifestyle AND diet AND exercise | intervention |

| | | |
|---|---|---|
| 3 | 1 AND 2 Filtered for date 1 Jan 2010 - 2 May 2025 | 49 |

Notes: cannot use two boolean operators in one field; cannot use brackets

**ClinicalTrials.gov**
Database: ClinicalTrials.gov
URL: https://clinicaltrials.gov/

| # | Search | Results | Field |
|---|---|---|---|
| 1 | cognition OR cognitive OR dementia OR alzheimers OR MCI OR "Neuro-Degenerative" OR "Neuro-degeneration" OR neuroprotective OR "neuro protective" | | Condition/Disease |
| 2 | "multi domain" OR "multi modal" OR "multi factorial" OR "multi factor" OR "multi component" OR "multi dimension" OR "multi facet" OR multidomain OR multicomponent OR multidimension OR multifacet OR multifactor OR multifactorial OR ((lifestyle AND diet AND exercise) OR (diet AND exercise AND (pharmacological OR pharmaceutical OR medication OR smoking OR drinking)) | | Intervention |
| | Combined 1 (condition) AND 2 (intervention) | **398** | |

Notes: filtering this resource by date was not possible

##### 1.4. Other cognitive function outcome measures

In addition to global cognition measured as composite score of validated neuropsychological tests in multiple domains (i.e., memory, attention, executive function, processing speed), other cognitive function outcome measures identified and included in the meta-analysis were memory, executive function, and processing speed (measured using validated neuropsychological tests); global cognition measured using validated screening tools for cognitive impairment (e.g., Mini-Mental State examination, MMSE; Montreal Cognitive Assessment, MoCA); Clinical Dementia Rating Sum of Boxes (CDR-SoB). For cognitive domains, standardized composite scores of multiple measures were included when available. When RCTs individually reported scores from multiple measures of the same domain without a standardized composite, only one outcome measure was pooled per RCT. The single outcome measure was selected based on whether it was frequently included in other studies, it was more comparable with similar measures included in other studies, or the level of validation.

##### 1.5. Data extraction and risk of bias

For each RCT, the data extracted were tabulated independently by two researchers (SA, EK, JL, ASRL, AR, GS) using an identical template and the output of the two independent data extractions was combined into a third table. The risk of bias (RoB) was assessed in relation to the cognitive function outcome. If a difference in the level of RoB was identified in the same RCT for different outcome measures included for the cognitive function outcome (e.g., different level of data missingness), the most conservative judgment (i.e., higher risk of bias) was made. Disagreements were resolved with a third researcher (FM). Each of the five domains (randomization; deviation from intervention; missing data; outcome measurement; reporting) was rated as ‘low risk’, ‘some concerns’, or ‘high risk’, with the study overall rate based on the highest rate recorded for any of the domains.

##### 1.6. Calculation of multidomain intervention Observed Intensity

Observed intensity was calculated as the ratio between the per-participant average number of intervention sessions attended/activities conducted (i.e., observed dose) and the study duration in months. The per-participant average observed dose was calculated based on publicly available data on adherence when available. The conversion was possible when the adherence was expressed as per-participant average ratio between observed dose and planned dose (the planned dose being the ratio between the per-protocol per-participant number of planned intervention and intervention duration), as follows:

$$\text{Adherence} = \frac{\text{Observed dose}}{\text{Planned dose}} \times 100 \longrightarrow \text{Observed intensity} = \frac{\text{Observed dose}}{\text{Study duration}}$$

As most RCTs report adherence per intervention component rather than for the intervention overall, in these cases the observed dose was calculated for each component and then combined. Data of observed intensity in the available RCTs is presented in eTable 2.

##### 1.7. Grading of Recommendations Assessment, Development and Evaluation (GRADE) assessment of the evidence

Grading of the evidence included the following criteria:
*Risk of Bias.* Evidence was downgraded if more than half of the overall domains across all studies included in each GRADE assessment were judged to be at higher risk than "low" (i.e., "some concerns" or "high-risk").
*Inconsistency.* Evidence was downgraded when heterogeneity was significant and more than "moderate".
*Indirectness.* Evidence was downgraded for indirectness if the researchers deemed that a substantial number of the studies included in each specific GRADE assessment did not fully match the PICO (Population, Intervention, Control, Outcome) question. For example, if the studies included mixed populations of young, mid-life, and older adults.
*Imprecision.* Result precision was judged mostly based on sample size and confidence interval. In particular, evidence was downgraded for sample size lower than 400 (200 per arm). When confidence intervals were wide and crossed the threshold for statistical significance, the evidence was also downgraded.
*Publication bias.* Evidence was downgraded when significant publication bias was identified based on Egger's test.

##### 1.8. Additional methodological aspects not included in the protocol registration

Although the PROSPERO registration described the plan for data synthesis, detailed methods could not be fully defined "a priori", due to differences in RCTs and intervention design, and the need for full details of eligible RCTs design and interventions. Therefore, additional methodological aspects, i.e., specific measures for the cognitive function outcome to be meta-analyzed; the use of meta-regression to explore factors associated with intervention effect-size; and the cut-offs for subgroup analyses had to be defined following data extraction. Additionally, since $I^2$ is an indicator of the proportion of variance that reflects variation in true effects, rather than an absolute measure of heterogeneity, 95% prediction intervals (PIs) were also included as indicators of heterogeneity.[1]

#### 2. Supplementary Results

##### 2.1. Description of eligible RCTs

Detailed description of the eligible RCTs is provided in Table 1 (study design, main manuscript) and eTable5 (intervention). In the 43 eligible RCTs (Table 1, main manuscript), sample size for cognitive function analysis ranged from 17 to 6104; range for mean age was 59.4-77.0 years; rate of female participants was 39%-90%; and APOEε4 carrier rate (available for 11 RCTs) 19%-73%. 15 RCTs were identified as focusing on a mild cognitive impairment (MCI)/prodromal-Alzheimer's disease (AD) population (based on different eligibility criteria), and 28 as focusing on a cognitively healthy/healthy at-risk population (although MCI participants were not specifically excluded via a full clinical work-up). Multidomain interventions had a varied range of designs eTable 5). Physical activity, diet, and cognitive training/stimulation were the most common components (33 trials included all three and ten a combination of two), followed by vascular/cardio-metabolic risk monitoring (26 RCTs) and social engagement (16 RCTs, normally incorporated with group activities for other components). Other components included, e.g., management of mental health risk factors, and supplementation/medical food. Interventions were mostly delivered with at least some in-person components (four RCTs fully remote) and with at least some and group activities (32 RCTs). The meta-analysis included 41 of the 43 eligible studies (N=23,209). Two eligible studies were excluded from the meta-analysis, due to the cognitive outcome (CDR-SoB) being reported as a dichotomous rather than a continuous variable (N=85);[2] or unclear reporting of statistical methods and results (N=19).[3]

##### 2.2. Risk of Bias assessment

When risk of bias was assessed, 48.8.0% (N=21) of eligible studies rated at "low risk", 39.5% (N=17) with "some concerns", and 11.6% (N=5) at "high risk" (eFigure 1a). Of the 220 bias domains assessed across the 43 studies, 82.8% (N=178) were rated at "low risk", 15.8% (N=34) with "some concerns", and 1.4% (N=3) at "high risk". "Selection of the reported result" was the most common downgraded domain and most of the domains with concerns were identified in older RCTs (eFigure 1b).

##### 2.3. Multidomain interventions effect on other cognitive function outcome measures

A small but significant intervention benefit was observed for all cognitive domains included (memory: RCTs=33, N=20,769, SMD=0.06, 95% CI: 0.01 to 0.10, eFigure 5a; executive function: RCTs=27, N=16,392, Standardized Mean Difference (SMD)=0.06, 95% Confidence Intervals (CI): 0.02 to 0.10, eFigure 5b; processing speed: RCTs=25, N=10,223, SMD=0.04, 95% CI: 0.00 to 0.08, eFigure 5c). Compared to global cognition measured as a composite score, lower heterogeneity was estimated for memory (Q=0.01; I2=35.44%, 95% PI= -0.080 to 0.194), executive function (Q<0.01; I2=18.03%; 95% PI=-0.037 to 0.155), and processing speed (Q =0.22; I2=0.00%; 95% PI= 0.001 to 0.082). A significant intervention benefit (RCTs=23; N=11,172; SMD=0.08; 95% CI: 0.03 to 0.13) without significant heterogeneity (Q=0.15; I2=27.04%; 95% PI=-0.049 to 0.212) was found similarly for global cognition measured using screening tools (eFigure 6a). No significant intervention effect was found for CDR-SoB (RCT=5; N=2,280; eFigure 6b).

##### 2.4. GRADE assessment

The evidence certainty assessed with GRADE (eTable7) of the effect of multidomain interventions on cognition measured using a composite score was "low" due to high heterogeneity. Evidence certainty was graded as "high" for domain-specific cognitive function measures and global cognition measured with screening tools".

##### 2.5. Multidomain interventions effect on incident MCI and dementia

Incident MCI was not included in the present meta-analysis, as no RCT reporting on intervention effect of MCI was identified. Two studies (N=3,652) were identified reporting incident dementia, and no significant intervention effect was found (eFigure 7).

#### 3. Supplementary Tables

##### Supplementary Table 1.

**Per-protocol Intensity of intervention in included studies (full study references provided in eTable4)**

| RCT | N sessions planned in the intervention | Intervention duration (months) | Per-protocol intervention intensity |
|---|---|---|---|
| Baker, 2025 | 855 | 24 | 35.6 |
| Brodaty, 2025 | 150 | 36 | 4.2 |
| Ide, 2025 | 210 | 18 | 11.7 |
| Moon, 2025 | 177 | 6 | 29.5 |
| Ponvel, 2025 | 389 | 24 | 16.2 |
| Xu, 2025 | 77 | 15 | 5.1 |
| Lee, 2024 | 16 | 12 | 1.3 |
| Meng, 2024 | 169 | 6 | 28.2 |
| Murukesu, 2024 | 54 | 6 | 9.0 |
| Oki, 2024 | 405 | 18 | 22.5 |
| Sakurai, 2024 | 245 | 18 | 13.6 |
| Sugimoto, 2024 | 48 | 18 | 2.7 |
| Tainta, 2024 | 156 | 12 | 13.0 |
| Yaffe, 2024 | 28 | 24 | 1.2 |
| Lee, 2023 | 54 | 10 | 5.4 |
| Liu, 2023 | 506 | 9 | 56.2 |
| Roach, 2023 | 778 | 24 | 32.4 |
| Zulke, 2023 | 527 | 24 | 22.0 |
| Chatterjee, 2022 | 118 | 6 | 19.7 |
| Kajita , 2022 | 20 | 30 | 0.7 |
| Yang , 2022 | 102 | 6 | 17.0 |
| de Souto Barreto, 2021 | 117 | 6 | 19.5 |
| Moon, 2021 | 169 | 6 | 28.2 |
| Ng, 2021 | 54 | 6 | 9.0 |
| Chen, 2020 | 16 | 12 | 1.3 |
| Kouzuki, 2020 | 6 | 6 | 1.0 |
| Xu, 2020 | 75 | 6 | 12.5 |
| Bae, 2019 | 48 | 6 | 8.0 |
| Piccirilli, 2019 | 52 | 6 | 8.7 |
| Richard, 2019 | 72 | 18 | 4.0 |
| Vanoh, 2019 | 107 | 6 | 17.8 |
| Ng, 2018 | 48 | 6 | 8.0 |
| Andrieu, 2017 | 43 | 36 | 1.2 |
| Maffei, 2017 | 182 | 7 | 26.0 |

| RCT | N sessions planned in the intervention | Intervention duration (months) | Per-protocol intervention intensity |
|---|---|---|---|
| Köbe, 2016 | 65 | 6 | 10.8 |
| Moll van Charante, 2016 | 18 | 72 | 0.3 |
| Clare, 2015 | 7 | 12 | 0.6 |
| Ngandu, 2015 | 256 | 24 | 10.7 |
| Johari, 2014 | 12 | 12 | 1.0 |
| Lee, 2014 | 11.6 | 18 | 0.6 |
| Koekkoek, 2012 | NA[a] | 72 | NA |

a. The exact number of sessions could not be calculated from the RCT published material
NA: Not available

**Supplementary Table 2.**
**Observed Intensity of intervention in included studies (full RCT references provided in eTable4)**

| RCT | Per-participant average number of sessions/activities planned/offered in the intervention | Per-participant average number of sessions completed/activities attended in the intervention | Intervention duration (months) | Observed intervention intensity |
|---|---|---|---|---|
| Ide, 2025 | 210 | 77.8 | 18 | 4.3 |
| Moon, 2025 | 177 | 153 | 6 | 25.5 |
| Ponvel, 2025 | 389 | 201.5 | 24 | 10.3 |
| Xu, 2025 | 77 | 47 | 15 | 3.1 |
| Meng, 2024 | 169 | 115 | 6 | 19.2 |
| Murukesu, 2024 | 54 | 53 | 6 | 8.8 |
| Sakurai, 2024 | 245 | 146 | 18 | 8.1 |
| Tainta, 2024 | 156 | 99 | 12 | 8.3 |
| Yaffe, 2024 | 28 | 18.8 | 24 | 0.8 |
| Lee, 2023 | 54 | 45 | 10 | 4.5 |
| Liu, 2023 | 506 | 268 | 9 | 29.8 |
| Moon, 2021 | 169 | 162 | 6 | 27 |
| Ng, 2021 | 54 | 41.5 | 6 | 6.9 |
| Xu, 2020 | 75 | 41 | 6 | 6.8 |
| Bae, 2019 | 48 | 35 | 6 | 5.8 |
| Richard, 2019 | 72 | 32 | 18 | 1.8 |
| Ng, 2018 | 48 | 42.2 | 6 | 7 |
| Andrieu, 2017 | 43 | 28.9 | 36 | 0.8 |
| Ngandu, 2015 | 256 | 97.3 | 24 | 4.1 |

#### Supplementary Table 3.

**Selection of target population based on eligibility criteria specifically addressed in the multidomain intervention (full RCT references provided in eTable4)**

| RCT | Modifiable risk factors part of eligibility criteria and/or cognitive performance criteria/status included for participants selection | Intervention components | N modifiable risk factors specifically addressed in the multidomain intervention |
|---|---|---|---|
| Baker, 2025 | Physical activity<br>Diet<br>Cardiometabolic risk factors | Physical activity<br>Diet/Nutrition/Weight management<br>Cardiometabolic risk factors management<br>Cognitive training<br>Social engagement | 3 |
| Brodaty, 2025 | Physical activity<br>Diet/Nutrition<br>Cognitive activity<br>Mental health risk factors | Physical activity<br>Diet/Nutrition/Weight management<br>Cognitive training<br>Mental health risk factors management | 4 |
| Ide, 2025 | BMI<br>Cardiometabolic risk factors | Diet/Nutrition/Weight management<br>Cardiometabolic risk factors management<br>Physical Activity<br>Cognitive training<br>Oral hygiene | 2 |
| Moon, 2025 | Cardiometabolic risk factors<br>BMI<br>Cognitive performance | Cardiometabolic risk factors management<br>Diet/Nutrition/Weight management<br>Cognitive Training<br>Physical activity<br>Mental health risk factors management | 3 |
| Ponvel, 2025 | Cognitive performance<br>Functional performance<br>Frailty | Cognitive training<br>Diet/Nutrition/Weight management<br>Cardiometabolic risk factors management<br>Physical activity<br>Social engagement | 3 |
| Xu, 2025 | Cognitive performance | Cognitive training<br>Cardiometabolic risk factors management<br>Physical activity | 1 |
| Lee, 2024 | Cardiometabolic risk factors | Cardiometabolic risk factors management<br>Diet/Nutrition/Weight management<br>Physical | 1 |

| RCT | Modifiable risk factors part of eligibility criteria and/or cognitive performance criteria/status included for participants selection | Intervention components | N modifiable risk factors specifically addressed in the multidomain intervention |
|---|---|---|---|
| | | Cognitive training | |
| Meng, 2024 | Physical activity<br>Cardiometabolic risk factors<br>Mental health risk factors<br><br>Social activity<br>Subjective cognition | Physical activity<br>Cardiometabolic risk factors management<br>Mental health risk factors management<br>Social engagement<br>Cognitive training<br>Diet/Nutrition/Weight management<br>Dementia Literacy | 5 |
| Murukesu, 2024 | Physical function<br>Frailty | Physical activity<br>Diet/Nutrition/Weight management<br>Cognitive training<br>Social engagement | 2 |
| Oki, 2024 | Cognitive performance<br>Functional performance<br>Cardiometabolic risk factors | Cognitive training<br>Physical activity<br>Cardiometabolic risk factors management<br>Diet/Nutrition/Weight management | 3 |
| Sakurai, 2024 | Cognitive performance | Cognitive training<br>Physical activity<br>Cardiometabolic risk factors management<br>Diet/Nutrition/Weight management | 1 |
| Sugimoto, 2024 | Cardiometabolic risk factors<br>Cognitive performance | Cardiometabolic risk factors management<br>Physical activity<br>Diet/Nutrition/Weight management<br>Social engagement | 1 |
| Tainta, 2024 | BMI<br><br>Cardiometabolic risk factors<br>Physical activity<br>Cognitive performance | Diet/Nutrition/Weight management<br>Cardiometabolic risk factors management<br>Physical activity<br>Cognitive training<br>Social engagement | 4 |
| Yaffe, 2024 | Physical activity<br><br>Cardiometabolic risk factors<br>Mental health risk factors<br>Social activity | Personalised management of risk factors | 4 |

| RCT | Modifiable risk factors part of eligibility criteria and/or cognitive performance criteria/status included for participants selection | Intervention components | N modifiable risk factors specifically addressed in the multidomain intervention |
|---|---|---|---|
| Zulke, 2023 | BMI<br><br>Cardiometabolic risk factors<br>Physical activity | Diet/Nutrition/Weight management<br>Cardiometabolic risk factors management<br>Physical activity<br>Cognitive training<br><br>Social engagement | 3 |
| Lee, 2023 | Cognitive performance | Cognitive training<br>Physical activity<br>Social engagement | 1 |
| Liu, 2023 | Subjective cognition & Cognitive performance<br>Physical activity<br>Mental health risk factors<br><br>Cardiometabolic risk factors | Cognitive training<br><br>Physical activity<br>Mental health risk factors management<br>Diet/Nutrition/Weight management | 3 |
| Roach, 2023 | Cognitive performance | Cognitive training<br>Physical activity<br>Mental health risk factors management<br>Diet/Nutrition/Weight management | 1 |
| Chatterjee, 2022 | Subjective cognition | Cognitive training<br>Physical activity<br>Diet/Nutrition/Weight management | 1 |
| Kajita, 2022 | Cognitive performance | Cognitive training<br>Physical activity<br>Diet/Nutrition/Weight management<br>Dementia literacy | 1 |
| Yang, 2022 | Cognitive performance | Cognitive training<br>Physical activity<br>Diet/Nutrition/Weight management<br>Cardiometabolic risk factors management | 1 |
| de Souto Barreto, 2021 | Subjective cognition | Cognitive training<br>Physical activity<br>Diet/Nutrition/Weight management | 1 |
| Moon, 2021 | Cardiometabolic risk factors<br>BMI | Cardiometabolic risk factors management<br>Diet/Nutrition/Weight management | 3 |

| RCT | Modifiable risk factors part of eligibility criteria and/or cognitive performance criteria/status included for participants selection | Intervention components | N modifiable risk factors specifically addressed in the multidomain intervention |
|---|---|---|---|
| | Physical activity<br>Social activity | Physical activity<br>Mental health risk factors management | |
| Ng, 2021 | Physical activity<br>BMI<br><br>Social activity<br>Cardiometabolic risk factors<br>Mental health risk factors | Physical activity<br>Diet/Nutrition/Weight management<br>Cognitive training | 2 |
| Chen, 2020 | Subjective cognition<br>Physical function | Cognitive training<br>Physical activity<br>Diet/Nutrition/Weight management<br>Cardiometabolic risk factors management | 2 |
| Kouzuki, 2020 | Cognitive performance | Cognitive training<br>Physical activity<br>Dementia literacy | 1 |
| Xu, 2020 | Cognitive performance | Cognitive training<br>Physical activity<br>Dementia literacy<br>Diet/Nutrition/Weight management | 1 |
| Bae, 2019 | Cognitive performance<br>Physical activity | Cognitive training<br>Physical activity<br>Social engagement | 2 |
| Piccirilli, 2019 | NA | Cognitive training<br>Physical activity<br>Social engagement | 0 |
| Richard, 2019 | Physical activity<br>BMI<br><br>Cardiometabolic risk factors | Physical activity<br>Diet/Nutrition/Weight management<br>Cardiometabolic risk factors management | 3 |
| Vanoh, 2019 | Cognitive performance | Physical activity<br>Cardiometabolic risk factors management | 0 |
| Ng, 2018 | Frailty | Cognitive training<br>Physical activity<br>Diet/Nutrition/Weight management | 1 |
| Andrieu, 2017 | Subjective cognition | Cognitive training | 2 |

| RCT | Modifiable risk factors part of eligibility criteria and/or cognitive performance criteria/status included for participants selection | Intervention components | N modifiable risk factors specifically addressed in the multidomain intervention |
|---|---|---|---|
| | Physical function | Physical activity<br>Diet/Nutrition/Weight management<br>Cardiometabolic risk factors management | |
| Maffei, 2017 | Cognitive performance<br>Functional performance | Cognitive training<br>Physical activity<br>Diet/Nutrition/Weight management | 2 |
| Kobe, 2016 | Cognitive performance | Cognitive training<br>Physical activity<br>Diet/Nutrition/Weight management | 1 |
| Moll van Charante, 2016 | NA | Personalised management of risk factors | 0 |
| Clare, 2015 | NA | Cognitive training<br>Physical activity<br>Social engagement | 0 |
| Ngandu, 2015 | BMI<br><br>Cardiometabolic risk factors<br>Physical activity<br>Cognitive performance | Diet/Nutrition/Weight management<br>Cardiometabolic risk factors management<br>Physical activity<br>Cognitive training<br>Social engagement | 4 |
| Johari, 2014 | Cognitive performance | Cognitive training<br>Physical activity<br>Diet/Nutrition/Weight management | 1 |
| Lee, 2014 | NA | Cognitive training<br>Physical activity<br>Diet/Nutrition/Weight management<br>Cardiometabolic risk factors management<br>Social engagement<br>Dementia literacy | 0 |
| Koekkoek, 2012 | Cardiometabolic risk factors<br>BMI | Cardiometabolic risk factors management<br>Diet/Nutrition/Weight management<br>Physical activity | 2 |

NA: Not available

#### Supplementary Table 4.
**Full references of included RCTs**

| First Author, Year | RCT Name | Full Reference |
|---|---|---|
| Andrieu, 2017 | MAPT | Andrieu S, Guyonnet S, Coley N, Cantet C, Bonnefoy M, Bordes S, et al. Effect of long-term omega 3 polyunsaturated fatty acid supplementation with or without multidomain intervention on cognitive function in elderly adults with memory complaints (MAPT): a randomized, placebo-controlled trial. Lancet Neurol. 2017;16(5):377–89. |
| Bae, 2019 | NA | Bae S, Lee S, Lee S, Jung S, Makino K, Harada K, et al. The effect of a multicomponent intervention to promote community activity on cognitive function in older adults with mild cognitive impairment: A randomized controlled trial. Complement Ther Med [Internet]. 2019;42(September 2018):164–9. Available from: https://doi.org/10.1016/j.ctim.2018.11.011 |
| Baker, 2025 | US POINTER | Baker LD, Espeland MA, Whitmer RA, Snyder HM, Leng X, Lovato L, et al. Structured vs Self-Guided Multidomain Lifestyle Interventions for Global Cognitive Function: The US POINTER Randomized Clinical Trial. JAMA [Internet]. 2025 Jul 28; Available from: https://doi.org/10.1001/jama.2025.12923 |
| Brodaty, 2025 | MYB | Brodaty H, Chau T, Heffernan M, Ginige JA, Andrews G, Millard M, et al. An online multidomain lifestyle intervention to prevent cognitive decline in at-risk older adults: a randomized controlled trial. Nat Med. 2025;31(2):565–73. |
| Chatterjee, 2022 | MISCI | Chatterjee P, Kumar DA, Naqushbandi S, Chaudhary P, Khenduja P, Madan S, et al. Effect of Multimodal Intervention (computer based cognitive training, diet and exercise) in comparison to health awareness among older adults with Subjective Cognitive Impairment (MISCI-Trial)-A Pilot Randomized Control Trial. PLoS One [Internet]. 2022;17(11 November):1–13. Available from: http://dx.doi.org/10.1371/journal.pone.0276986 |
| Chen, 2020 | THISCE | Chen LK, Hwang AC, Lee WJ, Peng LN, Lin MH, Neil DL, et al. Efficacy of multidomain interventions to improve physical frailty, depression and cognition: data from cluster-randomized controlled trials. J Cachexia Sarcopenia Muscle. 2020;11(3):650–62. |
| Clare, 2015 | Agewell | Clare L, Nelis SM, Jones IR, Hindle J V, Thom JM, Nixon JA, et al. The Agewell trial: a pilot randomized controlled trial of a behaviour change intervention to promote healthy ageing and reduce risk of dementia in later life. BMC Psychiatry [Internet]. 2015;15(1):25. Available from: https://doi.org/10.1186/s12888-015-0402-4 |
| de Souto Barreto, 2021 | eMIND | de Souto Barreto P, Pothier K, Soriano G, Lussier M, Bherer L, Guyonnet S, et al. A Web-Based Multidomain Lifestyle Intervention for Older Adults: The eMIND Randomized Controlled Trial. J Prev Alzheimer's Dis [Internet]. 2021;8(2):142–50. Available from: https://www.sciencedirect.com/science/article/pii/S2274580724003832 |
| Ide, 2025 | J-MINT PRIME Kanagawa | Ide K, Mizushima S, Saito K, Suzuki H, Chiba Y, Abe K, et al. Japan-multimodal intervention trial for the prevention of dementia in older people with lifestyle-related diseases: A community-based, 18-month, randomized controlled trial. J Alzheimers Dis. 2025 Jul;106(2):574–88. |
| Johari, 2014 | NA | Johari SM, Shahar S, Ng TP, Rajikan R. A preliminary randomized controlled trial of multifaceted educational intervention for mild cognitive impairment among elderly Malays in Kuala Lumpur. Int J Gerontol [Internet]. 2014;8(2):74–80. Available from: http://dx.doi.org/10.1016/j.ijge.2013.07.002 |
| Kajita, 2022 | NA | Kajita H, Maeda K, Osaki T, Kakei Y, Kothari KU, Nagai Y. The effect of a multimodal dementia prevention program involving community-dwelling elderly. Psychogeriatrics. 2022;22(1):113–21. |

| First Author, Year | RCT Name | Full Reference |
|---|---|---|
| Kouzuki, 2020 | NA | Kouzuki M, Kato T, Wada-Isoe K, Takeda S, Tamura A, Takanashi Y, et al. A program of exercise, brain training, and lecture to prevent cognitive decline. Ann Clin Transl Neurol. 2020;7(3):318–28. |
| Köbe, 2016 | NA | Köbe T, Witte AV, Schnelle A, Lesemann A, Fabian S, Tesky VA, et al. Combined omega-3 fatty acids, aerobic exercise and cognitive stimulation prevents decline in gray matter volume of the frontal, parietal and cingulate cortex in patients with mild cognitive impairment. Neuroimage. 2016 May;131:226–38. |
| Koekkoek, 2012 | ADDITION | Koekkoek PS, Ruis C, van den Donk M, Biessels GJ, Gorter KJ, Kappelle LJ, Rutten GE. Intensive multifactorial treatment and cognitive functioning in screen-detected type 2 diabetes--the ADDITION-Netherlands study: a cluster-randomized trial. J Neurol Sci. 2012 Mar 15;314(1-2):71-7. doi: 10.1016/j.jns.2011.10.028. |
| Lee, 2014 | PASCAL | Lee KS, Lee Y, Back JH, Son SJ, Choi SH, Chung YK, et al. Effects of a multidomain lifestyle modification on cognitive function in older adults: An eighteen-month community-based cluster randomized controlled trial. Psychother Psychosom. 2014;83(5):270–8. |
| Lee, 2023 | NA | Lee S, Harada K, Bae S, Harada K, Makino K, Anan Y, et al. A non-pharmacological multidomain intervention of dual-task exercise and social activity affects the cognitive function in community-dwelling older adults with mild to moderate cognitive decline: A randomized controlled trial. Front Aging Neurosci. 2023;15(March):1–10. |
| Lee, 2024 | TIGER | Lee WJ, Peng LN, Lin MH, Kim S, Hsiao FY, Chen LK. Enhancing Intrinsic Capacity and Related Biomarkers in Community-Dwelling Multimorbid Older Adults Through Integrated Multidomain Interventions: Ancillary Findings From the Taiwan Integrated Geriatric (TIGER) Trial. J Am Med Dir Assoc [Internet]. 2024;25(5):757-763.e4. Available from: https://doi.org/10.1016/j.jamda.2023.10.006 |
| Liu, 2023 | COMBAT | Liu X, Ma Z, Zhu X, Zheng Z, Li J, Fu J, et al. Cognitive Benefit of a Multidomain Intervention for Older Adults at Risk of Cognitive Decline: A Cluster-Randomized Controlled Trial. Am J Geriatr Psychiatry [Internet]. 2023;31(3):197–209. Available from: https://doi.org/10.1016/j.jagp.2022.10.006 |
| Maffei, 2017 | Train the Brain | Consortium T the B. Randomized trial on the effects of a combined physical/cognitive training in aged MCI subjects: the Train the Brain study. Sci Rep. 2017;7:39471. |
| Meng, 2024 | NA | Meng X, Su J, Gao T, Ma D, Zhao Y, Fang S, et al. Multidomain interventions based on a life-course model to prevent dementia in at-risk Chinese older adults: A randomized controlled trial. Int J Nurs Stud [Internet]. 2024;152:104701. Available from: https://doi.org/10.1016/j.ijnurstu.2024.104701 |
| Moon, 2025 | SUPERBRAIN-MEET | Moon SY, Park YK, Jeong JH, Hong CH, Jung J, Na HR, et al. South Korean study to prevent cognitive impairment and protect brain health through multidomain interventions via face-to-face and video communication platforms in mild cognitive impairment (SUPERBRAIN-MEET): A randomized controlled trial. Alzheimer's Dement. 2025;21(2):1–13. |
| Moon, 2021 | SUPERBRAIN | Moon SY, Hong CH, Jeong JH, Park YK, Na HR, Song H-S, et al. Facility-based and home-based multidomain interventions including cognitive training, exercise, diet, vascular risk management, and motivation for older adults: a randomized controlled feasibility trial. Aging (Albany NY). 2021 Jun;13(12):15898–916. |
| Murukesu, 2023 | WE-RISE™ | Murukesu R.R., Shahar S., Subramaniam P., Rasdi H.F.M., Nur A.M., Singh D.K.A. The WE-RISE TM Multidomain Intervention : A feasibility study for the potential reversal of cognitive frailty in Malaysian older persons from lower socioeconomic status-pre print. Res Sq. 2023;1–37. |

| First Author, Year | RCT Name | Full Reference |
|---|---|---|
| Ngandu, 2015 | FINGER | Ngandu T, Lehtisalo J, Solomon A, Levälahti E, Ahtiluoto S, Antikainen R, et al. A 2 year multidomain intervention of diet, exercise, cognitive training, and vascular risk monitoring versus control to prevent cognitive decline in at-risk elderly people (FINGER): A randomized controlled trial. Lancet. 2015;385(9984):2255–63. |
| Ng, 2021 | NA | Ng PEM, Nicholas SO, Wee SL, Yau TY, Chan A, Chng I, et al. Implementation and effectiveness of a multi-domain program for older adults at risk of cognitive impairment at neighborhood senior centres. Sci Rep. 2021 Feb;11(1):3787. |
| Ng, 2018 | Singapore Frailty Intervention Trial | Ng TP, Ling LHA, Feng L, Nyunt MSZ, Feng L, Niti M, et al. Cognitive Effects of Multi-Domain Interventions among Pre-Frail and Frail Community-Living Older Persons: Randomized Controlled Trial. Journals Gerontol - Ser A Biol Sci Med Sci. 2018;73(6):806–12. |
| Oki, 2024 | J-MINT PRIME Tamba | Oki Y, Osaki T, Kumagai R, Murata S, Encho H, Ono R, et al. An 18-month multimodal intervention trial for preventing dementia: J-MINT PRIME Tamba. Alzheimer's Dement. 2024;20(10):6972–83. |
| Park, 2019 | NA | Park JE, Jeon SY, Kim SA, Kim JH, Kim SH, Lee KW, et al. A Multidomain Intervention for Modifying Lifestyle Habits Reduces the Dementia Risk in Community-Dwelling Older Adults: A Single-Blinded Randomized Controlled Pilot Study. J Alzheimer's Dis. 2019;70(1):51–60. |
| Piccirilli, 2019 | NA | Piccirilli M, Pigliautile M, Arcelli P, Baratta I, Ferretti S. Improvement in cognitive performance and mood in healthy older adults: a multimodal approach. Eur J Ageing [Internet]. 2019;16(3):327–36. Available from: https://doi.org/10.1007/s10433-019-00503-3 |
| Ponvel, 2025 | AGELESS | Ponvel P, Shahar S, Singh DKA, Ludin AFM, Subramaniam P, Ibrahim N, Haron H, Ismail A, Ai-Vyrn C, Mohamad M, Fadzil H, Khalid NM, Safien AM, Hanipah JM, Ibrahim A, Lehtisalo J, Kivipelto M, Mangialasche F. The cost effectiveness of a multidomain intervention on physical, cognitive, vascular, dietary and psychosocial outcomes among community dwelling older adults with cognitive frailty in Malaysia: The AGELESS Trial. Alzheimers Res Ther. 2025 May 13;17(1):101. doi: 10.1186/s13195-025-01722-w. |
| Richard, 2019 | HATICE | Richard E, Moll van Charante EP, Hoevenaar-Blom MP, Coley N, Barbera M, van der Groep A, et al. Healthy ageing through internet counselling in the elderly (HATICE): a multinational, randomized controlled trial. Lancet Digit Heal [Internet]. 2019;1(8):e424–34. Available from: http://dx.doi.org/10.1016/S2589-7500(19)30153-0 |
| Roach, 2023 | COCA | Roach JC, Rapozo MK, Hara J, Glusman G, Lovejoy J, Shankle WR, et al. A Remotely Coached Multimodal Lifestyle Intervention for Alzheimer's Disease Ameliorates Functional and Cognitive Outcomes. J Alzheimer's Dis. 2023;96(2):591–607. |
| Sakurai, 2024 | J-MINT | Sakurai T, Sugimoto T, Akatsu H, Doi T, Fujiwara Y, Hirakawa A, et al. Japan-Multimodal Intervention Trial for the Prevention of Dementia: A randomized controlled trial. Alzheimer's Dement. 2024;20(6):3918–30. |
| Sugimoto, 2024 | J-MIND-Diabetes | Sugimoto T, Araki A, Fujita H, Fujita K, Honda K, Inagaki N, et al. Multidomain Intervention Trial for Preventing Cognitive Decline among Older Adults with Type 2 Diabetes: J-MIND-Diabetes. J Prev Alzheimer's Dis [Internet]. 2024;11(6):1604–14. Available from: http://dx.doi.org/10.14283/jpad.2024.117 |
| Tainta, 2024 | GOIZ ZAINDU | Tainta M, Ecay-Torres M, de Arriba M, Barandiaran M, Otaegui-Arrazola A, Iriondo A, et al. GOIZ ZAINDU study: a FINGER-like multidomain lifestyle intervention feasibility randomized trial to prevent dementia in Southern Europe. Alzheimers Res Ther. 2024 Feb;16(1):44. |
| Thunborg, 2024 | MIND-AD$_{mini}$ | Thunborg C, Wang R, Rosenberg A, Sindi S, Andersen P, Andrieu S, et al. Integrating a multimodal lifestyle intervention with medical food in prodromal Alzheimer's disease: the MIND-ADmini randomized |

| First Author, Year | RCT Name | Full Reference |
|---|---|---|
| | | controlled trial. Alzheimer's Res Ther [Internet]. 2024;16(1):1–12. Available from: https://doi.org/10.1186/s13195-024-01468-x |
| Van Charante, 2016 | preDIVA | van Charante EPM, Richard E, Eurelings LS, van Dalen JW, Ligthart SA, van Bussel EF, et al. Effectiveness of a 6-year multidomain vascular care intervention to prevent dementia (preDIVA): a cluster-randomized controlled trial. Lancet [Internet]. 2016;388(10046):797–805. Available from: http://dx.doi.org/10.1016/S0140-6736(16)30950-3 |
| Vanoh, 2019 | NA | Vanoh Divya, Shahar Suzana, Razali Rosdinom, Ali Nazlena Mohamad, Manaf Zahara Abdul, Mohd Noah Shahrul Azman, et al. The Effectiveness of a Web-Based Health Education Tool, WESIHAT 2.0, among Older Adults: A Randomized Controlled Trial. J Alzheimer's Dis [Internet]. 2019 Jun 24;70(s1):S255–70. Available from: https://doi.org/10.3233/JAD-180464 |
| Xu, 2025 | NA | Xu Z, Zhang D, Yip BHK, Lee EKP, Poon PKM, Peters R, et al. Combined mind–body physical exercise, cognitive training, and nurse-led risk factor modification to enhance cognition among older adults with mild cognitive impairment in primary care: a three-arm randomized controlled trial. Lancet Heal Longev [Internet]. 2025;6(4):100706. Available from: https://doi.org/10.1016/j.lanhl.2025.100706 |
| Xu, 2020 | NA | Xu Z, Zhang D, Lee ATC, Sit RWS, Wong C, Lee EKP, et al. A pilot feasibility randomized controlled trial on combining mind-body physical exercise, cognitive training, and nurse-led risk factor modification to reduce cognitive decline among older adults with mild cognitive impairment in primary care. PeerJ. 2020;8. |
| Yaffe, 2024 | SMARRT | Yaffe K, Vittinghoff E, Dublin S, Peltz CB, Fleckenstein LE, Rosenberg DE, et al. Effect of Personalized Risk-Reduction Strategies on Cognition and Dementia Risk Profile among Older Adults: The SMARRT Randomized Clinical Trial. JAMA Intern Med. 2024;184(1):54–62. |
| Yang, 2022 | NA | Yang Q hong, Lyu X, Lin Q ran, Wang Z wen, Tang L, Zhao Y, et al. Effects of a multicomponent intervention to slow mild cognitive impairment progression: A randomized controlled trial. Int J Nurs Stud [Internet]. 2022;125:104110. Available from: https://doi.org/10.1016/j.ijnurstu.2021.104110 |
| Zulke, 2024 | AgeWell.de | Zülke AE, Pabst A, Luppa M, Roehr S, Seidling H, Oey A, et al. A multidomain intervention against cognitive decline in an at-risk-population in Germany: Results from the cluster-randomized AgeWell.de trial. Alzheimer's Dement. 2024;20(1):615–28. |

NA: Not available

**Supplementary Table 5.**
**Full-read excluded studies with reason for exclusion**

| First Author, publication year (Country) | DOI | Title | Reason for exclusion (full) | Reason for exclusion (PRISMA flowchart) |
|---|---|---|---|---|
| Hardeman, 2009 (UK) | 10.1186/1479-5868-6-16 | Impact of a physical activity intervention program on cognitive predictors of behavior among adults at risk of Type 2 diabetes (ProActive randomized controlled trial) | Single-domain intervention | Wrong Intervention |
| Beck, 2010 (Denmark) | 10.1016/j.archger.2009.05.018 | Physical and social functional abilities seem to be maintained by a multifaceted randomized controlled nutritional intervention among old (>65 years) Danish nursing home residents | Intervention duration < 6 months | Wrong Intervention |
| Pieramico, 2012 (Italy) | 10.1371/journal.pone.0043901 | Combination Training in Aging Individuals Modifies Functional Connectivity and Cognition, and Is Potentially Affected by Dopamine-Related Genes | Two-domain Intervention | Wrong Intervention |
| Ng, 2014 (Singapore) | N/A | Effects of nutritional, physical, cognitive interventions on cognitive outcomes in the Singapore frailty intervention trial (S-FIT) | Abstract of a study already included | Conference abstract |
| Dannhauser, 2014 (UK) | 10.1186/1471-244X-14-129 | A complex multimodal activity intervention to reduce the risk of dementia in mild cognitive impairment-ThinkingFit: Pilot and feasibility study for a randomized controlled trial | Not randomized, intervention < 6 months, two-domain intervention | Wrong study design |
| Ihle-Hansen, 2014 (Norway) | 10.1111/j.1747-4949.2012.00928.x | Multifactorial vascular risk factor intervention to prevent cognitive impairment after stroke and TIA: A 12-month randomized controlled trial | Targeted to post-stroke participants | Wrong population |
| Kim, 2015 (South Korea) | N/A | Effects of multimodal cognitive enhancement therapy (MCET) for people with mild cognitive impairment and early-stage dementia: a randomized, controlled, double-blind, cross-over trial | Abstract of a study already included | Conference abstract |
| Diamond, 2015 (Australia) | 10.3233/JAD-142061 | Randomized controlled trial of a healthy brain ageing cognitive training program: effects on memory, mood, and sleep | Intervention duration < 6 months | Wrong Intervention |
| Santos, 2015 (Brazil) | 10.1590/0101-60830000000066 | Multidisciplinary rehabilitation program: Effects of a multimodal intervention for patients with Alzheimer's disease and cognitive impairment without dementia | Intervention duration < 6 months, including participants with moderate AD | Wrong Intervention |
| Matz, 2015 (Austria) | 10.1161/STROKEAHA.115.009992 | Multidomain lifestyle interventions for the prevention of cognitive decline after ischemic stroke randomized trial | Targeted to post-stroke participants | Wrong population |

| First Author, publication year (Country) | DOI | Title | Reason for exclusion (full) | Reason for exclusion (PRISMA flowchart) |
|---|---|---|---|---|
| Dominguez (Philippines), 2017 | N/A | A community-based ballroom dance intervention "INDAK" improved cognition among elderly with mild cognitive impairment | Abstract and intervention consisting of dance only | Conference abstract |
| Bruno (Italy), 2017 | 10.1097/01.hjh.0000522988.06828.b9 | Effect of a multimodality training on cognitive and vascular function in MCI patients with or without hypertension: the train the brain-mind the vessel study | Abstract of a study already included | Conference abstract |
| Rosli, 2017(Malaysia) | N/A | Effects of multicomponent exercise and therapeutic lifestyle (CERgAS) intervention on cognitive function in lower income elderly population: a cluster randomized controlled trial | Abstract | Conference abstract |
| Seino, 2017(Japan) | https://dx.doi.org/10.1111/ggi.13016 | Effects of a multifactorial intervention comprising resistance exercise, nutritional and psychosocial programs on frailty and functional health in community-dwelling older adults: A randomized, controlled, cross-over trial. | Intervention duration < 6 months | Wrong Intervention |
| Strandberg, 2017 (Finland) | 10.1016/j.eurger.2016.12.005 | Health-related quality of life in a multidomain intervention trial to prevent cognitive decline (FINGER) | Cognition not included | Wrong outcome |
| Streber (Germany), 2017 | 10.2147/CIA.S141163 | A multicenter controlled study for dementia prevention through physical, cognitive and social activities - GESTALT-kompakt | Intervention duration < 6 months | Wrong Intervention |
| Teuschl, 2017 (Austria) | 10.1177/1747493017702662 | Preventive effects of multiple domain interventions on lifestyle and risk factor changes in stroke survivors: Evidence from a two-year randomized trial | Adherence analysis cognition not included as outcome | Wrong outcome |
| Hayden, 2018 (USA) | 10.14283/jpad.2018.40 | The action for health in diabetes clinical trial: does a 10-year intensive multidomain lifestyle intervention provide cognitive benefits? | Abstract | Conference abstract |
| Keine, 2018(USA) | 10.2174/1874609811666181019101430 | Development, application, and results from a precision-medicine platform that personalizes multi-modal treatment plans for mild Alzheimer's disease and at-risk individuals | Methodology paper | Wrong study design |
| Romera-Liebana, 2018 (Spain) | 10.1093/gerona/glx259 | Effects of a Primary Care-Based Multifactorial Intervention on Physical and Cognitive Function in Frail, Elderly Individuals: A Randomized Controlled Trial | Intervention duration < 6 months | Wrong Intervention |
| Tabue-Teguo, 2018 (France) | 10.1007/s12603-018-1024-6 | Effect of Multidomain Intervention, Omega-3 Polyunsaturated Fatty Acids Supplementation or their Combinaison on Cognitive Function in Non-Demented Older Adults According to Frail Status: Results from the MAPT Study | Subgroup analysis of a study already included | Wrong study design |

| First Author, publication year (Country) | DOI | Title | Reason for exclusion (full) | Reason for exclusion (PRISMA flowchart) |
|---|---|---|---|---|
| Li, 2018 (China) | 10.1097/MD.0000000000011975 | Effect of long-term lifestyle intervention on mild cognitive impairment in hypertensive occupational population in China | Multidomain vs control comparison not randomized | Wrong study design |
| Bruno, 2018 (Italy) | 10.1161/HYPERTENSIONAHA.117.10066 | Vascular function is improved after an environmental enrichment program, the train the brain-mind the vessel study | Post-hoc secondary analysis of a study already included | Wrong study design |
| Chandler, 2019 (USA) | 10.1001/jamanetworkopen.2019.3016 | Comparative effectiveness of behavioral interventions on quality of life for older adults with mild cognitive impairment, a randomized Clinical Trial | Intervention duration < 6 months | Wrong Intervention |
| Kropacova, 2019 (Czech Republic) | 10.1007/s00702-019-02068-y | Cognitive effects of dance-movement intervention in a mixed group of seniors are not dependent on hippocampal atrophy | Intervention including dance only two-domain at most | Wrong Intervention |
| Isaacson, 2019 (USA) | 10.1016/j.jalz.2019.08.198 | Individualized clinical management of patients at risk for Alzheimer's dementia | Not RCT | Wrong study design |
| Anstey, 2020 (Australia) | 10.2196/19431 | An internet-based intervention augmented with a diet and physical activity consultation to decrease the risk of dementia in at-risk adults in a primary care setting: Pragmatic randomized controlled trial | Intervention duration < 6 months | Wrong Intervention |
| Falck, 2020 (Canada) | 10.3233/JAD-200383 | Effect of a Multimodal Lifestyle Intervention on Sleep and Cognitive Function in Older Adults with Probable Mild Cognitive Impairment and Poor Sleep: A Randomized Clinical Trial | Addressing only sleep | Wrong intervention |
| McMaster, 2020 (Australia) | 10.1111/jgs.16762 | Lifestyle Risk Factors and Cognitive Outcomes from the Multidomain Dementia Risk Reduction Randomized Controlled Trial, Body Brain Life for Cognitive Decline (BBL-CD) | Intervention duration < 6 months | Wrong Intervention |
| Hayden, 2021 (USA) | 10.1159/000517160 | Legacy of a 10-year multidomain lifestyle intervention on the cognitive trajectories of individuals with overweight/obesity and type 2 diabetes mellitus | Intervention specifically aimed at diabetes | Wrong setting |
| Lee, 2021 (Taiwan) | 10.1016/S2666-7568(21)00248-8 | Effects of incorporating multidomain interventions into integrated primary care on quality of life: a randomized controlled trial. | Intervention effect on quality of life, cognition not included as outcome | Wrong outcome |
| Liang, 2021 (Taiwan) | 10.1016/j.archger.2021.104392 | Efficacy of Multidomain Intervention Against Physio-cognitive Decline Syndrome: A Cluster-randomized Trial | Subgroup analysis from a study | Wrong study design |

| First Author, publication year (Country) | DOI | Title | Reason for exclusion (full) | Reason for exclusion (PRISMA flowchart) |
|---|---|---|---|---|
| | | | already included | |
| Liou, 2021 (USA) | 10.3390/brainsci11101306 | Compensatory and lifestyle-based brain health program for subjective cognitive decline: Self-implementation versus coaching | Intervention duration < 6 months | Wrong Intervention |
| Meeuwsen, 2021 (USA) | 10.3233/RNN-201053 | Z-score neurofeedback, heart rate variability biofeedback, and brain coaching for older adults with memory concerns | Intervention duration < 6 months | Wrong Intervention |
| Ruiz-Tagle, 2021(Chile) | 10.4067/S0034-98872021001101569 | Effects of a multidimensional intervention in older adults with mild cognitive impairment | Intervention duration < 6 months | Wrong Intervention |
| Chew, 2021 (Singapore) | 10.1002/trc2.12141 | SINgapore GERiatric intervention study to reduce physical frailty and cognitive decline (SINGER)–pilot: A feasibility study | FINGER intervention as control | Wrong Comparator |
| Han, 2022 (South Korea) | 10.4017/gt.2022.21.s.789.4.sp3 | SUPERBRAIN (the south Korean study to prevent cognitive impairment and protect BRAIN health through lifestyle intervention in at-risk elderly people.): a randomized controlled feasibility trial | Abstract of a study already included | Conference abstract |
| Belleville, 2022 (France) | 10.1002/alz.12544 | Is more always better? Dose effect in a multidomain intervention in older adults at risk of dementia | Post-hoc adherence dose-response analysis from a study already included | Wrong study design |
| Brasser, 2022 (Switzerland) | 10.3389/fpsyg.2022.866613 | A Randomized Controlled Trial Study of a Multimodal Intervention vs. Cognitive Training to Foster Cognitive and Affective Health in Older Adults | Intervention duration < 6 months | Wrong Intervention |
| Ihl, 2022 (Germany, Denmark) | https://dx.doi.org/10.1161/STROKEAHA.120.037503 | Patient-Centered Outcomes in a Randomized Trial Investigating a Multimodal Prevention Program After Transient Ischemic Attack or Minor Stroke: The INSPiRE-TMS Trial. | Secondary stroke prevention study | Wrong setting |
| Jung, 2022 (South Korea) | 10.3390/ijerph192114339 | Effects of Integrative Cognitive Function Improvement Program on Cognitive Function, Oral Health, and Mental Health in Older People: A Randomized Clinical Trial | Intervention duration < 6 months | Wrong Intervention |
| Lee, 2022 (Taiwan) | 10.1007/s12603-022-1843-3 | Clinical Efficacy of Multidomain Interventions among Multimorbid Older People Stratified by the Status of Physio-Cognitive Declines: A Secondary Analysis from the Randomized Controlled Trial for Healthy Aging | Secondary analysis of a study already included | Wrong study design |

| First Author, publication year (Country) | DOI | Title | Reason for exclusion (full) | Reason for exclusion (PRISMA flowchart) |
|---|---|---|---|---|
| Moon, 2022 (South Korea) | 10.3389/fnagi.2022.926077 | Impact of a multidomain lifestyle intervention on regional spontaneous brain activity | Neuroimaging sub study of a trial already included | Wrong study design |
| Sok, 2022 (South Korea) | 10.3390/ijerph191912113 | Effects of Multicomponent Oriental Integrative Intervention on Cognitive Function, Health Status, Life Satisfaction, and Yangsaeng of Community-Dwelling Elderly | Intervention duration < 6 months | Wrong Intervention |
| Zhu, 2022 (China) | 10.1016/j.jagp.2022.01.011 | A Multimodal Intervention to Improve Cognition in Community-dwelling Older Adults | Two-domain intervention, duration 6 weeks | Wrong Intervention |
| Oki, 2023 (Japan) | 10.14283/jpad.2022.129 | Effects of an 18-month multimodal intervention on cognitive function (J-MINT PRIME TAMBA): a randomized controlled trial. | Abstract of a study already included | Conference abstract |
| Zulke, 2023 (Germany) | 10.1055/s-0043-1770499 | Effectiveness of a multicomponent lifestyle intervention against cognitive decline and dementia in an at-risk-population in Germany - Preliminary results from the cluster-randomized AgeWell.de-trial | Abstract of a study already included | Conference abstract |
| Belleville, 2023 (Canada) | 10.1007/s11357-022-00674-5 | Pre-frail older adults show improved cognition with StayFitLonger computerized home-based training: a randomized controlled trial | 2 domains (Physical and cognitive Training) | Wrong Intervention |
| Chang, 2023 (Taiwan) | 10.5014/ajot.2023.050133 | A Multicomponent Cognitive Intervention May Improve Self-Reported Daily Function of Adults With Subjective Cognitive Decline | Single-arm two-period crossover trial; 16-week intervention | Wrong study design |
| Eunyoung, 2023 (South Korea) | 10.1016/j.gerinurse.2023.01.001 | The effect of a multimodal intervention program on cognitive and daily function of older persons residing in rural communities: A pilot study | Not randomized | Wrong study design |
| Kim, 2023 (South Korea) | 10.1016/j.pmedr.2023.102165 | The effects of a mobile-based multi-domain intervention on cognitive function among older adults | Intervention duration < 6 months | Wrong Intervention |
| Lee, 2023 (South Korea) | 10.3233/JAD-221299 | Efficacy of a Mobile-Based Multidomain Intervention to Improve Cognitive Function and Health-Related Outcomes among Older Korean Adults with Subjective Cognitive Decline | Intervention duration < 6 months | Wrong Intervention |
| Lee, 2023 (South Korea) | 10.3389/fnagi.2023.1266955 | Effects of the multidomain intervention with nutritional supplements on cognition and gut microbiome in early symptomatic Alzheimer's disease: a randomized controlled trial | Intervention duration < 6 months | Wrong Intervention |
| McMaster, 2023 (Australia) | 10.1080/13607863.20 | The feasibility of a multidomain dementia risk reduction randomized controlled trial for people experiencing | Intervention duration < 6 months | Wrong Intervention |

| First Author, publication year (Country) | DOI | Title | Reason for exclusion (full) | Reason for exclusion (PRISMA flowchart) |
|---|---|---|---|---|
| | 23.2190083 | cognitive decline: the Body, Brain, Life for Cognitive Decline (BBL-CD) | | |
| Montero-Odasso, 2023 (Canada) | 10.1001/jamanetworkopen.2023.24465 | Effects of Exercise Alone or Combined with Cognitive Training and Vitamin D Supplementation to Improve Cognition in Adults with Mild Cognitive Impairment: A Randomized Clinical Trial | Intervention duration < 6 months | Wrong Intervention |
| Novikova, 2023 (Russia) | 10.14412/2074-2711-2023-1-57-64 | Efficacy of a combination of non-drug therapies in patients with non-dementia vascular cognitive impairment | Intervention duration < 6 months | Wrong Intervention |
| Oh, 2023(South Korea) | 10.3233/NRE-220253 | Self-perception and anticipated efficacy of the anti-dementia multimodal program in 100 older adults with mild cognitive impairment | Not multidomain intervention | Wrong Intervention |
| Wu, 2023 (Taiwan) | 10.3390/nu15081976 | Efficacy of Dietary Intervention with Group Activities on Dietary Intakes, Frailty Status, and Working Memory: A Cluster-Randomized Controlled Trial in Community Strongholds | Intervention duration < 6 months | Wrong Intervention |
| Zhang, 2023(China) | 10.3389/fpubh.2023.1088833 | Incidence of cognitive impairment after hypothetical interventions on depression, nighttime sleep duration, and leisure activity engagement among older Chinese adults: An application of the parametric g-formula | Simulation of RCT from observational data | Wrong study design |
| Kim, 2024 (USA) | 10.1016/j.puhip.2024.100528 | The efficacy of a mobile-based multidomain program on cognitive functioning of residents in assisted living facilities | Intervention duration < 6 months | Wrong Intervention |
| Ma, 2024 (China) | 10.2139/ssrn.5073208 | Combination of Music-Based Movement, Multi-Domain Cognitive Training Synchronized with Neuromodulation, and Digital Emotion Regulation Education (MMNCE) on Cognition in Older Adults with Normal Cognition: a Randomized Controlled Trial | Intervention duration < 6 months | Wrong Intervention |
| Chen, 2024 (China) | 10.1080/01634372.2024.2355152 | The Effects of a Nonpharmacological Intervention Practice for Older Adults with Mild Cognitive Impairment and Their Family Caregivers in China | Non-randomized, single-domain intervention | Wrong study design |
| Chew, 2024 (Singapore) | https://dx.doi.org/10.3390/ijerph22010042 | ADL+: A Digital Toolkit for Multidomain Cognitive, Physical, and Nutritional Interventions to Prevent Cognitive Decline in Community-Dwelling Older Adults. | Longitudinal, quasi-experimental study, no RCT | Wrong study design |
| Fan, 2024(China) | 10.3233/JAD-231370 | Effect of Multimodal Intervention in Individuals with Mild Cognitive Impairment: A Randomized Clinical Trial in Shanghai | Intervention duration < 6 months | Wrong Intervention |

| First Author, publication year (Country) | DOI | Title | Reason for exclusion (full) | Reason for exclusion (PRISMA flowchart) |
|---|---|---|---|---|
| Garcia-Llorente, 2024 (Spain) | 10.1007/s40520-024-02700-2 | Multidomain interventions for sarcopenia and cognitive flexibility in older adults for promoting healthy aging: a systematic review and meta-analysis of randomized controlled trials | Systematic review and meta-analysis | Wrong study design |
| Hsieh, 2024 (Taiwan) | https://dx.doi.org/10.1177/15333175241256803 | Effectiveness of Early Multimodal Non-pharmacological Interventions in Cognitive Preservation in the Elderly. | Not randomized, intervention < 6 months, comparing non dementia vs dementia people | Wrong study design |
| Jennings, 2024 (UK) | https://dx.doi.org/10.1186/s12916-024-03815-z | Effectiveness and feasibility of a theory-informed intervention to improve Mediterranean diet adherence, physical activity and cognition in older adults at risk of dementia: the MedEx-UK randomized controlled trial. | Two-domain Intervention | Wrong intervention |
| Keawtep, 2024 (Thailand) | 10.1186/s12966-024-01580-z | Effects of combined dietary intervention and physical-cognitive exercise on cognitive function and cardiometabolic health of postmenopausal women with obesity: a randomized controlled trial | Intervention duration < 6 months | Wrong Intervention |
| Kunkler, 2024 (Germany) | 10.3233/ADR-230199 | Long-Term Effects of the Multicomponent Program BrainProtect® on Cognitive Function: One-Year Follow-Up in Healthy Adults | Intervention duration < 6 months and single domain | Wrong Intervention |
| Nakada, 2024 (Japan) | https://dx.doi.org/10.3390/healthcare12232365 | A Real-Time Web-Based Intervention with a Multicomponent Group-Based Program for Older Adults: Single-Arm Feasibility Study. | Intervention duration < 6 months | Wrong Intervention |
| Ornish, 2024 (USA) | 10.1186/s13195-024-01482-z | Effects of intensive lifestyle changes on the progression of mild cognitive impairment or early dementia due to Alzheimer's disease: a randomized, controlled clinical trial | Intervention duration < 6 months | Wrong Intervention |
| Sandison, 2024 (USA) | 10.3233/JAD-230004 | Observed Improvement in Cognition During a Personalized Lifestyle Intervention in People with Cognitive Decline | A protocol-driven, uncontrolled, pragmatic trial (No RCT) | Wrong study design |
| Siette, 2024(Australia) | 10.14283/jpad.2024.104 | A Pilot Study of BRAIN BOOTCAMP, a Low-Intensity Intervention on Diet, Exercise, Cognitive Activity, and Social Interaction to Improve Older Adults' Dementia Risk Scores | Intervention duration < 6 months | Wrong Intervention |
| Wang, 2024 (China) | 10.4088/JCP.23m15112 | Effects of a Multicomponent Intervention With Cognitive Training and Lifestyle Guidance for Older Adults at | Intervention duration < 6 months | Wrong Intervention |

| First Author, publication year (Country) | DOI | Title | Reason for exclusion (full) | Reason for exclusion (PRISMA flowchart) |
|---|---|---|---|---|
| | | Risk of Dementia: A Randomized Controlled Trial | | |
| Young, 2024 (China) | 10.1080/01634372.2024.2338066 | Multicomponent Intervention on Improving the Cognitive Ability of Older Adults with Mild Cognitive Impairment: A Pilot Randomized Controlled Trial | Intervention duration < 6 months | Wrong Intervention |
| Zülke, 2024 (Germany) | 10.1002/alz.14097 | Effects of a multidomain intervention against cognitive decline on dementia risk profiles - Results from the AgeWell.de trial | Intervention duration < 6 months | Wrong outcome |
| Liang, 2024 (Taiwan) | 10.14283/jpad.2024.20 | Roles of Baseline Intrinsic Capacity and its Subdomains on the Overall Efficacy of Multidomain Intervention in Promoting Healthy Aging among Community-Dwelling Older Adults: Analysis from a Nationwide Cluster-Randomized Controlled Trial | Post-hoc analysis of a study already included | Wrong study design |
| Forcano, 2025 (Spain) | https://doi.org/10.1016/j.tjpad.2025.100271 | A multimodal lifestyle intervention complemented with epigallocatechin gallate to prevent cognitive decline in APOE- ε4 carriers with Subjective Cognitive Decline: a randomized, double-blinded clinical trial (PENSA study) | Multidomain Intervention vs Lifestyle control not randomized | Wrong study design |
| Kuroda, 2025 (Japan) | https://dx.doi.org/10.1177/13872877251315042 | Evaluating the feasibility of a community-adapted multi-domain intervention for dementia prevention in older adults. | No RCT one arm only | Wrong study design |
| Lin, 2025 (China) | https://dx.doi.org/10.1038/s41598-025-92535-2 | Development and usability of the "Cognitive Evergreenland" app to engage individuals at high risk of dementia in lifestyle interventions. | No RCT app development study | Wrong study design |
| Lin, 2025 (China) | https://dx.doi.org/10.1186/s12877-025-05684-4 | A mobile-based multidomain lifestyle intervention using Cognitive Evergreenland for older adults with subjective cognitive decline: a feasibility study. | Open-label, single-group, self-controlled, No RCT | Wrong study design |
| Ong, 2025 (Malaysia) | https://dx.doi.org/10.1186/s12889-024-20704-5 | A qualitative study on the impact and participation in the AGELESS multidomain intervention: Insights from older adults with cognitive frailty and their caregivers. | Qualitative study | Wrong study design |
| Osaki, 2025 (Japan) | https://dx.doi.org/10.1177/13872877251332647 | Longitudinal deterioration of subjective cognitive decline in apolipoprotein epsilon4 carriers and improvement of subjective cognitive decline by multi-domain intervention for prevention of dementia: The cognitive function instrument assessment. | Sub-study of a study already included | Wrong setting |
| Siette, 2025 (Australia) | https://dx.doi.org/10.1 | Acceptability and fidelity of the multidomain 'Brain Bootcamp' dementia | Mixed methods | Wrong study design |

| First Author, publication year (Country) | DOI | Title | Reason for exclusion (full) | Reason for exclusion (PRISMA flowchart) |
|---|---|---|---|---|
| | 186/s12889-025-21641-7 | risk reduction program: a mixed-methods approach. | study on acceptability | |
| Sanchez-Arenas, 2025 (Mexico) | https://dx.doi.org/10.5281/zenodo.14200027 | Multi-domain intervention program on cognitive function in community-dwelling older adults: Pilot study. | Not randomized | Wrong study design |

**Supplementary Table 6.**
**Detailed description of the included RCTs' multidomain interventions (full study references provided eTable 4)**

| **RCT** | | **MULTIDOMAIN INTERVENTION** | | | | | |
|---|---|---|---|---|---|---|---|
| **First author, year Country** | **RCT name** | **Intervention Duration (Months)** | **Multidomain Intervention components/Risk factors targeted** | **Mean of delivery and activity types** | **Delivered by** | **Brief description of activity frequency and duration** | **Control** |
| Baker, 2025<br>USA | US POINTER | 24 | Physical activity<br>Diet<br>Cognitive training<br>Vascular risk monitoring<br>Social engagement | In person and remote;<br>Individual and group activities | Navigators (trained but without previous expertise) and interventionists (trained and with intervention relevant expertise) | 4x physical activity group meetings; 4x/week Aerobic (30-35 min/session), resistance (2x/week, 15-20 min/session), and flexibility (2x/week, 10-15 min/session) exercise<br>4x diet group team meetings; regular phone consultations with diet interventionist<br>4x cognition group meetings; independent computerised cognitive training (3x/week, 15-20 min/session)<br>4x vascular risk factors group meetings; biannual individual consultation with review of abnormal laboratory results and reinforcement of intervention goals | General/untailored health advice (publicly available education material; 6 times during the study) |
| Ide, 2025<br>Japan | J-MINT PRIME Kanagawa | 18 | Physical exercise<br>Diet<br>Cognitive training<br>Managements of lifestyle-related diseases<br>Oral hygiene | In person and remote;<br>Individual and groups activities | Public health nurses, dietitians, and other trained professionals | 2X/week (90 min) aerobic, dual-task, and strength exercise<br>3X (60 min) diet consultations (M1, M7, M13);<br>1x/month (10 min) diet follow-up phone call<br>≥ 4x/week (30 min) computerised cognitive training<br>Recommendations to seek the family doctor's advice based on specific needs at baseline and follow-up visits<br>Oral hygiene recommendations provided at baseline and follow-up visits | General/untailored health advice (every 6 months) |

| RCT | | MULTIDOMAIN INTERVENTION | | | | | |
|---|---|---|---|---|---|---|---|
| **First author, year Country** | **RCT name** | **Intervention Duration (Months)** | **Multidomain Intervention components/Risk factors targeted** | **Mean of delivery and activity types** | **Delivered by** | **Brief description of activity frequency and duration** | **Control** |
| Brodaty, 2025 Australia | Maintain Your Brain (MYB) | 36 | Physical activity<br>Diet<br>Cognitive training<br>Mental health (stress/anxiety management) | Remote (Online secure internet-based digital platform); Individual activities | Unsupervised via the digital platform - remotely available study team for technical support and lifestyle changes advice | Individually tailored feedback and goals for progressive aerobic exercise (300 min7week - moderate intensity or 150 min/week -vigorous intensity), strength (≥3 days/week - vigorous intensity) and balance (1x/day)<br>Recommendations for "mediterranean diet pattern"+ food advice (tips on cooking, recipes, cooking videos, shopping lists, and fact sheets)<br>30x cognitive training sessions (3x/week, 45 min) for the first 10 weeks, 1x/month afterwards<br>6 illustrated digital mental health program modules over 10 weeks | General/untailored health advice (Access to a less personalised non interactive digital platform) |
| Moon, 2025 South Korea | SUPERBRAIN-MEET | 6 | Physical activity<br>Diet<br>Cognitive training,<br>Cardiometabolic risk monitoring<br>Motivational enhancement | In person and remote; Individual and groups activities | Psychologists, occupational therapists, study nurses, professional trainers, study doctor | 3x/week (50 min) physical activities<br>12 diet group sessions<br>every 8 weeks;6 individual sessions<br>1x/week computerised cognitive training sessions independently (30-40 min) and in group (50 min 1x/week/2weeks)<br>4 motivational enhancement group sessions | General/untailored health advice and standard care |
| Ponvel, 2025 Malaysia | AGELESS | 24 | Physical activity<br>Diet<br>Cognitive training,<br>Cardiometabolic risk monitoring<br>Psychosocial intervention | In person; Individual and groups activities | Dietitian, psychologist, nurse, physician, physiotherapist | 2-3x/week individually tailored balance, coordination, aerobic, resistance and flexibility exercise.<br>1x/month (first 12months) individual diet consultations; group education sessions on food intake, nutrients and healthy eating<br>2-3x/week individually tailored "paper and pen" cognitive training<br>1x/6months individual consultation on cardiometabolic risk<br>1x/month individual consultation on mental wellbeing; group sessions on mental health, coping and visiting | Standard care, General/untailored health advice and 5x group counselling sessions |

| RCT | | MULTIDOMAIN INTERVENTION | | | | | |
|---|---|---|---|---|---|---|---|
| **First author, year Country** | **RCT name** | **Intervention Duration (Months)** | **Multidomain Intervention components/Risk factors targeted** | **Mean of delivery and activity types** | **Delivered by** | **Brief description of activity frequency and duration** | **Control** |
| Xu, 2025 Hong Kong | - | 15 | Physical exercise<br>Cognitive training,<br>Vascular and lifestyle risk monitoring | In person; Individual and groups activities | Trained research assistant, study primary care clinicians, study nurse, physical therapist | 3x/week (12 weeks, 30 min/session) Tai chi<br>3x/week (12 weeks, 30 min/session) leisure cognitive activities (board games, reading, mahjong etc.)<br>1x/3 months (30 min) risk monitoring consultations | General/untailored health advice (at baseline and via educational material) |
| Lee, 2024 Taiwan | TIGER | 12 | Physical activity<br>Diet<br>Cognitive training<br>Healthy aging and management of chronic condition | In person; Groups activities | Physicians, research nurse, | 16 structured 2-hour sessions (initially weekly and then gradual decrease in frequency) with multidomain content (45 min physical activity, 1h cognitive training, 15 min diet) and educational sessions on healthy ageing | General/untailored health advice and Standard care |
| Meng, 2024 China | NA | 6 | Dementia literacy<br>Physical activity<br>Diet<br>Cognitive training<br>Social engagement<br>Tobacco cessation<br>Depression management<br>Diabetes management | In person and remote; Individual and groups activities | Community worker (guided and supervised by a researcher) | Online component providing educational content and chat group covering all intervention components for dementia literacy.<br>4 articles + 4 audios (online platform) and 1 in-person session for physical activity<br>5 articles (online platform) on diet<br>4 article + 5 audios + 12-week cognitive training (1x/week)<br>7 articles + 6 audios on social activities (online platform) and 1 group in-person activity + 6 voluntary social activities | General/untailored health advice (Dementia literacy module and WeChat article weekly for 5 weeks) |

| RCT | | MULTIDOMAIN INTERVENTION | | | | | |
|---|---|---|---|---|---|---|---|
| **First author, year Country** | **RCT name** | **Intervention Duration (Months)** | **Multidomain Intervention components/Risk factors targeted** | **Mean of delivery and activity types** | **Delivered by** | **Brief description of activity frequency and duration** | **Control** |
| | | | COVID-19 prevention, | | | Chat group messages from study team on tobacco cessation<br>2 articles (online platform) on depression management<br>Chat group messages from study team on diabetes management<br>2 articles (online platform) on COVID-19 prevention | |
| Murukesu, 2024, Malaysia | WE-RISE™ | 6 | Physical activity<br>Diet<br>Cognitive training<br>Psychosocial support | In person and remote;<br>Individual and groups activities | Physiotherapist, psychologist, dietitian | 2x/week (90 min/session) physical activity and cognitive training combined (24 total sessions); exercise group sessions include flexibility/balance, aerobic exercise, and strength training (increasing intensity) + home-based intervention using a written activity manual/exercise instruction, a ball, and a pair of 2 kg weight cuffs; the cognitive training include different pen-and-paper games/activities/challenges/worksheets, e.g., jigsaw puzzle, memory cards, sorting game (increasing difficulty) + home-based intervention included worksheets of cognitive training activities and jigsaw puzzles<br>1diet group session including distribution of written material + home-based intervention including written dietary guidelines/material<br>Psychosocial support provided during group sessions + home-based motivational phone call every 2 weeks during the second half of the intervention | Standard care as per senior activity centres plans |

| RCT | | MULTIDOMAIN INTERVENTION | | | | | |
|---|---|---|---|---|---|---|---|
| **First author, year Country** | **RCT name** | **Intervention Duration (Months)** | **Multidomain Intervention components/Risk factors targeted** | **Mean of delivery and activity types** | **Delivered by** | **Brief description of activity frequency and duration** | **Control** |
| Oki, 2024, Japan | J-MINT PRIME Tamba | 18 | Physical activity<br>Diet<br>Cognitive training<br>Lifestyle-related disease management | In person;<br>Individual and groups activities | Physical therapists, occupational therapists, nurses, dieticians | In-person consultation for lifestyle-related disease management based on individual needs<br>1x/week aerobic (50 min), dual task/resistance (20 min) exercise + 20 min group session<br>3x diet counselling (M1, M7, M13) and telephone follow-ups after each appointment<br>≥4x/week (30 min/session) computerised cognitive training (practice and rest periods every 3 months) | General/untailored health advice (every 2 months) |
| Sakurai, 2024 Japan | J-MINT | 18 | Physical activity<br>Diet<br>Cognitive training<br>CVD treatment | In person and remote;<br>Individual and groups activities (online programmes used during the COVID-19 to replace in-person meetings) | Qualified health consultant for diet, study physician | 1x/week group-based physical exercise sessions (90 min 78 sessions in total)<br>3x in person and 12x telephone diet consultations sessions<br>Treatment for diabetes, hypertension, and dyslipidaemia based on relevant clinical practice guidelines<br>≥4x/week (30 min/session) computerised cognitive training<br>during the 4th to 6th, 10th to 12th, and 16th to 18th months (intensive training periods) | Standard care<br>General/untailored health advice (bi-monthly) |
| Sugimoto, 2024 Japan | J-MIND-Diabetes | 18 | Physical activity<br>Diet<br>Cardio-metabolic risk monitoring<br>Social engagement | In person and remote;<br>Individual and groups activities | Physiotherapists or trained instructors, dieticians, trained psychologist, study physician | ≥1x/2 weeks group based physical exercise sessions (total 39 sessions including muscle strength, postural balance training, aerobic exercise, and dual-task training); during the COVID-19 outbreak, ≥ 2x/week home-based sessions<br>Individual diet counselling 1x/2 months<br>Individual counselling for cardio-metabolic risk 1x/2 months<br>Recommendation to engage in social activities outside of home ≥3x/week (activities monitored) | Standard care, General/untailored health advice |

| RCT | | MULTIDOMAIN INTERVENTION | | | | | |
|---|---|---|---|---|---|---|---|
| **First author, year Country** | **RCT name** | **Interventio n Duration (Months)** | **Multidomain Intervention components/Risk factors targeted** | **Mean of delivery and activity types** | **Delivered by** | **Brief description of activity frequency and duration** | **Control** |
| Tainta, 2024 Spain | GOIZ ZAINDU | 12 | Physical activity<br>Diet<br>Cognitive training<br>Cardio-metabolic risk monitoring<br>Social engagement | In person and remote;<br>Individual and groups activities | GP, nurses, nutritionists, psychologists , neuropsychol ogist, personnel from the municipal sports centre | 2x/week indoor group exercise sessions (first 9 months) + recommendations to practice 2–6h/week of aerobic exercise<br>1x/3 months individual diet consultations + 2 workshops (M1; M7)<br>13 cognitive group sessions (90 min/session, 20 h in total) and individual computerised cognitive training 3x/week for 10 months (40 h in total)<br>1x/3 months individual cardio-metabolic risk consultations | Standard care, General/untailored health advice |
| Thunborg, 2024 Sweden, Finland, Germany, France | MIND-ADmini | 6 | Physical activity<br>Diet<br>Cognitive training<br>Cardio-metabolic risk monitoring<br>Social engagement<br>Medical food | In person;<br>Individual and groups activities | Nurses, physicians, nutritionists/d ieticians, physiotherapi st/personal trainer, occupational therapists, psychologists | 2x/week exercise (1h/session; strength training and aerobic exercise, tailored for each participant's fitness level, progressive programme)<br>3x individual diet consultations; 3-4x diet group meetings (60-75min per session)<br>2–3 cognitive group sessions (60-75min per session); individual 2x/week computerised cognitive training (increasing level of difficulty)<br>1x nurse consultation for cardio-metabolic risk<br>1x/day Fortasyn Connect (Souvenaid™, 125 ml) medical food drink | General/untailored health advice |
| Yaffe, 2024 USA | SMARRT | 24 | Multiple risk factors addressed (hypertension; diabetes; depressive symptoms; sleep; risky medications; physical inactivity; social isolation; current smoking) | In person (prior the COVID-19 outbreak) and remote;<br>Individual activities | health coach and nurse | Health coaching sessions 1x/4-6 weeks to set goals related to risk factors chosen by participants; more frequent visits in the first 3months, then 1x/6 weeks for the final 15 months<br>Sessions lasted longer (45 min) in the first 3months, then reduced to 20 minutes for the remainder of the calls<br>Participants were provided with educational material on selected risk factors, self-monitoring tools, and resources (e.g., fitness tracker, sleep | Standard care and General/untailored health advice (every 3 months) |

| RCT | | MULTIDOMAIN INTERVENTION | | | | | |
|---|---|---|---|---|---|---|---|
| **First author, year Country** | **RCT name** | **Intervention Duration (Months)** | **Multidomain Intervention components/Risk factors targeted** | **Mean of delivery and activity types** | **Delivered by** | **Brief description of activity frequency and duration** | **Control** |
| | | | based on the specific risk profile of each participant | | | workbook, brain games), depending on preference<br>Participants with uncontrolled diabetes, uncontrolled hypertension, or risky medication use received 1x/3 months contacts from the intervention nurse (session duration as above) | |
| Zulke, 2024 Germany | AgeWell.de | 24 | Physical activity<br>Diet<br>Cognitive training<br>Vascular risk monitoring<br>Social engagement | In person and remote;<br>Individual activities | Nurse, GP | 2x/week at-home standardised exercises for strength and flexibility/balance; 3-5x/week aerobic exercise<br>Dietary recommendations based on the guidelines of German Nutrition Society, e.g., consumption of five portions of fruit and vegetables a day, regular intake of fish, low consumption of salt and sugar, and sufficient hydration<br>3x/week (15 min/session) computerised cognitive training<br>Oral and written information (individual consultations) on the respective risk factors and ways of reducing risk factors<br>Advice on setting individual goals for social activities | Standard care and General/untailored health advice |
| Lee, 2023 Japan | NA | 10 | Physical activity<br>Cognitive training<br>Social engagement | In person;<br>Group activites | Trained professionals and staff members | 1x/week (90min/session) aerobic exercise alone or as dual task (30 min/session) in large groups (total 40 sessions) (one/week) (aerobic exercise, no resistance training); recommendations to exercise independently and self-monitor step count; homework, feedback/comments from the staff 1x/2 weeks | General/untailored health advice (3x 1h sessions on nutrition, oral care, and healthy longevity) |

| RCT | | MULTIDOMAIN INTERVENTION | | | | | |
|---|---|---|---|---|---|---|---|
| **First author, year Country** | **RCT name** | **Intervention Duration (Months)** | **Multidomain Intervention components/Risk factors targeted** | **Mean of delivery and activity types** | **Delivered by** | **Brief description of activity frequency and duration** | **Control** |
| | | | | | | Social engagement via small group sessions (14 in total 2x/month, 1-2h/session) including, e.g., Book club and discussion activities | |
| Liu, 2023 China | COMBAT | 9 | Physical activity<br>Diet<br>Cognitive training<br>Mindfulness/ meditation | In person (most) and remote (due to the COVID-19 outbreak); Individual and groups activities | Trained research assistants, fitness instructors, physicians | 2x exercise group sessions (1h/session, 1x aerobic exercise, and 1x resistance training and balance); recommendation for daily independent exercise (1h/session)<br>1x diet group session + 2 individual diet consultations (including also some advice on CVD risk management)<br>20 cognitive group sessions (1h/session) including, e.g., cognitive exercises/games, educational presentations + participant provided with weekly homework<br>1x mindfulness/ meditation group session (1h), and 1x/week sessions (5 min/session) + recommendation to practice independently daily | Standard care |
| Roach, 2023 USA | COCOA | 24 | Physical activity<br>Diet<br>Cognitive stimulation<br>Social engagement<br>Sleep and stress | Mostly remote (telephone/email/text messages); Individual activities | Registered dietitians, certified nutritionists, trained staff, | Participant provided with individual coaching and support for:<br>US public health guidelines for physical activity<br>Dietary recommendations based on the MIND diet<br>US public health recommendations for sleep and stress management<br>Individualized recommendations for enhancing social interactions and brain stimulating activities | Standard care |

| RCT | | MULTIDOMAIN INTERVENTION | | | | | |
|---|---|---|---|---|---|---|---|
| **First author, year Country** | **RCT name** | **Intervention Duration (Months)** | **Multidomain Intervention components/Risk factors targeted** | **Mean of delivery and activity types** | **Delivered by** | **Brief description of activity frequency and duration** | **Control** |
| Chatterjee, 2022 India | MISCI | 6 | Physical activity<br>Diet<br>Cognitive training | In person and remote;<br>Individual activities | Psychologist, dietitian, physiotherapist | 40 sessions of 40 min each of computerised cognitive (increasing difficulty based on previous performance)<br>1x/week phone call and 1x/month individual consultations for diet (guidance to adhere to a mediterranean-equivalent diet)<br>2x/week supervised exercise for the first 6 months (including aerobic exercise resistance training (increasing intensity) | Less Intensive intervention (Health awareness instructions for brain stimulating activities such as sudoku, mental maths, learning music and new skills. Bimonthly monitoring of activities by phone and in person) |
| Kajita, 2022 Japan | NA | 30 | Physical activity<br>Diet<br>Cognitive training<br>Dementia literacy | In person;<br>Individual and group activities | Dietitian and other study staff | Intensive 10-week training (DPP: dementia prevention class 1-2x/week for 10 weeks), followed by booster including:<br>Physical activity training 1x/3 months (aerobic exercise and strength)<br>Additional nutrition education<br>Cognitive training including multiple tasks, e.g., of working memory, calculation, spotting the difference between two similar pictures and reading aloud + dual task training, exercise and cognitive tasks were performed simultaneously, such as calculation while walking<br>Dementia focused presentations explained the factors related to the incidence of dementia and recommended physical and cognitively engaging activities, communication with neighbors, a well-balanced diet and a moderate amount of sleep in daily life | Less Intensive intervention (10-week training (DPP) without booster) |

| RCT | | MULTIDOMAIN INTERVENTION | | | | | |
|---|---|---|---|---|---|---|---|
| **First author, year Country** | **RCT name** | **Intervention Duration (Months)** | **Multidomain Intervention components/Risk factors targeted** | **Mean of delivery and activity types** | **Delivered by** | **Brief description of activity frequency and duration** | **Control** |
| Yang, 2022 China | NA | 6 | Physical activity<br>Diet<br>Cognitive training<br>Cardio-metabolic risk monitoring | In person; Individual and groups activities | Community healthcare workers (dietician, rehabilitation therapist, psychotherapist, nurse) | 1-2x/week individualized progressive muscle strength training and aerobic exercise, group-based<br>1x/3–4 weeks individual diet consultations (10–30 min/session, 6 in total)<br>1x/week (60–90 min/session) computerised cognitive training under the guidance of a psychotherapists<br>4x nurse consultations for cardio-metabolic risk (baseline, M1, M3, M6) and accesso to chat with nurse | Standard care, and General/untailored health advice (3x 45' health education sessions) |
| de Souto Barreto, 2021, France | eMIND | 6 | Physical activity<br>Diet<br>Cognitive training | Remote; Individual activities | Exercise scientist, hospital dietitian, cognitive scientist, study physician | Personalised exercise programme (pre-recorded videos) 2x/week<br>Diet advice (pre-recorded videos), 2x/month, ~5–8 min/video<br>Computerised cognitive training 2x/week | General/untailored health advice (link to information on multidomain activities produced by the research team) |
| Moon, 2021 South Korea | SUPERBRAIN | 6 | Physical activity<br>Diet<br>Cognitive training<br>Vascular risk monitoring<br>Social engagement<br>Dementia literacy | In person; Individual and group activities (Facility Group)<br>Remote; Individual activities (Home Group) | Nurse, physician, dietician, psychologist, trained exercise professional, trained health professional | 3x/week (1h/session) exercise session, including aerobic, balance, and strength training, increasing intensity (Facility Group: all in group; Home Group mostly individually at home, following videos on PC or written handouts<br>3x individual diet sessions (30min/session), 7 group diet sessions (50min/session) (Facility Group); OR 3x individual sessions (30min/session), 4 group sessions (50min/session) and 3 independent sessions using a workbook at home (Home Group)<br>2x individual vascular risk consultation with study physician (including medication management, if needed), 6x individual nurse consultations (1x/4 | Standard care, General/untailored health advice (educational material) |

| RCT | | MULTIDOMAIN INTERVENTION | | | | | |
|---|---|---|---|---|---|---|---|
| **First author, year Country** | **RCT name** | **Intervention Duration (Months)** | **Multidomain Intervention components/Risk factors targeted** | **Mean of delivery and activity types** | **Delivered by** | **Brief description of activity frequency and duration** | **Control** |
| | | | | | | weeks), and educational material provided about risk factor management and lifestyle for dementia prevention (both groups)<br>Social engagement supported through the group meetings of the other intervention components, 1x/month once a month, social activities (e.g., theatre, meeting friends, etc.) were organized<br>4x (Facility Group) OR 3x (Home Group) educational sessions (50 min/session) for dementia literacy and including also motivation + possibility for weekly self-assessments. Facility group: 4x group sessions plus self-assessment; 1x independent workbook session at home (Home Group) | |
| Ng, 2021 Singapore | NA | 6 | Physical activity<br>Diet<br>Cognitive training | In person and remote;<br>Individual and groups activities | Psychologist, dietician, and other trained study staff | Multidomain programme with sessions 2x/week (48 sessions in total)<br>31% of the sessions focused on exercise (aerobic, resistance, and dual-task training; progressive programme, moderate intensity) conducted in small group sessions (1h or 30min per session)<br>1x/month (1h/session) in person diet group meetings; diet guidance and recommendations also provided by a dietician through a mobile application including, e.g., education modules, quizzes/games, option to upload photos of meals to receive direct feedback<br>69% of the multidomain programme sessions focused on cognitive training; first 12 weeks: 1x/week (1h/session) paper-based games/cognitive challenges in small groups + 1x/week (1h/session) independent computerised cognitive training session; next 12 weeks: | Standard care, General/untailored health advice (monthly 1h educational talk) |

| RCT | | MULTIDOMAIN INTERVENTION | | | | | |
|---|---|---|---|---|---|---|---|
| **First author, year Country** | **RCT name** | **Interventio n Duration (Months)** | **Multidomain Intervention components/Risk factors targeted** | **Mean of delivery and activity types** | **Delivered by** | **Brief description of activity frequency and duration** | **Control** |
| | | | | | | 2x/weeks of computerised cognitive training (1h or 2h per session) | |
| Chen, 2020 Taiwan | THISCE | 12 | Physical activity<br>Diet<br>Cognitive training<br>Vascular risk monitoring/Chronic disease prevention | In person; Group activities | Fitness coach, physical therapist, occupational therapist, dietician, other trained staff | 16 multidomain sessions in total (2h/session) where normally:<br>45min included aerobic, resistance and balance training (+ participants encouraged to independently engage in aerobic exercise for 150min/week)<br>15 min included diet educational presentation<br>1h included cognitive training<br>In 3-4 of these in session educational presentations on preventing/managing chronic disease (healthy aging, dementia, CVD risk factors, osteoporosis, sarcopenia) were conducted (30–60 min/session) | General/untailored health advice (~3 monthly telephone calls) |
| Kouzuki, 2020 Japan | NA | 6 | Physical activity<br>Cognitive training<br>Dementia literacy and lifestyle prevention | In person; Individual and group activities | Occupational therapist, nurse, adviser with knowledge of dementia prevention | 1x/month group sessions (2h/session) (6 in total) addressing all intervention components:<br>50min aerobic and strength training exercises, light-to moderate intensity<br>Cognitive training individual and in group, following a tailored plan (i.e., difficult and content adjusted for each participant) prepared by the trained instructor | No intervention |

| RCT | | MULTIDOMAIN INTERVENTION | | | | | |
|---|---|---|---|---|---|---|---|
| First author, year Country | RCT name | Intervention Duration (Months) | Multidomain Intervention components/Risk factors targeted | Mean of delivery and activity types | Delivered by | Brief description of activity frequency and duration | Control |
| | | | | | | 20 min pre-recorded educational presentations on dementia prevention with specific focus on lifestyle | |
| Xu, 2020 Hong Kong | NA | 6 | Physical activity<br>Diet<br>Cognitive training<br>Vascular risk monitoring | In person; Individual and group activities | Nurse, Tai Chi master or trained physical therapists | 3x/week group-based Tai Chi session (first 12 weeks 30min/session, 36 in total); after that, participants were advised to practice using a recording, either individually or in groups (of their choice, in settings outside the study)<br>2x physician and 3x nurse individual consultations, including vascular risk management, diet, and educational presentation of physical activity | General/untailored health advice at baseline |
| Bae, 2019 Japan | NA | 6 | Physical activity<br>Cognitive training<br>Social engagement | In person; Group activities | Staff members (trained non-health professionals recruited from the general population) | Intervention on all components was delivered together in 2x/week group sessions (90 min/session, 48 in total) structured as follows:<br>15 min condition check and stretching<br>60 min of physical, cognitive, or social activities,<br>15 min of report writing, and discussion of the next session's schedule | General/untailored health advice (2x 90' health education sessions) |

| RCT | | MULTIDOMAIN INTERVENTION | | | | | |
|---|---|---|---|---|---|---|---|
| **First author, year Country** | **RCT name** | **Interventio n Duration (Months)** | **Multidomain Intervention components/Risk factors targeted** | **Mean of delivery and activity types** | **Delivered by** | **Brief description of activity frequency and duration** | **Control** |
| Park, 2019 South Korea | NA | 6 | Physical activity<br>Diet<br>Cognitive training<br>Vascular risk monitoring<br>Social engagement | In person; Individual and group activities | Nurses, physiotherapists, and social workers, psychologist; physician | 4x group sessions (1.5h/session) of resistance training; step count checked and goals were set at each group sessions (so not just the exercise sessions); home tasks<br>1x group diet educational and training session (1.5h) including feedback on individual diet; home tasks<br>1x group session (1.5h) joint for cognitive training and social engagement including educational and training activities; home tasks<br>1x group session (1.5h) including educational and training activities on vascular risk, monitoring of risk factors and medication; home tasks<br>1x group session (1.5h) to support participants in for making a plan to sustain the modified lifestyle habits | Less intensive intervention (3-monthly health check-ups at the centre) |
| Piccirilli, 2019 Italy | NA | 6 | Physical activity<br>Cognitive training<br>Social engagement | In person; Group activities | Psychologist | 2x/week group sessions (60 min/sessions) including all intervention components<br>Physical activity was mostly aerobic exercise (walking at steady pace)<br>While walking participants listened to their preferred cognitive audios via headphones<br>After the simultaneous physical and cognitive training, participants held group discussion for the social engagement component | No intervention |
| Richard, 2019 The Netherlands, Finland, France | HATICE | 18 | Physical activity<br>Diet<br>Cardio-metabolic risk monitoring | Remote (with possibility of interaction with a remote health-coach); Individual activities | Trained health-coach (various health professional background) | Intervention delivered via an internet-based platform with remote support from a health-coach and including 7 modules (physical activity; diet; blood pressure; cholesterolemia; diabetes; overweight; smoking); modules included, e.g., educational content possibility of setting goals, monitoring activities and parameters, feedback; participants were invited and supported to | General/untailored health advice (access to a static platform, similar to the intervention one in appearance, with limited general health information) |

| RCT | | MULTIDOMAIN INTERVENTION | | | | | |
|---|---|---|---|---|---|---|---|
| **First author, year Country** | **RCT name** | **Intervention Duration (Months)** | **Multidomain Intervention components/Risk factors targeted** | **Mean of delivery and activity types** | **Delivered by** | **Brief description of activity frequency and duration** | **Control** |
| | | | | | | prioritise health-factors based on their individual risk profile assessed at baseline and during the study based on progress and changes | |
| Vanoh, 2019 Malaysia | NA | 6 | Physical activity<br>Diet<br>Cognitive stimulation<br>Cardio-metabolic risk monitoring<br>Dementia risk literacy | In person and remote;<br>Individual and groups activities | Trained field workers | The intervention was mainly delivered via a web-based application for the whole duration of the trial; participants were recommended to use the app ≥30min/day; the app included:<br>A 10-item screening tool for identifying risk of memory impairment<br>10 lifestyle-based guides control (focusing on blood sugar, control blood cholesterol, intake of fruits and vegetables, intake of fish, calorie restriction, mental stimulating activities, physical activity, smoking and alcohol, social activities and holistic health care)<br>Health Diary, a platform for users to save their blood test results<br>Healthy Food Menu, including tips for healthy meals, shopping guidelines, and nutrition related quiz<br>Group sessions were organised in the first 3 months (number not specified) to train and educate participants on the use of the web-app; browsing manual was also provided | General/untailored health advice |

| RCT | | MULTIDOMAIN INTERVENTION | | | | | |
|---|---|---|---|---|---|---|---|
| **First author, year Country** | **RCT name** | **Interventio n Duration (Months)** | **Multidomain Intervention components/Risk factors targeted** | **Mean of delivery and activity types** | **Delivered by** | **Brief description of activity frequency and duration** | **Control** |
| Ng, 2018 Singapore | Singapore Frailty Interventio n Trial | 6 | Physical activity<br>Diet<br>Cognitive Training | In person; Individual and groups activities | Psychologist, professional trainer, nurse facilitator | 2x/week exercise group sessions (90 min/session, 24 in total, for the first 12 weeks moderate) including gradually increasing intensity and tailored, progressive program of resistance training and balance, with dual-task elements; independent exercise was also encouraged; from week 13 home-based exercises (frequency not specified)<br>Daily multicomplex of vitamins/supplements (Fortisip® Multi Fibre, including iron and folate, vitamins B6, B12, D, and calcium)<br>1x/week 2h session of cognitive training/exercises/games. Next 12 weeks: 2h booster session every 2 weeks. In total, 18 sessions. | Standard care and community based social, recreational and day care rehabilitation services for older people, and placebo product to the active nutritional supplements |
| Andrieu, 2017 France, Monaco | MAPT | 36 | Physical activity<br>Diet<br>Cognitive training<br>Vascular risk monitoring | In person; Individual and groups activities | Psychologist s, physical activity instructors, nurses, physician | 12x multidomain sessions M1-M2 and 2x multidomain booster sessions (M12 and M24) including:<br>45min physical activity educational and motivational presentations<br>15min diet educational presentation<br>1h cognitive educational presentation and cognitive training<br>1x group session/month for either physical activity; diet; or cognitive training (M3-M36)<br>1x/6 months (M3-M36; 6 in total) multidomain individual motivational interviews<br>Independent exercise encouraged to meet guidelines of 150min/week moderate intensity exercise<br>3x individual consultations with the study physician focusing on vascular risk, but also | No intervention |

| RCT | | MULTIDOMAIN INTERVENTION | | | | | |
|---|---|---|---|---|---|---|---|
| **First author, year Country** | **RCT name** | **Intervention Duration (Months)** | **Multidomain Intervention components/Risk factors targeted** | **Mean of delivery and activity types** | **Delivered by** | **Brief description of activity frequency and duration** | **Control** |
| | | | | | | including overall health and wellbeing, as needed, e.g., oral health, functional status, mental health | |
| Maffei, 2017 Italy | Train the Brain | 7 | Physical activity<br>Cognitive training<br>Social engagement | In person; Group activities | Physiotherapists and professional trainer, other trained staff | 3x/week group exercise sessions (1h/session) including aerobic and muscle strength training (gradual intensity increase)<br>6x/week, group cognitive training sessions (1h/session) including educational presentations and paper-pen and computer-based training (gradual increase in exercises complexity);<br>1x/week group music therapy sessions (1h/session) including listening, singing, and playing musical instruments<br>In addition to the group sessions for the other intervention components, social engagement was supported with 1x/month session of movie watching and discussion | No intervention |
| Köbe, 2016 Germany | NA | 6 | Physical activity<br>Cognitive stimulation<br>Omega-3 fatty acid supplementation | In person; Individual and groups activities | Trained exercise leaders, a trained psychometrician, | 2x/week aerobic exercise sessions (45min/sessions)<br>AKTIVA cognitive stimulation programme (active cognitive stimulation–prevention in the elderly: Aktive Kognitive Stimulation–Vorbeugung im Alter; here adapted for MCI patients) is an approach to enhance cognitive activity in | Less intensive intervention (Omega-3 fatty acid stretching and toning condition) |

| RCT | | MULTIDOMAIN INTERVENTION | | | | | |
|---|---|---|---|---|---|---|---|
| **First author, year Country** | **RCT name** | **Intervention Duration (Months)** | **Multidomain Intervention components/Risk factors targeted** | **Mean of delivery and activity types** | **Delivered by** | **Brief description of activity frequency and duration** | **Control** |
| | | | | | | everyday life via encouraging the use of cognitively stimulating leisure activities and memory strategies, and conveying a positive attitude towards aging, disease, and self-perception.<br>2200 mg/day long-chain omega-3 fatty acid (4 capsules comprising 1320 mg eicosapentaenoic acid, 880 mg docosahexaenoic acid and additional 15mg of vitamin E) | |
| Moll van Charante, 2016, The Netherlands | preDIVA | 72 | Any modifiable vascular/metabolic/lifestyle risk factors linked to dementia based on each participant's specific risk profile | In person; Individual activities | General practice nurse | 1x/4months nurse consultation (18 in total). The consultations included individually tailored lifestyle advice according to a detailed protocol conforming with prevailing Dutch general practitioner guidelines on cardiovascular risk management (e.g., diet, physical activity, smoking, VBMI, blood pressure, diabetes, cholesterol). Initiation/optimisation of drug treatment for hypertension, dyslipidaemia, and type 2 diabetes mellitus was initiated or optimised, if indicate. | Standard care, according to the prevailing standards for cardiovascular risk management |
| Clare, 2015 UK | Agewell | 12 | Physical activity<br>Diet<br>Cognitive training<br>Social engagement | In person and remote; Individual activities | Trained researchers | Baseline interview (up to 90 min), where the focus was on goal setting (discuss and identify goals for the year in different lifestyle areas, physical activity was one of the areas)<br>5x bi-monthly mentoring telephone calls to, e.g., review progress, provide extra support<br>Follow-up interview at M12 | General/untailored health advice (2x 90' health education sessions) |

| RCT | | MULTIDOMAIN INTERVENTION | | | | | |
|---|---|---|---|---|---|---|---|
| **First author, year Country** | **RCT name** | **Intervention Duration (Months)** | **Multidomain Intervention components/Risk factors targeted** | **Mean of delivery and activity types** | **Delivered by** | **Brief description of activity frequency and duration** | **Control** |
| Ngandu, 2015 Finland | FINGER | 24 | Physical activity<br>Diet<br>Cognitive training<br>Cardio-metabolic risk monitoring<br>Social engagement | In person; Individual and group activities | Nurse, physician, nutritionist, psychologists, physiotherapist | Up to 3x/week muscle strength (mostly in group); up to 5x/week aerobic exercise (independent)<br>3x individual diet consultations; 7-9x diet group meetings<br>11x cognitive group sessions; 72x2 independent cognitive training sessions<br>3x nurse consultations; 3x physician consultations for cardiometabolic risk | General/untailored health advice |
| Johari, 2014 Malaysia | NA | 12 | Physical activity<br>Diet<br>Cognitive training | In-person; Group activities | Trained researchers; clinical psychologist | Monthly aerobic exercise group session<br>Monthly diet group session dietary providing recommendations, diet counselling, a food quiz, and demonstration of healthy food preparation<br>Group sessions including crossword puzzle, and board games etc.<br>(sessions frequency not specified) | Less intensive intervention (12 months supplement isocaloric placebo - 3000 mg corn oil/day - as part of a larger study where they are control/ placebo) |
| Lee, 2014 South Korea | PASCAL | 18 | Physical activity<br>Diet<br>Cognitive training<br>Vascular risk monitoring<br>Social engagement | In person or remote (depending on the intervention group allocation[a]); individual activities | Nurses | Health education, personalised advice, goal setting, encouragement for independent exercise, healthy diet, engage in cognitive activity, alcohol consumption and smoking; treatment adjustments according to evidence-based guidelines; education about the importance of social connectedness; health education, information about dementia prevention and role of lifestyle in supporting cognitive health. Depending on the group allocation[a] the intervention was delivered bimonthly (1), monthly (2) on the phone, or in person bimonthly with (3) or without (4) rewards. | Standard care (at the senior citizen centres) |
| Koekkoek, 2012 | ADDITION | 72 | Physical activity<br>Diet | In person; individual activities | Neuropsychologist and general practitioners | Intervention via individual consultations (frequency not specified) | Standard care, (according to the Dutch college of |

| RCT | | MULTIDOMAIN INTERVENTION | | | | | |
|---|---|---|---|---|---|---|---|
| First author, year Country | RCT name | Intervention Duration (Months) | Multidomain Intervention components/Risk factors targeted | Mean of delivery and activity types | Delivered by | Brief description of activity frequency and duration | Control |
| The Netherlands | | | Cardio-metabolic risk monitoring and control | | | Protocol driven, including medications where necessary to maintain the following target: (a) HbA1c<53 mmol/mol (7.0%); (b) blood pressure ≤120/80 mmHg; (c) total cholesterol ≤3.5 mmol/L<br>Advice and counselling were provided for all lifestyle-based intervention components (e.g., physical activity, diet, smoking) | general practitioners) |

a. Intervention 1: Telephonic care management bimonthly; Intervention 2: Telephonic care management monthly; Intervention 3: Health-worker initiated visit care management bimonthly; Intervention 4: Intervention 3 + rewards

CVD: Cardiovascular Disease; DPP: Dementia Prevention Programme GP: General Practitioner; MIND: Mediterranean-DASH Intervention for Neurodegenerative Delay; NA: Not available.

#### Supplementary Table 7.
**Egger's test for assessment of Publication Bias**

| Outcome | beta-coefficient | SE | P-value |
|---|---|---|---|
| Global cognition from composite score | 2.329 | 0.870 | **0.007** |
| Executive function | -0.266 | 0.324 | 0.411 |
| Memory | 0.072 | 0.312 | 0.817 |
| Processing speed & Attention | 0.070 | 0.321 | 0.823 |
| Global cognition using screening tools for dementia/cognitive impairment | 0.640 | 0.347 | 0.065 |
| CDR Sum of Boxes | -0.781 | 0.757 | 0.302 |
| Incident Dementia | 0.360 | 1.008 | 0.718 |

CDR: Clinical dementia rating; SE: Standard error.

**Supplementary Table 8.**
**GRADE assessment of the evidence for the effect multidomain intervention on global cognition**

| Outcome (N included RCTs) | N Participants | Risk of bias | Inconsistency | Indirectness | Imprecision | Other considerations | Effect Absolute (95% CI) | Certainty |
|---|---|---|---|---|---|---|---|---|
| Global cognition as composite score of validated tests (25) | 17,082 | Not serious | Very Serious | Not detected | Not detected | Not detected | SMD: 0.28 SD higher (0.10 higher to 0.45 higher) | ⊕⊕◯◯ (Low) |
| Memory (33) | 20,0769 | Not serious | Not serious | Not detected | Not detected | Not detected | SMD: 0.06 SD higher (0.01 higher to 0.10 higher) | ⊕⊕⊕⊕ (High) |
| Executive Function (27) | 16,392 | Not serious | Not serious | Not detected | Not detected | Not detected | SMD: 0.06 SD higher (0.02 higher to 0.10 higher) | ⊕⊕⊕⊕ (High) |
| Processing speed (25) | 10,223 | Not serious | Not detected | Not detected | Not detected | Not detected | SMD: 0.04 SD higher (0.00 higher to 0.08 higher) | ⊕⊕⊕⊕ (High) |
| Global cognition using screening tools for dementia/cognitive impairment (23) | 10,223 | Not serious | Not detected | Not detected | Not detected | Not detected | SMD: 0.08 SD higher (0.03 higher to 0.13 higher) | ⊕⊕⊕⊕ (High) |

#### 4. Supplementary Figures

##### Supplementary Figure 1.

**Risk of bias assessment for the Cognitive Function outcome displayed as percentage contribution for each risk of bias domain; overall risk of bias; and all domains combined (a) and detailed ratings per study (b).**

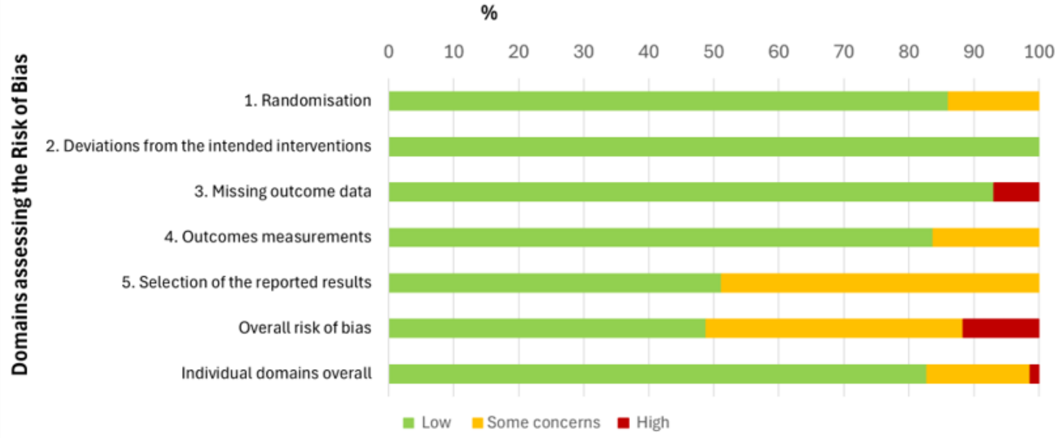

a

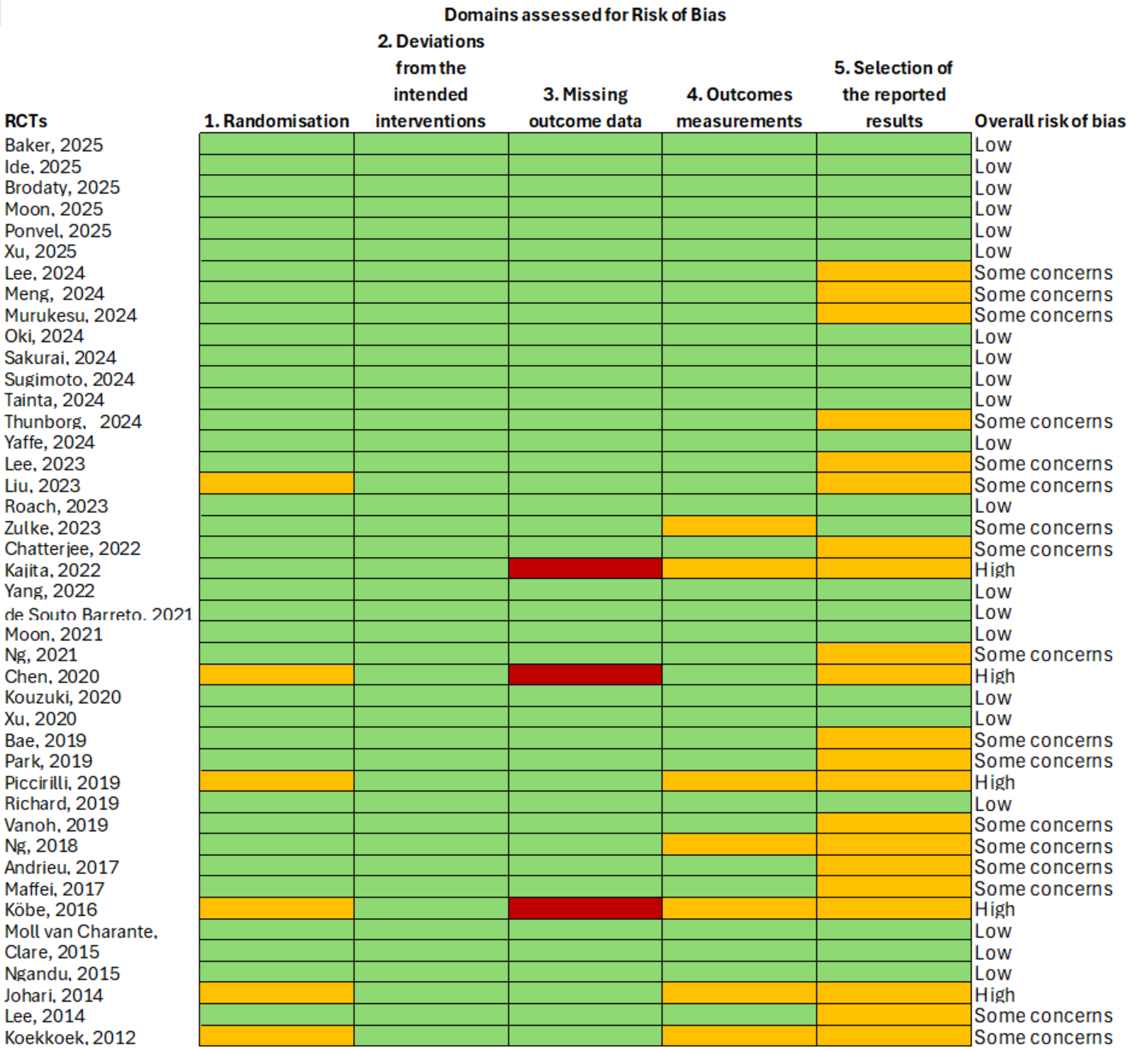

b

**Supplementary Figure 2.**

**Funnel plot of global cognition measured as a composite score of validated neuropsychological tests, based on pooled analysis of all eligible studies, for investigation of indicators potential publication bias.**

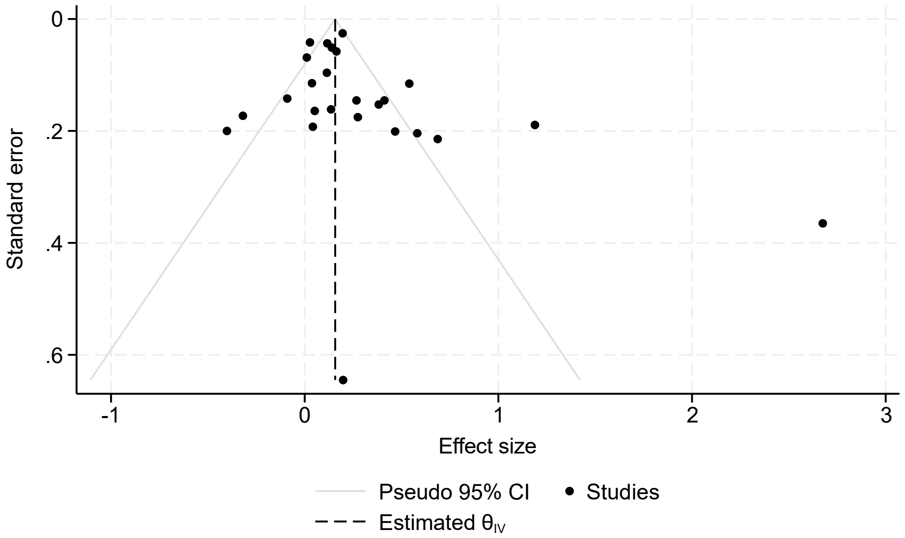

**Supplementary Figure 3.**

**Subgroup analysis of the effect of multidomain interventions on global cognition (composite score) by intervention duration (a); intervention intensity (b); number of modifiable risk factors used for participant selection also addressed in the multidomain intervention (level of target populations selection) (c); or target population cognitive status (d).**

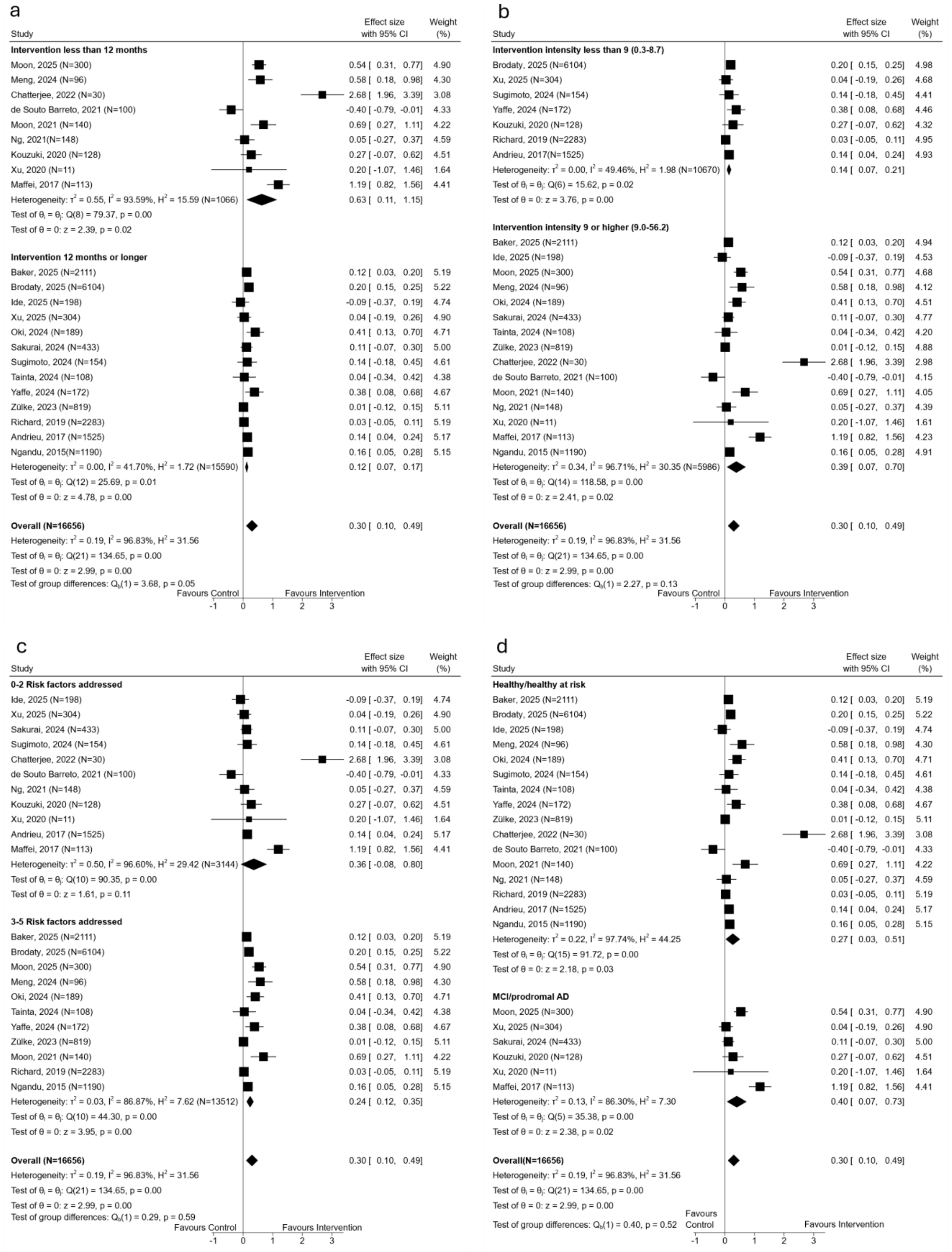

#### Supplementary Figure 4.

**Sensitivity analysis on the effect of multidomain interventions on global cognition measured as a composite score of validated neuropsychological tests, by excluding three RCTs identified as important sources of heterogeneity (a); and by excluding all RCTs with N<100 (b).**

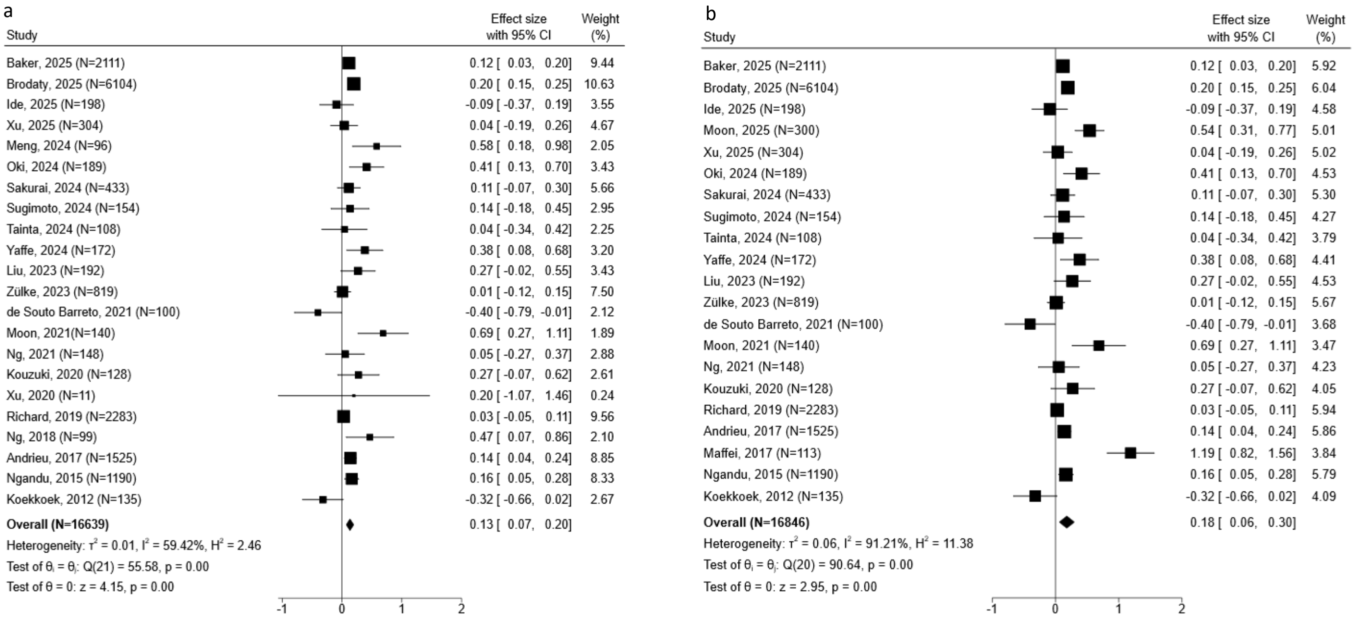

#### Supplementary Figure 5.

**Effect of multidomain interventions on memory (a); executive function (b); or processing speed (c).**

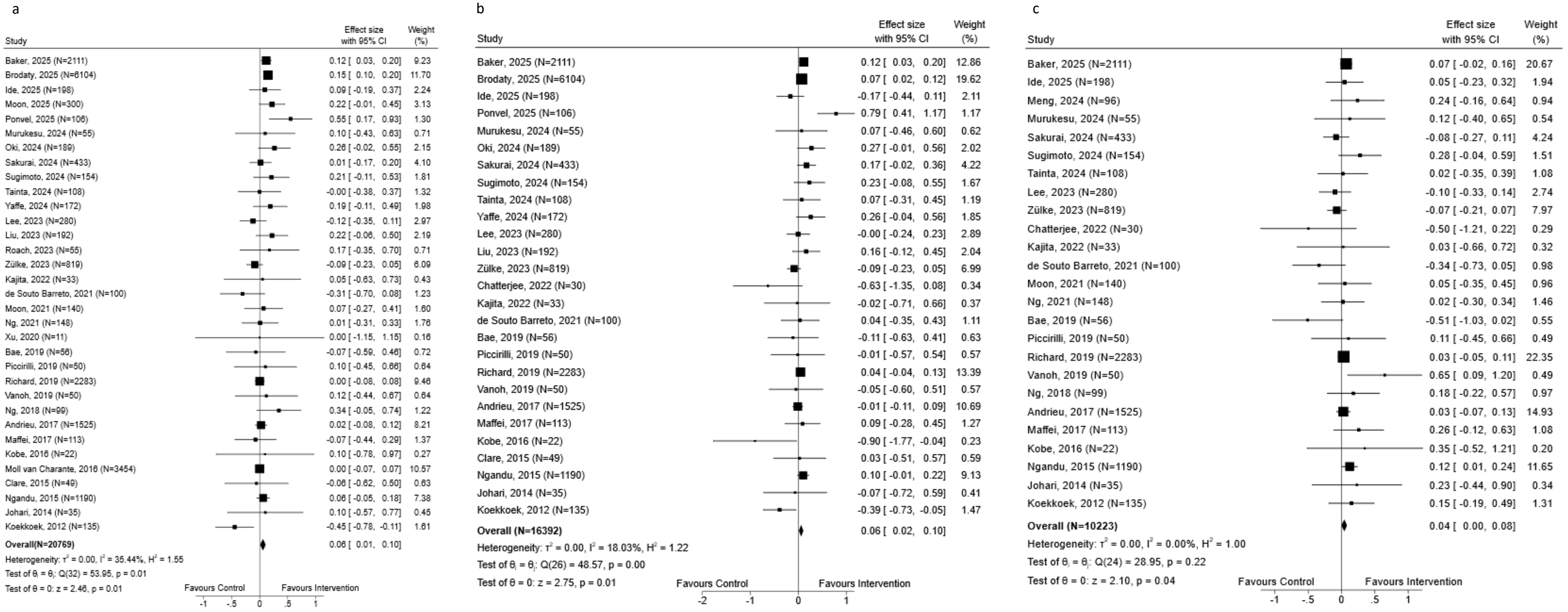

#### Supplementary Figure 6.

**Effect of multidomain interventions on global cognition measured screening tools for cognitive impairment and dementia (a); or Clinical Dementia Rating – Sum of Boxes (b).**

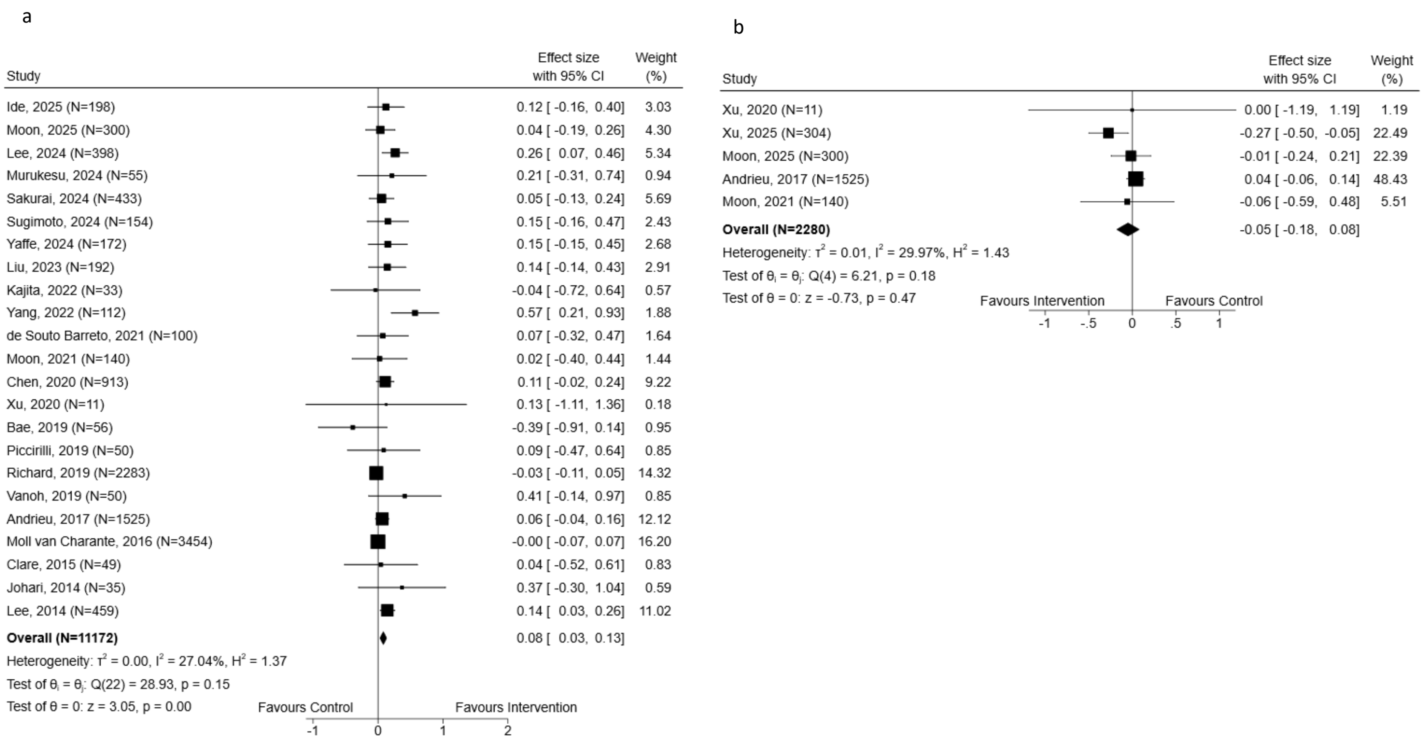

#### Supplementary Figure 7.
##### Multidomain interventions effects on incident dementia

| Study | Intervention: Dementia | Intervention: No Dementia | Control: Dementia | Control: No Dementia | Log odds-ratio with 95% CI | Weight (%) |
|---|---|---|---|---|---|---|
| Ide, 2025 | 2 | 97 | 0 | 99 | 6.52 [ -29.29, 42.34] | 0.01 |
| Moll van Charante, 2016 | 121 | 1,732 | 112 | 1,489 | -0.07 [ -0.34, 0.19] | 99.99 |
| **Overall** | | | | | -0.07 [ -0.34, 0.19] | |

Heterogeneity: $\tau^2 = 0.00$, $I^2 = 0.00\%$, $H^2 = 1.00$

Test of $\theta_i = \theta_j$: $Q(1) = 0.13$, $p = 0.72$

Test of $\theta = 0$: $z = -0.54$, $p = 0.59$

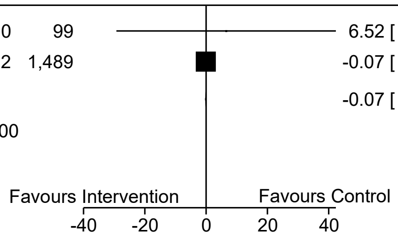
